# Cost-effectiveness of food fortification for reducing global malnutrition: a systematic review of economic evaluations across 63 countries

**DOI:** 10.1101/2025.06.12.25329169

**Authors:** Elise Cogo, Ferruccio Pelone, Helena Pachón, Brian Buckley, Maria Christou, Gemma Villanueva, Monica Woldt, Nicholas Henschke, Becky L Tsang

## Abstract

**Background:** Fortification, the addition of essential micronutrients during food processing, reduces mortality and malnutrition. Our objective was to comprehensively synthesize global evidence on the cost-effectiveness (CE) and cost savings of food fortification.

**Methods:** We employed systematic review methodology, PROSPERO registration (CRD42023493795), searching six databases to January 2024. Eligible studies included economic analyses comparing staple food post-harvest micronutrient fortification to no fortification. Quality appraisal used Philips’ modeling framework. We converted incremental cost-effectiveness ratios (ICERs) to 2022 US$; and synthesized the data overall and by micronutrient. For illustration, findings were also categorized by “hypothetical” CE thresholds based on common example percentages of gross domestic product per capita (GDP pc) per country.

**Findings:** After screening 6,425 abstracts, 56 studies in 66 reports were included, reporting >200 analyses. Sixty-three countries were represented, including >40 low- and middle-income economies (LMICs). Most frequent interventions were: vitamin A, folic acid, iron, and iodine added to cereal grains/products (e.g., flours) and condiments (e.g., oils, sugar, salt). Models were heterogeneous and employed various perspectives. Most evaluations (58%; 135/232) had ICERs less than $150 per disability-adjusted life year (DALY) averted (or healthy life year gained). We found 87% (201/232) overall were within a hypothetical CE threshold of “50% GDP pc”. With an example “35% GDP pc” level among LMICs, 84% (190/227) were estimated to be cost-effective; and 71% (37/52) were less than “20% GDP pc” among low-income countries. Additionally, six out of eight cost-utility studies’ ICERs were dominant. Moreover, 47 total unique benefit–cost ratios found benefits outweighed costs, ranging from 1·50:1 to 100·6:1.

**Interpretation:** Food fortification programs are likely cost-effective in the majority of contexts. While cost-effectiveness evaluations are specific to local factors and methodology, this research can assist with evidence-informed decision-making for global health policy and priority setting, particularly in resource-constrained economies.

**Funding:** U.S. Agency for International Development.

**Research in context:** *Evidence before this study:* Database searches of MEDLINE, Embase, EconLit, and the National Health Service Economic Evaluation Database (NHS EED), from inception to January 2024, were conducted for previous systematic reviews on the economics of food fortification. No language, date, or publication status limits were applied. While the impact of food fortification on health outcomes has been widely studied, we did not find a comprehensive systematic review of the cost-effectiveness of all types of large-scale food fortification. Several smaller systematic reviews and some in-depth narrative reviews have studied the economics of food fortification but their aims were not broad evaluations of its cost-effectiveness as they limited their scope to specific foods or nutrients.

*Added value of this study:* This large systematic review of 56 economic studies reporting over 200 analyses provides a substantially broader synthesis of the economic evidence base of food fortification. Of note, it also spans 63 countries, including >40 low- and middle-income economies (LMICs), thereby augmenting the international research on a system-level intervention for reducing global malnutrition, mortality, and morbidity. We found that food fortification programs are likely cost-effective in the majority of contexts. Overall, across the many diverse economic models, incremental cost-effectiveness ratios (ICERs) for most evaluations (58%; 135/232) were less than $150 per disability-adjusted life year (DALY) averted (or healthy life year gained). As an illustration using a “hypothetical” cost-effectiveness threshold example of ‘35% of gross domestic product (GDP) per capita’ among LMICs, 84% (190/227) of ICERs were estimated to be cost-effective. Additionally, six out of eight cost-utility studies’ (i.e., measuring quality-adjusted life years, QALYs) ICERs were dominant (i.e., fortification was less costly and more effective than the comparator). Moreover, 47 benefit–cost ratios found that food fortification programs’ benefits outweighed the costs (with ranges from 1·50:1 to 100·6:1).

*Implications of all the available evidence:* There are significant policy implications from this research. The decision by policymakers to enact or strengthen food fortification programs in their countries is predicated by many factors, including costs and cost-effectiveness. Synthesizing the evidence of the economic implications of food fortification could translate to improved global advocacy efforts by partners seeking to introduce and scale up food fortification programs. Nutrition for Growth aims to mobilize governments, bilateral agencies, private investors, businesses, civil society, donors, and others to increase and sustain their funding for nutrition actions, especially those that are evidence-based. Through the *Global Nutrition Report,* such financial commitments are being tracked internationally. The expectation is that this more robust evidence base of the cost-effectiveness of large-scale food fortification will encourage greater investment in initiating and strengthening fortification programs where they are needed. While cost-effectiveness evaluations are specific to local factors and methodology, this research can assist with evidence-informed decision-making for global health policy and priority setting, particularly in resource-constrained economies.

## INTRODUCTION

Micronutrient deficiencies (MNDs) are one of the most widespread forms of malnutrition, affecting millions worldwide and leading to substantial mortality and morbidity.(1) Also termed hidden hunger, an important cause of MNDs is inadequate intake of essential vitamins or minerals. Children and pregnant women are particularly susceptible, and globally it is estimated that 56% of children 6–59 months and 69% of non-pregnant women 15–49 years suffer from MNDs.(2) MNDs can worsen and be particularly severe during times of food insecurity and rising food prices, as vulnerable populations struggle to consume varied and nutritious diets.(3)

Large-scale food fortification (LSFF) (conducted at the post-harvest, food processing stage) is a system-level intervention defined as, “…deliberately increasing the content of essential micronutrients—i.e., vitamins and minerals (including trace elements)—in a food so as to improve the nutritional quality of the food supply and to provide a public health benefit with minimal risk to health.”(4) In 2023, the World Health Assembly adopted a resolution on accelerating efforts to prevent MNDs through safe and effective LSFF. The resolution “urges Member States to make decisions on food fortification with micronutrients and/or supplementation and to consider ways of strengthening financing and monitoring mechanisms.”(5)

While the impact of food fortification on health outcomes has been widely studied,(6, 7) its implications on economic outcomes is more limited, as a comprehensive systematic review of economic outcomes related to all types of LSFF has not been published. The 2008 Copenhagen Consensus ranked several food fortification interventions among the top five cost-effective priorities of micronutrient interventions.(8) Other systematic reviews(9–13) and in-depth narrative reviews(14–18) have been published about food fortification economics, but their aims were not broad systematic evaluations of its cost-effectiveness as they limited their scope to specific foods or nutrients.

Synthesizing the evidence of the economic implications of LSFF could translate to improved global advocacy efforts by partners seeking to introduce and scale up food fortification programs, particularly in resource-constrained economies. Our objective was to evaluate, comprehensively synthesize, and critically appraise the evidence on the economic outcomes associated with LSFF.

## METHODS

We conducted a systematic review following guidance from the Cochrane Handbook,(19) Campbell and Cochrane Economics Methods Group,(20) and PRISMA 2020.(21) Protocol details were registered prospectively (PROSPERO CRD42023493795).

### Search strategy and selection criteria

Inclusion criteria were studies evaluating the consumption (modeled or actual) of fortified staple foods (inclusive of beverages and condiments) against economic outcomes (all populations). Food fortification as defined above. The intervention was compared to unfortified food, no fortification at the time of evaluation (including voluntary and low levels of coverage), or pre-fortification period. All comparative study designs reporting economic outcomes were eligible, including controlled studies and economic models (mathematical/simulation).

Economic outcomes comprising cost–effectiveness (CE), cost–utility, and cost–benefit measures were eligible, such as: incremental cost-effectiveness ratios (ICERs), including incremental cost per disability-adjusted life year (DALY) averted and incremental cost per quality-adjusted life year (QALY) gained; cost–benefit ratios; and net monetary/economic benefits (e.g., increase in income level). World Health Organization (WHO)’s Choosing Interventions that are Cost-Effective (CHOICE) methodology advises that cost per DALY averted (also called healthy life year gained) is the preferred metric in low- and middle-income settings.(22) DALYs are measures of disease burden calculated based on years of life lost and years lived with disability. ICER measures the incremental costs per incremental health effect, as: ICER = [Difference in Costs] ÷ [Difference in Health Effects] for intervention versus comparator. The lower the ICER, the more cost-effective an intervention is (i.e., the less it costs per specified benefit).

We excluded the following interventions: biofortification; home fortification; individual supplementation; addition of non-micronutrients (e.g., probiotics, fiber); or products with added macronutrients (e.g., infant formula, meal replacements, therapeutic foods). Additionally, ‘head-to-head’ comparisons of fortification interventions compared to each other (i.e., without a non-fortified control) and studies with no comparison group were excluded.

An experienced information specialist conducted comprehensive literature searches, regardless of publication status (published or unpublished/grey literature). No date or language limits were applied. We searched six electronic databases from inception to January 17, 2024: MEDLINE (Ovid), Embase (Ovid), Cochrane Central Register of Controlled Trials (CENTRAL; via EBM Reviews in Ovid), EconLit (EBSCOhost), NHS EED (via University of York Centre for Reviews and Dissemination), and Tufts Medical Center Cost-Effectiveness Analysis (CEA) Registry. The main multi-database search strategy is in the Appendix. Supplemental searching included scanning reference lists of systematic reviews and key websites.

Two authors independently screened abstracts and full-text articles using Distiller SR software (v2.35); a third author resolved disagreements.

### Data analysis

Data extraction was conducted using predefined, piloted forms in Distiller SR, with one author extracting and a second author cross-checking data. We resolved disagreements by consensus. For studies reporting multiple economic outcome measures, to avoid double-counting of results, only the top-level health outcome was extracted based on the following hierarchy preference: DALY/QALY; deaths; a clinical condition (e.g., neural tube defects/cancers/anemia); a nutrition measure (e.g., deficiency/adequate intake). For example, if a study reported multiple ICERs for the same intervention, such as cost per DALY and cost per birth defect, we only extracted cost per DALY.

In addition to outcomes data, we extracted: study characteristics (i.e., country, setting, population, study design, time horizon, funding, conflict of interest); intervention and comparator details (i.e., micronutrient, food vehicle, dose, delivery platform, regulatory context); and economic context (i.e., currency, price year, analytic perspective, types of costs captured). The economic “perspective” is the point of view of an analysis; for example, a social welfare perspective considers costs and benefits for the whole society, and a healthcare perspective includes costs and effects on the health sector.

Methodological quality assessment was conducted per economics best practices using the following checklist tools, depending on study design: Philips’ modeling framework for economic models;(23) and Consensus Health Economic Criteria (CHEC) list for primary studies.(24) The Philips checklist is a broad framework of >50 questions divided into 15 sections that assesses three key modeling dimensions: ‘structure’ of the economic model, ‘data’ used, and model ‘consistency’.

For comparability of results, especially across low- and middle-income economy countries (LMICs), all ICERs were converted to 2022 US$ (or in a few cases to 2020/2021 when recent rates were not posted) using the following procedure based on Turner et al. (2019):(25) first, the local currency was inflated to year 2022 using local inflation rates from GDP implicit price deflators (World Bank); then exchanged to US$ using World Bank local currency rates. For non-USA studies reporting in US$ or International dollars, results were first converted to local currency for the study period, then inflated, and finally exchanged back to US$. When study results were only reported as multinational regional estimates, conversion rates used were population-weighted averages for the available countries as an approximation for the region. We obtained rates for a few missing countries from the International Monetary Fund website.

We tabulated results, categorized, and presented them in summary tables, along with appendices of data provided for each study. A map displaying study countries was created using Excel (Microsoft 365). Evidence was presented overall and also grouped by micronutrient. Due to substantial heterogeneity across the diverse models, meta-analysis was not appropriate.

Guidance on determining cost-effectiveness has advanced over the past two decades, and the use of generic cost-effectiveness thresholds as decision rules is not recommended; instead, WHO-CHOICE (2021) advises to use cost-effectiveness analysis as part of a transparent and fair decision-making process.(22) Especially for resource-constrained countries, each country needs to judge local cost-effectiveness amid resource allocation based on a range of factors specific to their context. There are a variety of methodologies for developing CE thresholds, and recent research provides alternative frameworks for countries to estimate them. From these new developments, for illustration and knowledge translation purposes we also present our results according to “hypothetical” CE thresholds based on example percentages of gross domestic product per capita (GDP pc; World Bank) per country, while acknowledging that these examples are not applicable to all contexts. Specifically, these were calculated per ICER for each study country using three “common examples” of 50%, 35%, and 20% of GDP pc (i.e., 0·5, 0·35, and 0·2 x GDP pc per country).(26–28)

## RESULTS

After de-duplication, two authors screened 6,425 unique abstracts and 1,512 full-texts (comprising 1,498 from databases plus 14 reports from supplemental searches) for eligibility. Ultimately, 56 studies in 66 reports(8, 9, 29–92) were included in our review (including 10 companion reports or studies that have been updated); figure 1 provides exclusion reasons and PRISMA flow diagram. From the 56 studies, >200 evaluations (analyses) were included since several studies reported relevant data on multiple different interventions, countries, etc. In particular, one very large study was conducted by Fiedler & Macdonald (2009)(48) that reported 171 total included ICERs from six interventions in 46 countries.

**Figure 1.**
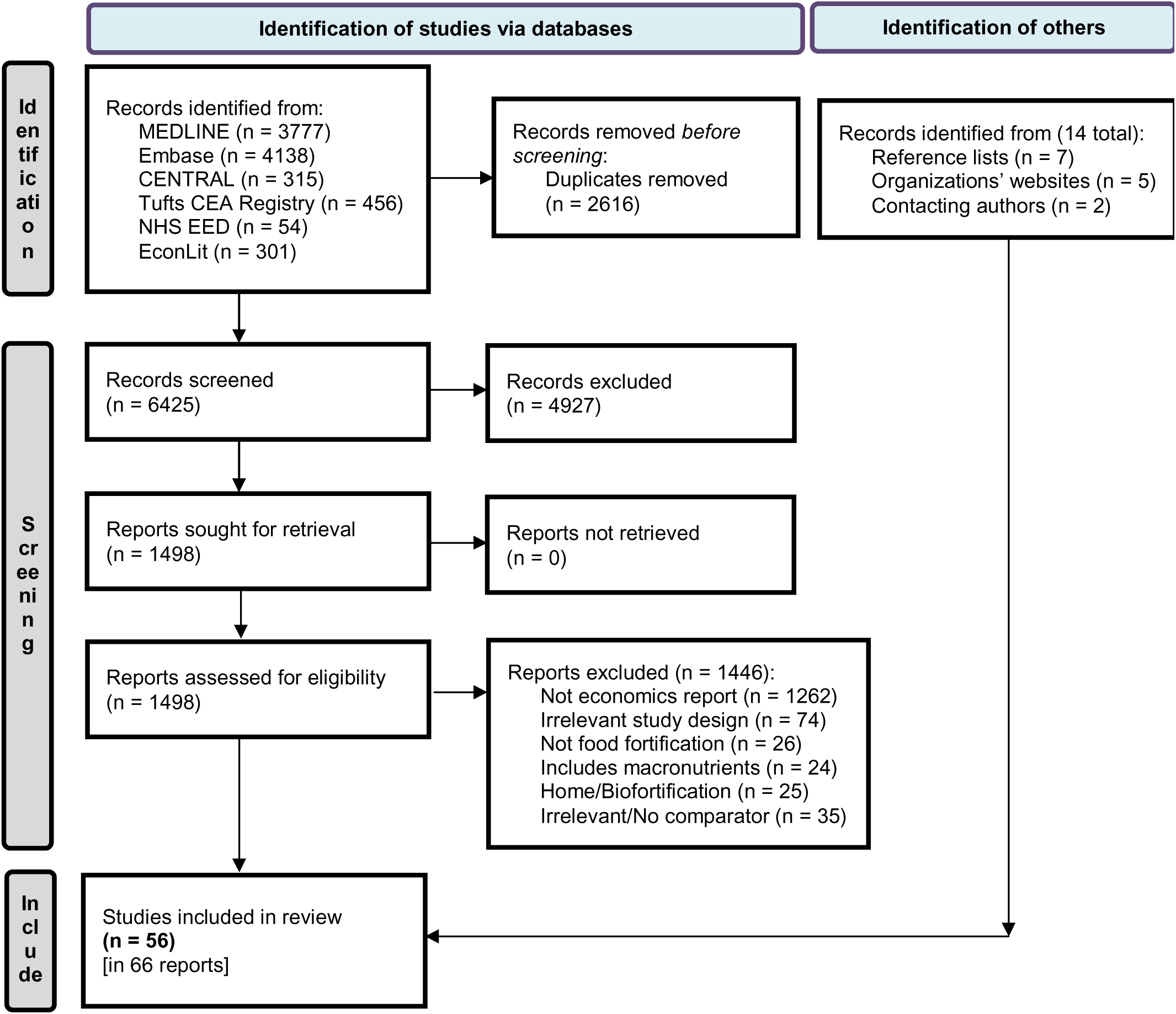
PRISMA flow diagram indicating searching results and study selection details

Across studies, 63 countries were represented (including 46 LMICs in the DALYs analysis), as shown in figure 2. India, China, Ethiopia, Bangladesh, and Australia were included in the most studies (with 5–8 each) (table 1). Analyses also included four studies that reported on multinational regions,(8, 32, 41, 51) which spanned all six WHO regions in total. Table 1 presents the number of studies per country, grouped by region, along with their World Bank economy income classification and GDP pc (in 2022 US$).

**Figure 2.**
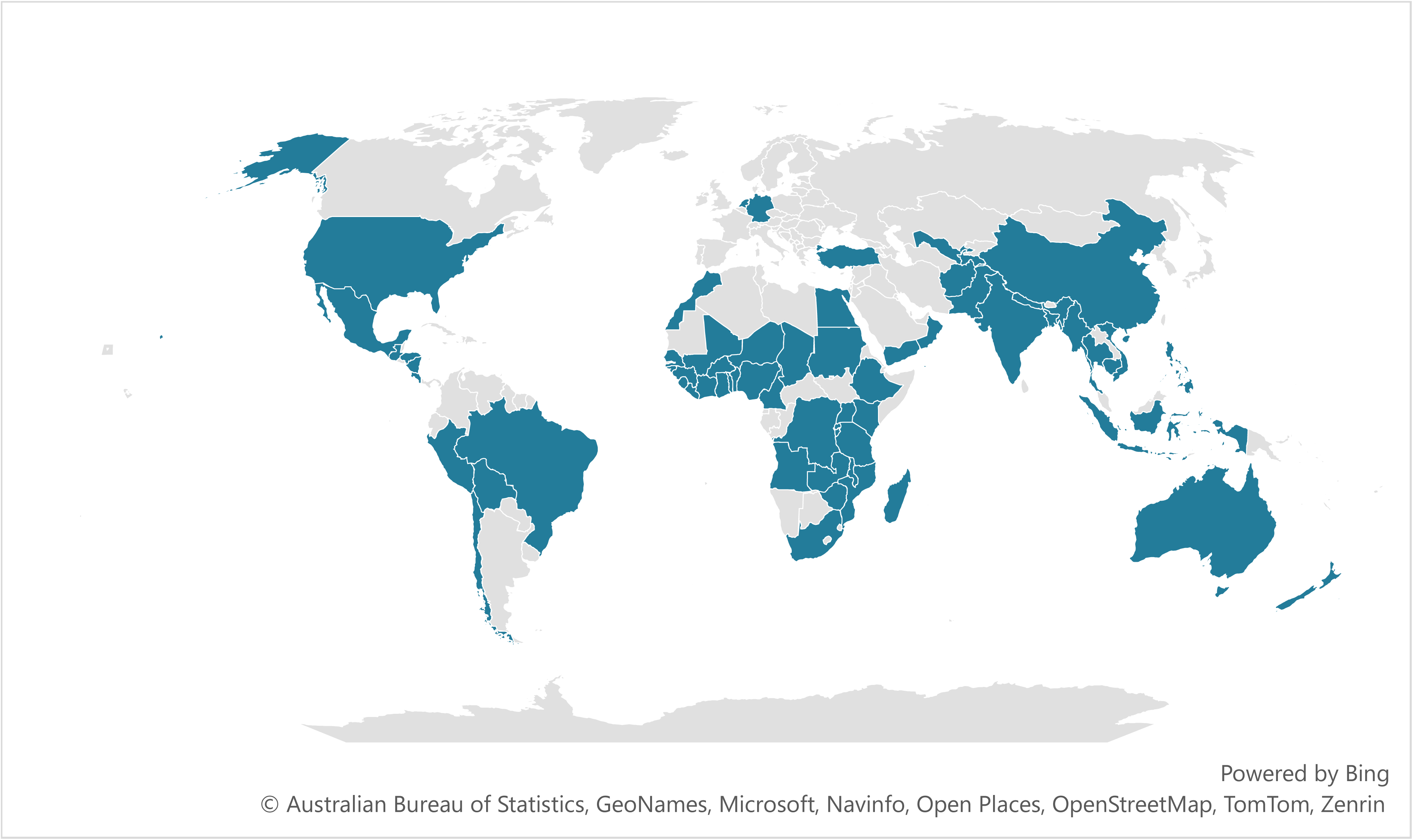
Map of 63 countries, including >40 LMICs, represented in the studies

**Table 1.** Study countries and regions, economy income classification, and GDP per capita.

| Countries,<br>by WHO Region | Studies*<br>(n) | Economy<br>classification<sup>†</sup> | GDP per capita<br>(2022 US\$)<sup>‡</sup> |
| --- | --- | --- | --- |
| **AFRICA** |  |  |  |
| Angola | 2 | LM | 2,933 |
| Benin | 2 | LM | 1,305 |
| Burkina Faso | 2 | L | 830 |
| Burundi | 1 | L | 259 |
| Cameroon | 3 | LM | 1,563 |
| Chad | 1 | L | 699 |
| Congo, Democratic Republic | 1 | L | 665 |
| Côte d'Ivoire | 3 | LM | 2,492 |
| Ethiopia | 5 | L | 1,027 |
| Ghana | 2 | LM | 2,218 |
| Guinea | 1 | LM | 1,515 |
| Guinea-Bissau | 1 | L | 814 |
| Kenya | 1 | LM | 2,099 |
| Liberia | 1 | L | 755 |
| Madagascar | 1 | L | 517 |
| Malawi | 1 | L | 643 |
| Mali | 3 | L | 831 |
| Mozambique | 1 | L | 558 |
| Niger | 2 | L | 589 |
| Nigeria | 2 | LM | 2,163 |
| Rwanda | 1 | L | 967 |
| Senegal | 2 | LM | 1,595 |
| Sierra Leone | 1 | L | 476 |
| South Africa | 3 | UM | 6,766 |
| Tanzania | 3 | LM | 1,193 |
| Togo | 1 | L | 923 |
| Uganda | 2 | L | 964 |
| Zambia | 4 | LM | 1,457 |
| Zimbabwe | 1 | LM | 1,677 |
| **AMERICAS** |  |  |  |
| Bolivia | 2 | LM | 3,600 |
| Brazil | 2 | UM | 9,065 |
| Chile | 2 | H | 15,411 |
| Costa Rica | 1 | UM | 13,365 |
| Guatemala | 2 | UM | 5,473 |
| Honduras | 1 | LM | 3,012 |
| Mexico | 1 | UM | 11,477 |
| Nicaragua | 1 | LM | 2,252 |
| Peru | 1 | UM | 7,239 |
| USA | 3 | H | 77,247 |
| **EASTERN MEDITERRANEAN** |  |  |  |
| Afghanistan | 2 | L | 353 |
| Egypt | 3 | LM | 4,295 |
| Morocco | 1 | LM | 3,442 |
| Oman | 1 | H | 25,057 |
| Pakistan | 4 | LM | 1,589 |
| Sudan | 1 | L | 1,102 |
| Yemen | 1 | L | 699 |
| **EUROPE** |  |  |  |
| Germany | 3 | H | 48,718 |
| Netherlands | 1 | H | 57,025 |
| Tajikistan | 2 | LM | 1,076 |
| Turkey | 1 | UM | 10,675 |
| Uzbekistan | 1 | LM | 2,276 |
| **SOUTH-EAST ASIA** |  |  |  |
| Bangladesh | 5 | LM | 2,688 |
| India | 8 | LM | 2,366 |
| Indonesia | 3 | UM | 4,788 |
| Myanmar | 1 | LM | 1,149 |
| Nepal | 1 | LM | 1,348 |
| Thailand | 1 | UM | 6,913 |
| **WESTERN PACIFIC** |  |  |  |
| Australia | 5 | H | 65,078 |
| Cambodia | 1 | LM | 1,760 |
| China | 7 | UM | 12,663 |
| New Zealand | 3 | H | 48,217 |
| Philippines | 3 | LM | 3,499 |
| Vietnam | 2 | LM | 4,179 |
| TOTAL | 131 | — | — |

\*Numbers sum to >56 as some studies reported on >1 country.
<sup>†</sup>L=low income, LM=lower-middle income, UM=upper-middle income, H=high income. World Bank. The world by income and region; 2022. [cited 2024 July 2]. Available from:
https://datatopics.worldbank.org/world-development-indicators/the-world-by-income-and-region.html
<sup>‡</sup>World Bank. GDP per capita (current US \$); [cited 2024 July 2].
Available from:
https://data.worldbank.org/indicator/NY.GDP.PCAP.CD
Abbreviations: GDP=gross domestic product; WHO=World Health Organization

Table 2 reports the study characteristics summary, while the Appendix describes the details of each individual study (supplemental tables S1 & S2). Health effects were reported using mortality-based outcomes (i.e., measures incorporating deaths) in the majority of studies: as DALY averted in 17 (30%) studies, QALY gained in six (11%) studies, and deaths in six (11%); whereas 12 (21%) studies used other health outcomes. The comparators used in most studies were either no fortification or pre-fortification. All except three studies were economic models, including 20 cost–benefit analyses (CBA). There was substantial heterogeneity among the studies due to varying interventions (i.e., a variety of nutrients and foods), industry costs (not shown), and modeling parameters (e.g., health outcome measures, time horizons). The 56 studies employed a variety of methodological approaches, such as decision trees and Markov models. Economic analytical perspectives also varied, including societal and healthcare system perspectives (yet most were not explicitly reported), with different costing methods utilized, for example, some studies focused on direct healthcare costs, while others encompassed broader economic impacts.

**Table 2.** Study characteristics summary.

| Characteristic | Studies (n)* | Percent |
| --- | --- | --- |
| **ECONOMIC EVALUATION TYPE** |  |  |
| Model (n=53), as follows: |  |  |
| CEA model | 27 | 48% |
| Cost–utility (CUA) model | 6 | 11% |
| CBA model | 14 | 25% |
| Both CEA & CBA models | 6 | 11% |
| Primary study | 3 | 5% |
| TOTAL | 56 | 100% |
| **HEALTH OUTCOME** |  |  |
| DALY averted<sup>†</sup> | 17 | 30% |
| QALY gained | 6 | 11% |
| Death | 6 | 11% |
| Adequate intake or Deficiency level | 4 | 7% |
| Anemia | 3 | 5% |
| Neural tube defect | 2 | 4% |
| Encephalopathy | 1 | 2% |
| Fracture | 1 | 2% |
| Goiter | 1 | 2% |
| Monetized health benefits | 15 | 27% |
| TOTAL | 56 | 100% |
| **COMPARATOR** |  |  |
| No/Pre-fortification | 34 | 61% |
| Unfortified food | 18 | 32% |
| Voluntary fortification | 4 | 7% |
| TOTAL | 56 | 100% |
| **PERSPECTIVE<sup>‡</sup>** |  |  |
| Societal (or social) perspective | 20 | 36% |
| Healthcare (or public health) perspective | 3 | 5% |
| Not reported | 33 | 59% |
| TOTAL | 56 | 100% |
| **COUNTRY ECONOMY<sup>§</sup>** |  |  |
| LMIC | 42 | 75% |
| High-income | 18 | 32% |
| Combined (multiple) incomes | 2 | 4% |
| TOTAL | 62 | 111% |
| **FUNDING SOURCE** |  |  |
| Public/Non-profit | 28 | 50% |
| Private/Industry | 2 | 4% |
| Mixed public/private | 5 | 9% |
| Unclear | 2 | 4% |
| No funding received | 6 | 11% |
| Not reported | 13 | 23% |
| TOTAL | 56 | 100% |
| **YEAR OF PUBLICATION** |  |  |
| 2020–2023 | 13 | 23% |
| 2010–2019 | 25 | 45% |
| 2000–2009 | 14 | 25% |
| 1990–1999 | 3 | 5% |
| 1980–1989 | 1 | 2% |
| TOTAL | 56 | 100% |

\*Number of studies may sum to >56 as some studies reported on >1 characteristic.
† Also known as Healthy life year gained.
‡The point of view of an economic study: a societal perspective considers costs and benefits for society; and a healthcare perspective includes costs and effects on the health sector.
§World Bank. The world by income and region; 2022 [cited 2024 July 2]. Available from:
https://datatopics.worldbank.org/world-development-indicators/the-world-by-income-and-region.html

Foods were fortified with 17 micronutrients, either alone or in combination with other nutrients (table 3). The single nutrients most frequently included in the studies were vitamin A, folic acid, iron, and iodine. Table 4 outlines the food vehicles assessed. Cereal grains/products (predominantly wheat flour) and condiments, such as edible oils, sugar, and salt, were the most common foods. Supplemental table S2 provides intervention details for each study.

**Table 3.** Micronutrients that foods were fortified with in the studies.

| Micronutrient | Fortified with single nutrient |  | Co-fortified with multiple nutrients |  |
| --- | --- | --- | --- | --- |
|  | Studies (n) | Number of Evaluations* | Studies (n) | Number of Evaluations* |
| Calcium | 1 | 1 | 5 | 15 |
| Copper | 0 | 0 | 1 | 1 |
| Folic acid | 12 | 22 | 14 | 147 |
| Iodine | 6 | 8 | 2 | 3 |
| Iron | 7 | 11 | 16 | 154 |
| Magnesium | 0 | 0 | 1 | 1 |
| Potassium | 1 | 1 | 0 | 0 |
| Vitamin A | 17 | 112 | 9 | 75 |
| Vitamin B1 | 1 | 2 | 8 | 69 |
| Vitamin B2 | 0 | 0 | 7 | 69 |
| Vitamin B3 | 0 | 0 | 8 | 68 |
| Vitamin B6 | 0 | 0 | 4 | 61 |
| Vitamin B12 | 0 | 0 | 7 | 118 |
| Vitamin C | 0 | 0 | 2 | 4 |
| Vitamin D | 1 | 4 | 2 | 2 |
| Vitamin E | 0 | 0 | 1 | 1 |
| Zinc | 4 | 7 | 7 | 70 |
| TOTALS | 50<sup>†</sup> | 168 | — | — |

\*From all outcomes, including CEAs, CUAs, and CBAs. Many studies reported relevant data on multiple evaluations (analyses) since they may have evaluated different interventions, countries, etc.
<sup>†</sup>Sums to <56 as some studies reported only on multi-ingredient combinations.

**Table 4.** Foods fortified in the studies.

| Food vehicles | Studies*<br>(n) | Number of<br>Evaluations<sup>†</sup> |
| --- | --- | --- |
| Cereal grains (n=39), reported as follows: |  |  |
| Wheat flour | 22 | 137 |
| Maize flour/meal | 6 | 35 |
| Flour | 6 | 9 |
| Rice | 3 | 11 |
| Cereal flours | 1 | 3 |
| Wheat | 1 | 4 |
| Products made with cereal grains (n=6), as follows: |  |  |
| Bread | 3 | 4 |
| Grain products | 2 | 4 |
| Biscuit | 1 | 1 |
| Infant cereals | 3 | 3 |
| Condiments (n=32), reported as follows: |  |  |
| Oils | 12 | 63 |
| Sugar | 7 | 54 |
| Salt | 7 | 8 |
| Soy sauce | 3 | 3 |
| Bouillon cubes | 1 | 3 |
| Fish sauce | 1 | 2 |
| Monosodium glutamate (MSG) | 1 | 2 |
| Milk powder/product | 2 | 2 |
| Combined several food types<sup>‡</sup> | 1 | 1 |
| TOTAL | 83 | — |

\*Sums to >56 as some studies reported on >1 food.
<sup>†</sup>From all outcomes. Many studies reported relevant data on multiple evaluations (analyses) since they may have evaluated different interventions, countries, etc.
<sup>‡</sup>Milk products, soy-/cereal-based drinks, fat spreads, bread, and orange juice combined.
Foods are reported as described in the studies, and where information was inadequate, foods may be misclassified.

An extensive supplemental table S2 reports full CE results from each study. Overall, across the many diverse economic models, ICERs measured as cost per DALY averted (or per healthy life year gained) were reported in 232 evaluations (analyses) in total (227 of which were from LMICs). Figure 3 displays the number of evaluations in a given range of ICER result values, after all were converted to 2022 US$. In the ICER category “<$150 per DALY averted” (selected as an arbitrary point of reference), there were 135 (58%) evaluations; and 194 (84%) evaluations in total had ICERs less than $1,000 per DALY averted. Eleven (5%) evaluations had ICERs higher than $10,000 per DALY averted; two of these were from lower-middle income, five from upper-middle income, and four from high-income countries.

**Figure 3.**
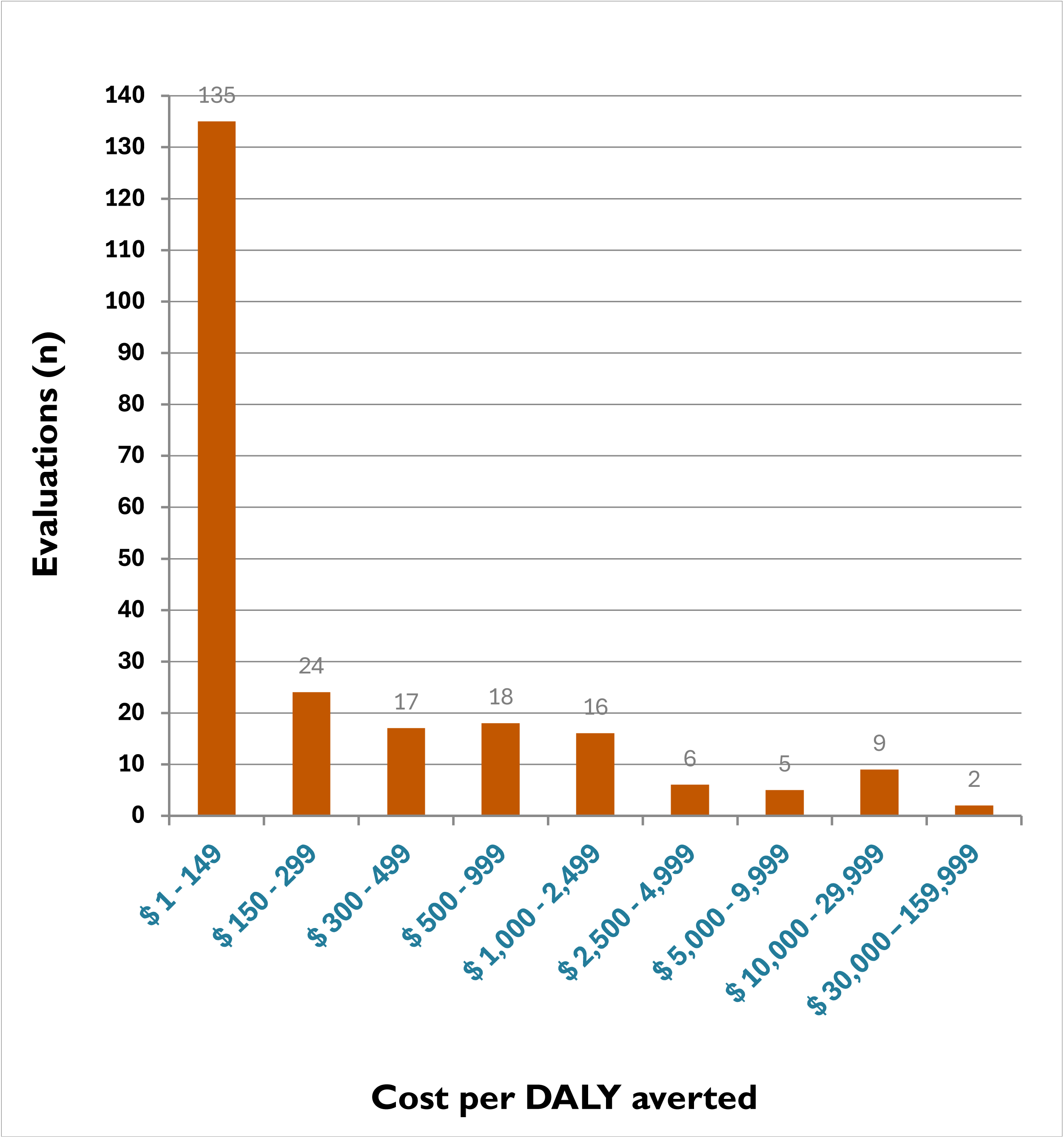
Overall cost-effectiveness results of fortification. Each bar shows the number of evaluations found within each ICER range (out of 232 total). ICER categories are expressed as 2022 US$ per DALY averted. Many studies reported relevant data on multiple evaluations (analyses) since they may have evaluated different interventions, countries, etc.

Regarding “hypothetical” cost-effectiveness threshold illustrations, results are presented in the Appendix (supplemental table S3), both overall and categorized by economy income. Overall, 87% (201/232) of ICERs (i.e., evaluations measured as cost per DALY averted) fell within 50% of GDP pc per country. Among LMICs and using as an example 35% of GDP pc, 84% (190 out of 227 evaluations) of ICERs were estimated to be cost-effective. Furthermore, focusing on low-income economy countries and using a lower example CE threshold of 20% of GDP per capita estimated that 71% (37/52) of ICERs were cost-effective.

Table 5 provides a matrix summary of ICER (DALY) results for each micronutrient alone and combination of micronutrients evaluated. Vitamin A had the most data with 90 evaluations (mainly due to the Fiedler & Macdonald, 2009 study(48)), of which 62 (69%) ICERs were less than $500 per DALY averted.

**Table 5.**
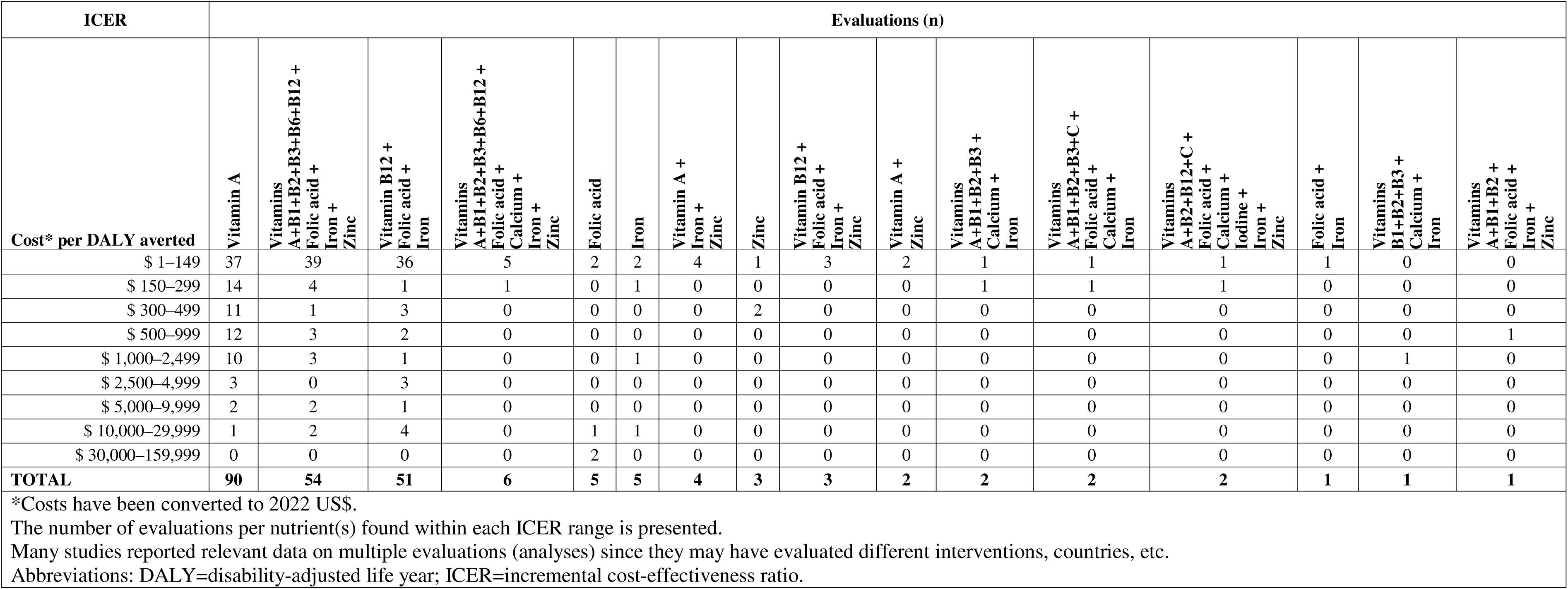
Fortification cost-effectiveness results per micronutrient & their combinations (ordered by total number of evaluations)

| ICER | Evaluations (n) |  |  |  |  |  |  |  |  |  |  |  |  |  |  |  |
| --- | --- | --- | --- | --- | --- | --- | --- | --- | --- | --- | --- | --- | --- | --- | --- | --- |
| Cost* per DALY averted | Vitamin A | Vitamins A+B1+B2+B3+B6+B12 + Folic acid + Iron + Zinc | Vitamin B12 + Folic acid + Iron | Vitamins A+B1+B2+B3+B6+B12 + Folic acid + Calcium + Iron + Zinc | Folic acid | Iron | Vitamin A + Iron + Zinc | Zinc | Vitamin B12 + Folic acid + Iron + Zinc | Vitamin A + Zinc | Vitamins A+B1+B2+B3 + Calcium + Iron | Vitamins A+B1+B2+B3+C + Folic acid + Calcium + Iron | Vitamins A+B2+B12+C + Folic acid + Calcium + Iodine + Iron + Zinc | Folic acid + Iron | Vitamins B1+B2+B3 + Calcium + Iron | Vitamins A+B1+B2 + Folic acid + Iron + Zinc |
| \$ 1–149 | 37 | 39 | 36 | 5 | 2 | 2 | 4 | 1 | 3 | 2 | 1 | 1 | 1 | 1 | 0 | 0 |
| \$ 150–299 | 14 | 4 | 1 | 1 | 0 | 1 | 0 | 0 | 0 | 0 | 1 | 1 | 1 | 0 | 0 | 0 |
| \$ 300–499 | 11 | 1 | 3 | 0 | 0 | 0 | 0 | 2 | 0 | 0 | 0 | 0 | 0 | 0 | 0 | 0 |
| \$ 500–999 | 12 | 3 | 2 | 0 | 0 | 0 | 0 | 0 | 0 | 0 | 0 | 0 | 0 | 0 | 0 | 1 |
| \$ 1,000–2,499 | 10 | 3 | 1 | 0 | 0 | 1 | 0 | 0 | 0 | 0 | 0 | 0 | 0 | 0 | 1 | 0 |
| \$ 2,500–4,999 | 3 | 0 | 3 | 0 | 0 | 0 | 0 | 0 | 0 | 0 | 0 | 0 | 0 | 0 | 0 | 0 |
| \$ 5,000–9,999 | 2 | 2 | 1 | 0 | 0 | 0 | 0 | 0 | 0 | 0 | 0 | 0 | 0 | 0 | 0 | 0 |
| \$ 10,000–29,999 | 1 | 2 | 4 | 0 | 1 | 1 | 0 | 0 | 0 | 0 | 0 | 0 | 0 | 0 | 0 | 0 |
| \$ 30,000–159,999 | 0 | 0 | 0 | 0 | 2 | 0 | 0 | 0 | 0 | 0 | 0 | 0 | 0 | 0 | 0 | 0 |
| **TOTAL** | **90** | **54** | **51** | **6** | **5** | **5** | **4** | **3** | **3** | **2** | **2** | **2** | **2** | **1** | **1** | **1** |
| *Costs have been converted to 2022 US\$. | | | | | | | | | | | | | | | | |
| The number of evaluations per nutrient(s) found within each ICER range is presented. |  |  |  |  |  |  |  |  |  |  |  |  |  |  |  |  |
| Many studies reported relevant data on multiple evaluations (analyses) since they may have evaluated different interventions, countries, etc. |  |  |  |  |  |  |  |  |  |  |  |  |  |  |  |  |
| Abbreviations: DALY=disability-adjusted life year; ICER=incremental cost-effectiveness ratio. |  |  |  |  |  |  |  |  |  |  |  |  |  |  |  |  |

Among the six cost-utility studies reporting QALYs gained,(9, 34, 37, 57, 77, 79) there were eight evaluations in total (seven of which were from high-income countries; supplemental table S2). Six of these eight were dominant (i.e., fortification was less costly and more effective than comparator); while the other two ICERs were ~$7,000 and ~$13,000 per QALY gained (from an upper-middle income and high-income country, respectively). Furthermore, if both DALY and QALY ICERs are combined for a hypothetical CE threshold example, 87% (208/240) overall fall below 50% of GDP per capita per country, which is similar to the results above. Additionally, from six studies on mortality alone,(30, 65–67, 78, 88) there were 18 total evaluations, five of which were dominant. Across the other ICERs, nine were <$1,000 and another two were <$2,500 per death averted. The other two ICERs were ~$9,000 and ~$62,000 per death averted (both from the same study in a lower-middle income country) (supplemental table S2).

CBAs presented a total of 47 unique evaluations of monetized benefit–cost ratios (BCRs), which report comparisons of relative costs and benefits valued in monetary terms, with B/C ratio values >1 indicating that the calculated benefits outweigh the costs.(8, 29, 30, 33, 40, 49, 50, 55, 58, 61, 63, 68, 71, 74, 76, 78, 84, 85, 92) All BCR estimates exceeded one, with a range of 1·50:1 to 100·6:1. Moreover, 16 BCRs estimated benefits valued at ten times or more of the costs invested on food fortification. Table 6 reports the BCRs categorized, and supplemental table S2 provides details of these evaluations.

**Table 6.** Cost-benefit analyses summary results of food fortification.

| **Benefit-cost ratio**<br>**(BCR) range\*** | **Evaluations**<br>**(n)** |
| --- | --- |
| 1.5–4.9 (to :1) | 15 |
| 5.0–9.9 (to :1) | 16 |
| 10.0–19.9 (to :1) | 10 |
| 20.0–29.9 (to :1) | 0 |
| 30.0–39.9 (to :1) | 3 |
| 40.0–49.9 (to :1) | 1 |
| 50.0–59.9 (to :1) | 0 |
| 60.0–69.9 (to :1) | 0 |
| 70.0–79.9 (to :1) | 0 |
| 80.0–89.9 (to :1) | 1 |
| 90.0–100.6 (to :1) | 1 |
| **TOTAL** | **47** |

\*Each category groups a range of reported values for a benefit-cost ratio (compared to 1).
Many studies reported relevant data on multiple evaluations since they may have evaluated different interventions, countries, etc.

The quality of reporting of the studies had many deficiencies. The Appendix provides detailed quality appraisals of each individual study (supplemental tables S4 & S5). Figure 4 displays the quality appraisal results from the Philips’ modeling framework for each of the three key modeling dimensions assessed; a “Yes” response indicates there was sufficient information reported for each question. Overall across the 53 modeling studies, the majority of the 21 items about ‘structure of the economic model’ scored positively with a median of 80% (interquartile range, IQR: 67–89%) judged with “Yes” responses (indicating that the “median study” scored a “Yes” response on 80% of the 21 questions assessing ‘structure’). However, items about model ‘data’ and ‘consistency’ scored poorly with medians of 52% (IQR: 35–65%) and 50% (IQR: 25– 60%), respectively. The main elements regarding ‘data used in the models’ that were poorly reported were related to uncertainty/sensitivity analyses and half cycle correction; and ‘consistency’ items that scored lowest included calibration and testing of the mathematical logic. Furthermore, comparators used in the models were often not clearly described. The three primary studies assessed using the CHEC list tool had several deficiencies, including limited time horizons, analytical perspectives, and sensitivity analyses.

**Figure 4.**
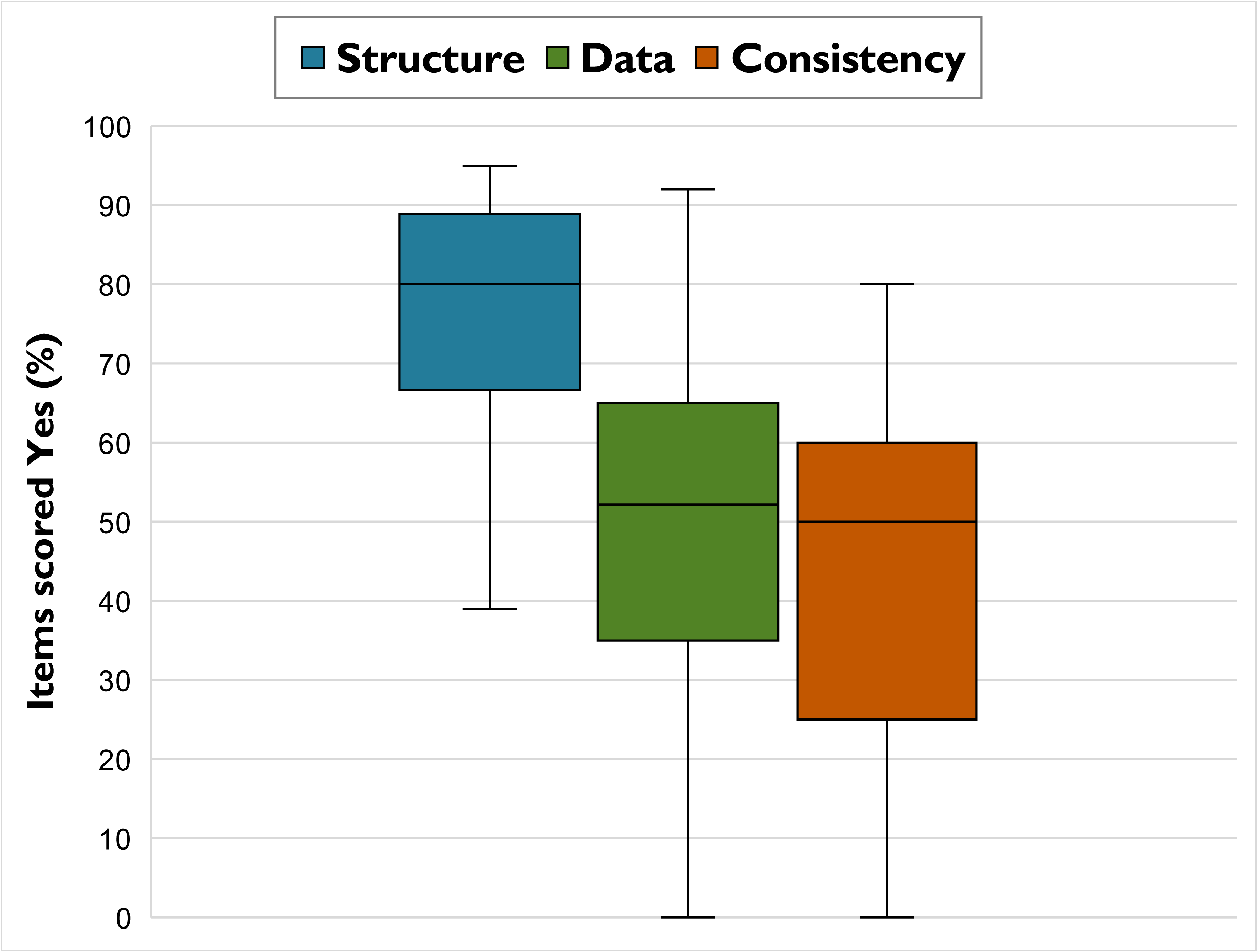
Quality appraisal of the economic models of food fortification. Philips tool’s^23^ key dimensions assessed are: ‘Structure’ of the model (blue), ‘Data’ used (green), and ‘Consistency’ of the model (orange). Legend: middle line = median; box length = interquartile range; whiskers = range.

Since a large amount of data was included from one study, i.e., Fiedler & Macdonald (2009),(48) a sensitivity analysis was conducted without its 171 evaluations. The results were consistent with those presented above from the remaining 61 evaluations in 16 other studies: 40 (66%) evaluations had ICERs less than $150 per DALY averted; and 53 (87%) evaluations had ICERs <$1,000 per DALY averted.(32, 35, 38, 41–43, 45–47, 54, 60, 63, 64, 69, 82, 91) Four (7%) evaluations had ICERs higher than $10,000 per DALY averted and these were all from high-income countries.

## DISCUSSION

Our comprehensive systematic review of 55 economic studies reporting over 200 analyses and spanning 63 countries, including >40 LMICs, found the majority of results suggest that food fortification programs are cost-effective. We found 84% of evaluations had ICERs <$1,000 per DALY averted, and 58% were <$150 per DALY averted (all converted to 2022 US$). Six out of eight cost–utility (QALY) ICERs were dominant, meaning fortification was less costly and more effective than the comparator. A total of 47 BCRs reported were positive (range 1·50:1 to 100·6:1), indicating the benefits of LSFF outweighed costs.

Our illustrations using “hypothetical” cost-effectiveness thresholds found that: overall, 87% of (DALY) ICERs fell within 50% of GDP pc per country; for LMICs and using 35% of GDP pc, 84% of ICERs were lower; and among low-income economy countries and using 20% of GDP pc, 71% were estimated to be cost-effective. Regarding the additional presentation of these CE threshold illustrations, they are intended simply as examples since it is important to highlight, as noted above, that judging local cost-effectiveness amid resource allocation needs to be determined by each country and guidance recommends against using generic cost-effectiveness thresholds as rules for decision-making.(22) Furthermore, we fully acknowledge that these example threshold illustrations are not meant to apply to all countries and contexts and they may not be ideal. Recognizing these limitations, we selected this method and these percentages of GDP pc per country based on recent research of commonly used examples.(26–28)

Significant heterogeneity existed between studies due to a wide variety of interventions, model parameters, and national contexts, such as differing malnutrition burdens, health systems, government subsidies, and diets. This methodological diversity reflects the complexity and context-specific nature of economic evaluation of LSFF. Therefore, meta-analysis was not appropriate, as is the case in most economic reviews of a broad and international nature. Additionally, depending on the CE threshold used, roughly 15-30% of the DALY ICERs were estimated to not be cost-effective, which may reflect differing methods and contexts among other factors. Quality appraisal identified many deficiencies in the quality of reporting of the studies, which affects the strength of the evidence. In particular, the lowest scoring components from Philips’ modeling framework(23) included items relating to uncertainty/sensitivity analyses, half cycle correction/cycle length, model calibration, and testing of the mathematical logic.

This large systematic review of LSFF provides a substantially broader synthesis of the economic evidence base than has previously been reported, filling a significant research gap. Importantly, the breadth of countries included and income economies represented augments the international literature on a system-level intervention for reducing global malnutrition, mortality, and morbidity. Other reviews have studied cost-effectiveness but their scope focused on specific foods or nutrients. Several economics systematic reviews examined fortification with folic acid, comprising up to 13 relevant studies each, with positive results. Rodrigues et al. (2021) found fortification cost-benefit analyses yielded a median ratio of 17.5:1 (range 0·98:1 to 417·1:1).(11) A second systematic review examined mandatory folic acid fortification in Australia and concluded the initiative was health producing and cost saving.(9) Yi et al. (2011) concluded that fortification with folic acid is a cost-effective way to reduce neural tube birth defects.(13) Another review of folic acid fortification noted that none of the included studies evaluated the economic impact in LMICs.(12) Finally, a scoping review including three cost-effectiveness studies in Europe on vitamin D-fortified dairy products for elderly fracture prevention reported that results suggest the intervention is generally cost-effective.(10)

This systematic review has several strengths. We employed rigorous systematic review methodology and reporting guidance, including *a priori* protocol registration, comprehensive literature searching, study quality appraisal, and consistent and transparent cost conversion methods. The main limitation at the review-level was that meta-analysis was not feasible as the economic models were very heterogeneous. This impacts the generalizability of the findings and caution should be applied in drawing conclusions across different contexts. The main study-level limitation of the results was the quality assessment of model reporting which found many deficiencies. In addition, it is relevant to highlight that economic modeling involves a multitude of assumptions and estimates, so results should be interpreted more generally as orders of magnitude rather than as definitive point estimates. Future cost-effectiveness studies of LSFF should endeavor to report more complete details underlying their models and methods, and they may benefit from presenting these in an appendix when journal space limitations are a factor. We also identified some research gaps as sparse economic data were available for certain beneficial nutrients, such as vitamin D. Future research syntheses on ‘head-to-head’ comparisons of fortification interventions (i.e., comparing one to another) could provide additional data for program prioritization.

The decision by policymakers to enact or strengthen LSFF programs in their countries is predicated by many factors, including costs and cost-effectiveness.(93) Nutrition for Growth aims to mobilize governments, bilateral agencies, private investors, businesses, civil society, donors, and others to increase and sustain their funding for nutrition actions, especially those that are evidence-based.(94) Through the *Global Nutrition Report,* such financial commitments are being tracked internationally.(95) The evidence base is provided by LSFF’s efficacy and effectiveness.(7, 96) While cost-effectiveness has been studied in limited scenarios, our systematic review is significantly broader in geographic, fortification vehicle, and nutrient scope. The expectation is that this more robust evidence base will encourage greater investment in initiating and strengthening fortification programs.

This comprehensive systematic review covering a broad scope of economic evaluations found that food fortification programs are likely cost-effective in most contexts. These findings may assist with evidence-informed decision-making for global health policy, particularly in resource-constrained countries.

## Data Availability

This research, including an outcomes data spreadsheet, will be submitted for peer-reviewed journal publication. All data produced in the present study are available upon reasonable request to the authors (Nicholas Henschke).

## Contributors

BLT and HP conceptualized the project. EC, FP, HP, MC, GV, NH, and BLT developed the protocol and study design. FP, EC, GV, and NH contributed to the study’s methodology. EC searched the literature. EC, FP, BB, and MC were involved in eligibility screening, data collection and assessment, and verified the data. EC and FP conducted data analysis. FP performed cost conversions. EC, FP, HP, BB, MC, GV, MW, NH, and BLT contributed to data interpretation. NH, HP, and BLT supervised the study. All authors assisted with writing and revising the manuscript.

## Acknowledgments

Funding: This work was supported by the U.S. Agency for International Development (USAID; cooperative agreement number 7200AA22LE00002; USAID Advancing Food Fortification Opportunities to Reinforce Diets). The contents are the responsibility of TechnoServe and do not necessarily reflect the views of USAID or the U.S. Government.

We are grateful for the valuable assistance provided by: Brent Wibberley (TechnoServe and USAID AFFORD); Dominic Schofield (TechnoServe); Katherine Adams (University of California Davis, Institute for Global Nutrition); Kylee Rappaport (Emory University, Rollins School of Public Health); Heather McIntosh and Jennifer Petkovic (Cochrane Response); and Courtney Meyer (USAID AFFORD). We also wish to thank study authors who responded to our queries.

