## Supplemental Appendix for "Cost-effectiveness of food fortification for reducing global malnutrition: a systematic review of economic evaluations across 63 countries"

### Database Search Strategies

*Ovid: Embase Classic+Embase 1947 to 2024 January 17; MEDLINE(R) ALL 1946 to January 17, 2024; EBM Reviews - Cochrane Central Register of Controlled Trials December 2023*

1. Food, Fortified/
2. (food* adj8 (fortif* or enrich*)).mp.
3. ((grain or grains or cereal* or flour or flours or salt or sauce or sauces or milk or milks or bread or breads or sugar or beverage* or juice or juices or oil or oils or maize or corn or wheat or rice or condiment* or yog?ourt* or yog?urt* or micronutrient* or micro nutrient* or vitamin* or mineral* or postharvest* or post harvest*) adj6 (fortif* or enriched or enrichment)).mp.
4. (LSFF or SAPFF or salt iodization).mp.
5. or/1-4
6. limit 5 to ("economics (maximizes sensitivity)" or "costs (maximizes sensitivity)") [Ovid filters]
7. Cost-Benefit Analysis/ or Quality-Adjusted Life Years/ or Markov Chains/ or exp Models, Economic/
8. cost*.ti.
9. (cost* adj2 (utilit* or effective* or assess* or evaluat* or analys* or model* or benefit* or threshold* or quality or expens* or saving* or reduc*)).tw.
10. (economic* adj2 (evaluat* or assess* or analys* or model* or outcome* or benefit* or threshold* or expens* or saving* or reduc*)).tw.
11. ((qualit* or disabilit*) adj2 adjust* adj2 life*).tw.
12. (QALY* or DALY*).tw.
13. (incremental* adj2 cost*).tw.
14. (ICER or utilities or markov* or dollar* or USD or cents or pound or pounds or GBP or sterling* or pence or euro or euros or yen or JPY or Eq5D* or EQ-5D*).tw.
15. ((utility or effective*) adj2 analys*).tw.
16. (willing* adj2 pay*).tw.
17. ((euroqol or euro-qol or euroquol or euro-quol or eurocol or euro-col) adj3 ("5" or five)).tw.
18. (european* adj2 quality adj3 ("5" or five)).tw.
19. or/7-18 [Hubbard 2022 CUA filter]
20. 5 and 19
21. Economics/ or exp "Costs and Cost Analysis"/ or Economics, Nursing/ or Economics, Medical/ or Economics, Pharmaceutical/ or exp Economics, Hospital/ or Economics, Dental/ or exp "Fees and Charges"/ or exp Budgets/
22. budget*.ti,ab,kf.
23. (economic* or cost or costs or costly or costing or price or prices or pricing or pharmacoeconomic* or pharmaco-economic* or expenditure or expenditures or expense or expenses or financial or finance or finances or financed).ti,kf.
24. (economic* or cost or costs or costly or costing or price or prices or pricing or pharmacoeconomic* or pharmaco-economic* or expenditure or expenditures or expense or expenses or financial or finance or finances or financed).ab. /freq=2
25. (cost* adj2 (effective* or utilit* or benefit* or minimi* or analy* or outcome or outcomes)).ab,kf.
26. (value adj2 (money or monetary)).ti,ab,kf.
27. exp models, economic/ or markov chains/ or monte carlo method/ or exp Decision Theory/
28. economic model*.ab,kf.
29. (markov or monte carlo).ti,ab,kf.
30. (decision* adj2 (tree* or analy* or model*)).ti,ab,kf.
31. or/21-30 [CADTH 2023 Economic Evaluations & Models filter]
32. 5 and 31
33. "Value of Life"/ or Quality of Life/ or Quality-Adjusted Life Years/ or Disability-Adjusted Life Years/ or Healthy Life Expectancy/
34. quality of life.ti,kf.
35. ((instrument or instruments) adj3 quality of life).ab.
36. (quality adjusted life or qaly* or qald* or qale* or qtime* or life year or life years or disability adjusted life or daly* or disability free life expectanc* or haly* or health* life expectanc*).ti,ab,kf.
37. (sf36 or sf 36 or short form 36 or shortform 36 or short form36 or shortform36 or sf thirtysix or sfthirtysix or sfthirty six or sf thirty six or shortform thirtysix or shortform thirty six or short form thirtysix or short form thirty

six or sf6 or sf 6 or short form 6 or shortform 6 or sf six or sfsix or shortform six or short form six or shortform6 or short form6 or sf8 or sf 8 or sf eight or sfeight or shortform 8 or shortform 8 or shortform8 or short form8 or shortform eight or short form eight or sf12 or sf 12 or short form 12 or shortform 12 or short form12 or shortform12 or sf twelve or sftwelve or shortform twelve or short form twelve or sf16 or sf 16 or short form 16 or shortform 16 or short form16 or shortform16 or sf sixteen or sfsixteen or shortform sixteen or short form sixteen or sf20 or sf 20 or short form 20 or shortform 20 or short form20 or shortform20 or sf twenty or sftwenty or shortform twenty or short form twenty or hql or hqol or h qol or hrqol or hrqol or hye or hyes or pqol or qls or quality of wellbeing or quality of well being or index of wellbeing or index of well being or qwb or nottingham health profile* or sickness impact profile).ti,ab,kf.
38. (health* adj2 year* adj2 equivalent*).ti,ab,kf.
39. exp health status indicators/
40. (health adj3 (utilit* or status)).ti,ab,kf.
41. (utilit* adj3 (valu* or measur* or health or life or estimat* or elicit* or disease or score* or weight)).ti,ab,kf.
42. (preference* adj3 (valu* or measur* or health or life or estimat* or elicit* or disease or score* or instrument or instruments)).ti,ab,kf.
43. (disutilit* or rosser or willingness to pay or standard gamble* or time trade off or time tradeoff or tto or hui or hui1 or hui2 or hui3 or eq or euroqol or euro qol or eq5d or eq 5d or euroqual or euro qual or duke health profile or functional status questionnaire or dartmouth coop functional health assessment*).ti,ab,kf.
44. or/33-43 [CADTH 2023 Economic - Health Utilities / Quality of Life filter]
45. 5 and 44
46. 6 or 20 or 32 or 45
47. exp animals/ not humans/
48. 46 not 47
49. (comment or editorial or newspaper article).pt.
50. 48 not 49
51. 50 use medall
52. 20 or 32 or 45
53. 52 use cctr
54. fortified food/
55. (food* adj8 (fortif* or enrich*)).mp.
56. ((grain or grains or cereal* or flour or flours or salt or sauce or sauces or milk or milks or bread or breads or sugar or beverage* or juice or juices or oil or oils or maize or corn or wheat or rice or condiment* or yog?ourt* or yog?urt* or micronutrient* or micro nutrient* or vitamin* or mineral* or postharvest* or post harvest*) adj6 (fortif* or enriched or enrichment)).mp.
57. (LSFF or SAPFF or salt iodization).mp.
58. or/54-57
59. limit 58 to "economics (maximizes sensitivity)"
60. cost*.ti.
61. (cost* adj2 (utilit* or effective* or assess* or evaluat* or analys* or model* or benefit* or threshold* or quality or expens* or saving* or reduc*)).tw.
62. (economic* adj2 (evaluat* or assess* or analys* or model* or outcome* or benefit* or threshold* or expens* or saving* or reduc*)).tw.
63. ((qualit* or disabilit*) adj2 adjust* adj2 life*).tw.
64. (QALY* or DALY*).tw.
65. (incremental* adj2 cost*).tw.
66. (ICER or utilities or markov* or dollar* or USD or cents or pound or pounds or GBP or sterling* or pence or euro or euros or yen or JPY or Eq5D* or EQ-5D*).tw.
67. ((utility or effective*) adj2 analys*).tw.
68. (willing* adj2 pay*).tw.
69. ((euroqol or euro-qol or euroquol or euro-quol or eurocol or euro-col) adj3 ("5" or five)).tw.
70. (european* adj2 quality adj3 ("5" or five)).tw.
71. budget*.ti,ab,kf.
72. (economic* or cost or costs or costly or costing or price or prices or pricing or pharmacoeconomic* or pharmaco-economic* or expenditure or expenditures or expense or expenses or financial or finance or finances or financed).ti,kf.

73. (economic* or cost or costs or costly or costing or price or prices or pricing or pharmacoeconomic* or pharmaco-economic* or expenditure or expenditures or expense or expenses or financial or finance or finances or financed).ab. /freq=2
74. (cost* adj2 (effective* or utilit* or benefit* or minimi* or analy* or outcome or outcomes)).ab,kf.
75. (value adj2 (money or monetary)).ti,ab,kf.
76. economic model*.ab,kf.
77. (markov or monte carlo).ti,ab,kf.
78. (decision* adj2 (tree* or analy* or model*)).ti,ab,kf.
79. quality of life.ti,kf.
80. ((instrument or instruments) adj3 quality of life).ab.
81. (quality adjusted life or qaly* or qald* or qale* or qtime* or life year or life years or disability adjusted life or daly* or disability free life expectanc* or haly* or health* life expectanc*).ti,ab,kf.
82. (sf36 or sf 36 or short form 36 or shortform 36 or short form36 or shortform36 or sf thirtysix or sfthirtysix or sfthirty six or sf thirty six or shortform thirtysix or shortform thirty six or short form thirtysix or short form thirty six or sf6 or sf 6 or short form 6 or shortform 6 or sf six or sfsix or shortform six or short form six or shortform6 or short form6 or sf8 or sf 8 or sf eight or sfeight or shortform 8 or shortform 8 or shortform8 or short form8 or shortform eight or short form eight or sf12 or sf 12 or short form 12 or shortform 12 or short form12 or shortform12 or sf twelve or sftwelve or shortform twelve or short form twelve or sf16 or sf 16 or short form 16 or shortform 16 or short form16 or shortform16 or sf sixteen or sfsixteen or shortform sixteen or short form sixteen or sf20 or sf 20 or short form 20 or shortform 20 or short form20 or shortform20 or sf twenty or sftwenty or shortform twenty or short form twenty or hql or hqol or h qol or hrqol or hrqol or hye or hyes or pqol or qls or quality of wellbeing or quality of well being or index of wellbeing or index of well being or qwb or nottingham health profile* or sickness impact profile).ti,ab,kf.
83. (health* adj2 year* adj2 equivalent*).ti,ab,kf.
84. (health adj3 (utilit* or status)).ti,ab,kf.
85. (utilit* adj3 (valu* or measur* or health or life or estimat* or elicit* or disease or score* or weight)).ti,ab,kf.
86. (preference* adj3 (valu* or measur* or health or life or estimat* or elicit* or disease or score* or instrument or instruments)).ti,ab,kf.
87. (disutilit* or rosser or willingness to pay or standard gamble* or time trade off or time tradeoff or tto or hui or hui1 or hui2 or hui3 or eq or euroqol or euro qol or eq5d or eq 5d or euroqual or euro qual or duke health profile or functional status questionnaire or dartmouth coop functional health assessment*).ti,ab,kf.
88. cost utility analysis/ or quality adjusted life year/
89. or/60-70,88
90. 58 and 89
91. Economics/ or Cost/ or exp Health Economics/ or Budget/
92. Statistical Model/ or exp economic model/ or Probability/ or monte carlo method/ or Decision Theory/ or Decision Tree/
93. or/71-78,91-92
94. 58 and 93
95. socioeconomics/ or exp Quality of Life/ or Quality-Adjusted Life Year/ or disability-adjusted life year/ or healthy life expectancy/
96. exp Short form 36/
97. exp assessment of humans/
98. health status indicator/ or Willingness To Pay/ or Standard Gamble/ or time trade-off method/
99. or/79-87,95-98
100. 58 and 99
101. 59 or 90 or 94 or 100
102. (exp animal/ or exp animal experimentation/ or exp animal model/ or exp animal experiment/ or nonhuman/ or exp vertebrate/) not (exp human/ or exp human experimentation/ or exp human experiment/)
103. 101 not 102
104. editorial.pt.
105. 103 not 104
106. 105 use emczd
107. 51 or 53 or 106
108. limit 107 to yr="1946 - 2010"

109. limit 107 to yr="2011 -Current"
110. 108 or 109
111. 107 not 110
112. remove duplicates from 108
113. remove duplicates from 109
114. 111 or 112 or 113

Economics search filters included the following:

Economic Evaluations & Models – MEDLINE & Embase. In: CADTH Search Filters Database. Ottawa: CADTH; 2023: https://searchfilters.cadth.ca/link/16

Economic - Health Utilities / Quality of Life - MEDLINE. In: CADTH Search Filters Database. Ottawa: CADTH; 2023: https://searchfilters.cadth.ca/link/19

Hubbard W, Walsh N, Hudson T, Heath A, Dietz J, Rogers G. Development and validation of paired MEDLINE and Embase search filters for cost-utility studies. BMC Med Res Methodol. 2022 Dec 3;22(1):310.

Ovid. Economics & Costs search filters (maximizes sensitivity) for MEDLINE and Embase databases. Accessed August 31, 2023.

### List of Included Studies' IDs and Bibliography

| Study ID (Author Year) | Bibliography |
|---|---|
| Access Economics 2006 | Access Economics. Cost benefit analysis of fortifying the food supply with folic acid. Food Standards Australia New Zealand (FSANZ); 2006. |
| Adams 2022 | Adams KP, Luo H, Vosti SA, et al. Comparing estimated cost-effectiveness of micronutrient intervention programs using primary and secondary data: evidence from Cameroon. Annals of the New York Academy of Sciences 2022; 1510(1): 100-20. https://doi.org/10.1111/nyas.14726 |
| Asian Develop. Bank 2004 | Asian Development Bank. 2004 June. Food Fortification in Asia: Improving Health and Building Economies. Manila, Philippines. |
| Australian HMAC 2017 | Australian Health Ministers' Advisory Council (HMAC), 2017, The effectiveness and cost-effectiveness of mandatory folic acid and iodine fortification. |
| Baltussen 2004 | Baltussen R, Knai C, Sharan M. Iron fortification and iron supplementation are cost-effective interventions to reduce iron deficiency in four subregions of the world. The Journal of nutrition 2004; 134(10): 2678-84. |
| Bentley 2009 | Bentley TG, Weinstein MC, Willett WC, Kuntz KM. A cost-effectiveness analysis of folic acid fortification policy in the United States. Public health nutrition 2009; 12(4): 455-67. |
| Chow 2010 | Chow J, Klein EY, Laxminarayan R. Cost-effectiveness of "golden mustard" for treating vitamin A deficiency in India. PloS one 2010; 5(8): e12046. |
| Connelly 1996 | Connelly L, Price J. Preventing the Wernicke-Korsakoff syndrome in Australia: cost-effectiveness of thiamin-supplementation alternatives. Australian and New Zealand journal of public health 1996; 20(2): 181-7. |
| Dainelli 2017 | Dainelli L, Xu T, Li M, et al. Cost-effectiveness of milk powder fortified with potassium to decrease blood pressure and prevent cardiovascular events among the adult population in China: a Markov model. BMJ open 2017; 7(9): e017136. |
| Dalziel 2010 | Dalziel K, Segal L, Katz R. Cost-effectiveness of mandatory folate fortification v. other options for the prevention of neural tube defects: results from Australia and New Zealand. Public health nutrition 2010; 13(4): 566-78. |
| Detzel 2016 | Detzel P. Estimating the contribution on the reduction of the burden of iron deficiency anemia from the current fortification strategy of commercial foods targeted to infants and toddlers in india. Scenarios using systematic review and real world evidence. Value in Health 2016; 19(7): A834. |
| Edejer 2005 | Edejer TT-T, Aikins M, Black R, Wolfson L, Hutubessy R, Evans DB. Cost effectiveness analysis of strategies for child health in developing countries. BMJ (Clinical research ed) 2005; 331(7526): 1177. |
| Fiedler 2000 | Fiedler JL, Dado DR, Maglalang H, Juban N, Capistrano M, Magpantay MV. Cost analysis as a vitamin A program design and evaluation tool: a case study of the Philippines. Social science & medicine (1982) 2000; 51(2): 223-42. |
| Fiedler 2009 | Fiedler JL, Macdonald B. A strategic approach to the unfinished fortification agenda: feasibility, costs, and cost-effectiveness analysis of fortification programs in 48 countries. Food and nutrition bulletin 2009; 30(4): 283-316. https://doi.org/10.1177/156482650903000401 |
| Fiedler 2010 | Fiedler JL, Afidra R. Vitamin A fortification in Uganda: comparing the feasibility, coverage, costs, and cost-effectiveness of fortifying vegetable oil and sugar. Food and nutrition bulletin 2010; 31(2): 193-205. |
| Fiedler 2012 | Fiedler JL, Babu S, Smitz M-F, Lividini K, Bermudez O. Indian social safety net programs as platforms for introducing wheat flour fortification: a case study of Gujarat, India. Food and nutrition bulletin 2012; 33(1): 11-30. |
| Fiedler 2013 | Fiedler JL, Lividini K, Kabaghe G, et al. Assessing Zambia's industrial fortification options: Getting beyond changes in prevalence and cost effectiveness. Food and Nutrition Bulletin 2013; 34(4): 501-19. |
| Fiedler 2014 | Fiedler JL, Lividini K. Managing the vitamin A program portfolio: a case study of Zambia, 2013-2042. Food and nutrition bulletin 2014; 35(1): 105-25. |
| Fiedler 2015 | Fiedler JL, Lividini K, Guyondet C, Bermudez OI. Assessing alternative industrial fortification portfolios: a Bangladesh case study. Food and nutrition bulletin 2015; 36(1): 57-74. |
| GAIN 2017 | Global Alliance for Improved Nutrition (GAIN). Report on Analysis of Economic Losses Due to Iron & Folic Acid Deficiencies in Afghanistan: Food Fortification as a Cost-Effective Stratgey for Economic Growth. United States Agency for International Development (USAID); 2017 June. |
| Ghauri 2015 | Ghauri K. Analysis of Economic Losses Due to Iron & Folic Acid Deficiencies in Pakistan: Food Fortification, a Cost-Effective Strategy to Reduce Losses. Global Alliance for Improved Nutrition (GAIN); 2015 July. |
| Ghauri 2016 | Ghauri K. Food Fortification in Tajikistan: A Cost-Effective Strategy for Sustainable Economic Growth. United States Agency for International Development (USAID); 2016 June. |
| Gorstein 2020 | Gorstein JL, Bagriansky J, Pearce EN, Kupka R, Zimmermann MB. Estimating the Health and Economic Benefits of Universal Salt Iodization Programs to Correct Iodine Deficiency Disorders. Thyroid : official journal of the American Thyroid Association 2020; 30(12): 1802-9. |
| Grosse 2005 | Grosse SD, Waitzman NJ, Romano PS, Mulinare J. Reevaluating the benefits of folic acid fortification in the United States: economic analysis, regulation, and public health. American journal of public health 2005; 95(11): 1917-22. |
| Grosse 2016 | Grosse SD, Berry RJ, Mick Tilford J, Kucik JE, Waitzman NJ. Retrospective Assessment of Cost Savings From Prevention: Folic Acid Fortification and Spina Bifida in the U.S. American journal of preventive medicine 2016; 50(5 Suppl 1): S74-S80. |
| Hoddinott 2018 | Hoddinott J. The investment case for folic acid fortification in developing countries. Annals of the New York Academy of Sciences 2018; 1414(1): 72-81. |
| Horton 2003 | Horton S, Ross J. The Economics of Iron Deficiency. Food Policy 2003; 28(1): 51-75. Corrigendum in: Food Policy 2007; 32(1): 141-143. |

| Study ID (Author Year) | Bibliography |
|---|---|
| Horton 2008 | Horton S, Alderman H, Rivera JA. Copenhagen Consensus 2008 Challenge Paper: Hunger and Malnutrition. Tewksbury, MA: Copenhagen Consensus Center; 2008 May. https://copenhagenconsensus.com/research-topic/food-security-nutrition |
| Huang 2020 | Huang Q, Liu C, Zhou L-A. Farewell to the God of Plague: Estimating the Effects of China's Universal Salt Iodization on Educational Outcomes. Journal of Comparative Economics 2020; 48(1): 20-36. |
| Jentink 2008 | Jentink J, van de Vrie-Hoekstra NW, de Jong-van den Berg LTW, Postma MJ. Economic evaluation of folic acid food fortification in The Netherlands. European journal of public health 2008; 18(3): 270-4. |
| Johnson 2021 | Johnson Q, Asfaw E. Technical report: Cost benefit analysis of oil and wheat flour fortification in Ethiopia. Global Alliance for Improved Nutrition; 2021. |
| Kagin 2015 | Kagin J, Vosti SA, Engle-Stone R, et al. Measuring the Costs of Vitamin A Interventions: Institutional, Spatial, and Temporal Issues in the Context of Cameroon. Food and nutrition bulletin 2015; 36(3 Suppl): S172-92. |
| Kakietek 2018 | Kakietek J, Provo A, Mehta M, Sharmin F, Shekar M. Supporting the National Action Plan on Nutrition: Estimating the Cost, Impact, Cost-Effectiveness, and Economic Benefits of Expanding the Coverage of Direct Nutrition Interventions in Bangladesh. Washington, DC: The World Bank; 2018 April. |
| Kancherla 2021 | Kancherla V, Chadha M, Rowe L, et al. Reducing the Burden of Anemia and Neural Tube Defects in Low- and Middle-Income Countries: An Analysis to Identify Countries with an Immediate Potential to Benefit from Large-Scale Mandatory Fortification of Wheat Flour and Rice. Nutrients 2021; 13(1). |
| Lara 2015 | Lara C, Detzel P. A cost-effectiveness study on the increased intake of potassium and vitamin B2 among adults in China. Value in Health 2015; 18(7): A394. |
| Llanos 2007 | Llanos A, Hertrampf E, Cortes F, Pardo A, Grosse SD, Uauy R. Cost-effectiveness of a folic acid fortification program in Chile. Health policy (Amsterdam, Netherlands) 2007; 83(2-3): 295-303. |
| Ma 2008 | Ma G, Jin Y, Li Y, et al. Iron and zinc deficiencies in China: what is a feasible and cost-effective strategy? Public health nutrition 2008; 11(6): 632-8. |
| Mardones-Santander 1991 | Mardones-Santander F, Rosso P, Zamora R, Mardones-Restat F, Gonzalez N, Uiterwaal D. Cost-effectiveness of a nutrition intervention program for pregnant women. Nutrition Research 1991; 11(4): 295-307. |
| Moges 2020 | Moges T, Tesfaye B, Vosti S, Engle-Stone R, Woldegebreal D, Luo H, Asfaw E, Kagin J. (2020). Fortifying oil with vitamin A and wheat flour with zinc: Predicted impact on nutritional outcomes, lives saved and economic benefits. Ethiopian Public Health Institute. |
| MQSUN 2014 | Maximising the Quality of Scaling up Nutrition Programmes (MQSUN). 2014 April. Pakistan Food Fortification Scoping Study. |
| Niedermaier 2021 | Niedermaier T, Gredner T, Kuznia S, Schottker B, Mons U, Brenner H. Potential of Vitamin D Food Fortification in Prevention of Cancer Deaths-A Modeling Study. Nutrients 2021; 13(11). |
| Niemesh 2015 | Niemesh GT. Ironing Out Deficiencies: Evidence from the United States on the Economic Effects of Iron Deficiency. Journal of Human Resources 2015; 50(4): 910-58. |
| Noshirvan 2021 | Noshirvan A, Wu B, Luo H, et al. Predicted Effects and Cost-Effectiveness of Wheat Flour Fortification for Reducing Micronutrient Deficiencies, Maternal Anemia, and Neural Tube Defects in Yaounde and Douala, Cameroon. Food and nutrition bulletin 2021; 42(4): 551-66. |
| Palacios 2022 | Palacios A, Rojas-Roque C, Balan D, et al. Fortification of staple foods with calcium: a novel costing tool to inform decision making. Annals of the New York Academy of Sciences 2022; 1513(1): 79-88. |
| Pandav 2012 | Pandav CS. Economic evaluation of iodine deficiency disorder control program in Sikkim: a cost-benefit analysis. Indian journal of public health 2012; 56(3): 214-22. |
| Patron 2016 | Patron AP, Detzel P, Hutton Z. Estimating the contribution on the reduction of the burden of iron deficiency anemia from the current fortification strategy for commercial foods targeted to infants and toddlers in India. Journal of Pediatric Gastroenterology and Nutrition 2016; 62(SUPPL. 1): 830. |
| Phillips 1996 | Phillips M, Sanghvi T, Suarez R, McKigney J, Fiedler J. The costs and effectiveness of three vitamin A interventions in Guatemala. Social science & medicine (1982) 1996; 42(12): 1661-8. |
| Popkin 1980 | Popkin BM, Solon FS, Fernandez T, Latham MC. Benefit-cost analysis in the nutrition area: A spoject in The Philippines. Social Science and Medicine 1980; 14 C(3): 207-16. |
| Prieto-Patron 2022 | Prieto-Patron A, Detzel P, Ramayulis R, Sudikno, Irene, Wibowo Y. Impact of Fortified Infant Cereals on the Burden of Iron Deficiency Anemia in 6- to 23-Month-Old Indonesian Infants and Young Children: A Health Economic Simulation Model. International journal of environmental research and public health 2022; 19(9). |
| Qureshy 2023 | Qureshy LF, Alderman H, Manchanda N. Benefit-Cost Analysis of Iron Fortification of Rice in India: Modelling Potential Economic Gains from Improving Haemoglobin and Averting Anaemia. Journal of Development Effectiveness 2023; 15(1): 91-110. |
| Rabovskaja 2013 | Rabovskaja V, Parkinson B, Goodall S. The cost-effectiveness of mandatory folic acid fortification in Australia. The Journal of nutrition 2013; 143(1): 59-66. |
| Rajkumar 2012 | Rajkumar AS, Gaukler C, Tilahun J. Combating malnutrition in Ethiopia: an evidence-based approach for sustained results. Washington DC: The World Bank; 2012. |
| Rochau 2019 | Rochau U, Schaffner M, Muhlberger N, et al. PDB51 Prevention as a public health measure: long-term effectiveness and cost effectiveness of a poulation-based iodine deficiency diseases prevention program. Value in Health 2019; 22(Supplement 3): S581-S2. |
| Rodrigues 2023 | Rodrigues VB, Silva ENd, Dos Santos AM, Santos LMP. Prevented cases of neural tube defects and cost savings after folic acid fortification of flour in Brazil. PloS one 2023; 18(2): e0281077. |

| Study ID (Author Year) | Bibliography |
|---|---|
| Romano 1995 | Romano PS, Waitzman NJ, Scheffler RM, Pi RD. Folic acid fortification of grain: an economic analysis. American journal of public health 1995; 85(5): 667-76. |
| Sablah 2012 | Sablah M, Klopp J, Steinberg D, Touaoro Z, Laillou A, Baker S. Thriving public-private partnership to fortify cooking oil in the West African Economic and Monetary Union (UEMOA) to control vitamin A deficiency: Faire Tache d'Huile en Afrique de l'Ouest. Food and nutrition bulletin 2012; 33(4 Suppl): S310-20. |
| Saing 2019 | Saing S, Haywood P, van der Linden N, Manipis K, Meshcheriakova E, Goodall S. Real-World Cost Effectiveness of Mandatory Folic Acid Fortification of Bread-Making Flour in Australia. Applied health economics and health policy 2019; 17(2): 243-54. |
| Sandmann 2017 | Sandmann A, Amling M, Barvencik F, Konig H-H, Bleibler F. Economic evaluation of vitamin D and calcium food fortification for fracture prevention in Germany. Public health nutrition 2017; 20(10): 1874-83. |
| Sayed 2008 | Sayed A-R, Bourne D, Pattinson R, Nixon J, Henderson B. Decline in the prevalence of neural tube defects following folic acid fortification and its cost-benefit in South Africa. Birth defects research Part A, Clinical and molecular teratology 2008; 82(4): 211-6. |
| Segal 2007 | Segal L, Dalziel K, Katz R. A report to FSANZ. Informing a strategy to increase folate levels to prevent neural tube defects: a cost-effectiveness analysis of options. Adelaide, Australia: Centre for Health Economics, Monash University and Division of Health Sciences, University of South Australia; 2007. |
| van Stuijvenberg 2001 | van Stuijvenberg ME, Dhansay MA, Lombard CJ, Faber M, Benade AJ. The effect of a biscuit with red palm oil as a source of beta-carotene on the vitamin A status of primary school children: a comparison with beta-carotene from a synthetic source in a randomised controlled trial. European journal of clinical nutrition 2001; 55(8): 657-62. |
| Vosti 2020a | Vosti SA, Kagin J, Engle-Stone R, et al. Strategies to achieve adequate vitamin A intake for young children: options for Cameroon. Annals of the New York Academy of Sciences 2020a; 1465(1): 161-80. |
| Vosti 2020b | Vosti S; Engle-Stone R; Woldegebreal, D; Luo, H; Asfaw, E; Kagin, J; Moges, T; Tesfaye, B. Estimated Effectiveness and Cost-effectiveness of a Fortified Edible Oils Program in Ethiopia. Curr Dev Nutr 2020b; 4: 1735. |
| Vosti 2023 | Vosti SA, Adams KP, Michuda A, et al. Impacts of micronutrient intervention programs on effective coverage and lives saved: Modeled evidence from Cameroon. Annals of the New York Academy of Sciences 2023; 1519(1): 199-210. |
| Walters 2019 | Walters D, Ndau E, Saleh N, Mosha T, Horton S. Cost-effectiveness of sunflower oil fortification with vitamin A in Tanzania by scale. Maternal & child nutrition 2019; 15 Suppl 3: e12720. |
| Wei 2023 | Wei Y, Ding G, Huo J, et al. [Cost-benefit analysis on the intervention of application iron-fortified soy sauce for anemia in Deqing 15-54 years women, Zhejiang]. Wei sheng yan jiu = Journal of hygiene research 2023; 52(3): 429-33. |

*Table S1*: **Characteristics of each individual study**

| Study ID* (Author Year) | Country | Setting | Population target | Source of funding | Conflicts of interest | Coverage | Regulatory context | Delivery platform | Type of economic evaluation | Model type or study design | Size of population | Perspective† | Types of costs captured | Time horizon |
|---|---|---|---|---|---|---|---|---|---|---|---|---|---|---|
| Access Economics 2006 | Australia, New Zealand | National | General population | Food Standards Australia New Zealand (FSANZ) | NR | NR | Mandatory fortification | open market | CBA | NR [presumably decision tree] | NR | NR [presumably Societal] | Disability and premature mortality, total outlays on health care and personal care, production losses, and efficiency losses that arise from lower taxation revenues and higher welfare payments; costs to government of administering and enforcing mandatory fortification and the costs to industry of fortifying their product. | Lifetime |
| Asian Develop. Bank 2004 | Indonesia, Pakistan, China, Thailand, Viet Nam | National | General population, pregnant women, WRA, men, children | Asian Development Bank and Keystone Center [comment: Unclear] | NR | NR | Mandatory fortification | Public/government and private/industry sectors | CBA | PROFILES model | NR | Societal | Equipment, Administration and Overhead (calculated as 3% of the premix cost), Labor, Quality Assurance, Recurring Production Costs (including fortificant premix and Food Control and Biological Monitoring Costs) | 10 years for benefit and costs |
| Australian HMAC 2017 | Australia, New Zealand | National | General population | Australian Health Ministers' Advisory Council (AHMAC) | NR | NR | Mandatory fortification | open market | CUA | Decision analytic framework; Markov model | 2 populations: 23,524,055; 4,509,680 | Societal | Costs for the health system, the population, government and manufacturers (salt manufacturers, bakers), including sunk costs | 70 years (Australia); 10 years (NZ) |
| Baltussen 2004 | 4 subregions: African, South American, European, Southeast Asian | 4 multinational WHO regions: AfrD, AmrB, EurA, SearD | Pregnant women | NR | NR | geographical coverage 50%, 80%, 95%; access 60-95% (based on region) | NR | NR | CEA | Population model; econometric model | 4 populations: 334,580,563; 411,889,100; 442,130,339; 1,334,810,348 | Societal | Industry production costs, including for scales, dosifiers and fortificant. Program costs at national, provincial and districts levels include management, legislation, health nutrition education and supervision. | NR [presumably lifelong; costs estimated for 10 years] |

| Study ID* (Author Year) | Country | Setting | Population target | Source of funding | Conflicts of interest | Coverage | Regulatory context | Delivery platform | Type of economic evaluation | Model type or study design | Size of population | Perspective† | Types of costs captured | Time horizon |
|---|---|---|---|---|---|---|---|---|---|---|---|---|---|---|
| Bentley 2009 | USA | National | Adults | Dana-Farber/Harvard Cancer Center Program in Cancer Outcomes Research Training, National Cancer Institute, and CISNET. | There are no conflicts of interest. | NR | Mandatory fortification | NR | CUA | Markov model | NR | NR [presumably Healthcare sector] | Disease-related costs (for NTDs, MIs, colorectal cancers, B12 deficiency maskings), including short-term care, outpatient care, medications, diagnostics; and fortification costs | Lifetime |
| Chow 2010 | India | Subset of India's states comprising a set of rural and urban areas | Preschool children; Pregnant women | International Center for Tropical Agriculture (CIAT), Colombia. | The authors have declared that no competing interests exist. | NR | NR | Industrial | CEA & CBA | NR [presumably decision tree] | NR | NR [presumably Industry/Food fortification provider] | Production costs (Cost to fortify 1 kg of oil), Implementation and monitoring and surveillance costs (Administrative and regulatory costs, training, promotional and educational materials, and program monitoring and evaluation -per kg of oil), Product costs (Bottling costs - per l of oil) | 20 years |
| Connelly 1996 | Australia | National | High risk of developing Wernicke-Korsakoff syndrome | NR | NR | NR | Hyphothetical | NR | CEA | NR [presumably decision tree] | NR | NR [presumably Public health sector] | Fortification costs, including costs of equipment and its installation, costs of fortificant, monitoring/assay costs & other recurrent costs | 40 years |
| Dainelli 2017 | China | National | 50–79 year olds | Nestlé Research Center of Beijing (China) and the Nestlé Research Center of Lausanne (Switzerland). | Hai Fang and Yangfeng Wu were paid as consultants by Nestlé. | Currently 8.67% of 50–79 year olds who regularly consume milk (modelled reach) | NR | NR | CUA | Markov model | 362.74 million | Societal | Medical costs (OP drug costs), Outpatient visits, Hypertension screening costs, Inpatient costs and days, Indirect costs (lost work days, salary) | 30 years |
| Dalziel 2010 | Australia, New Zealand | National level - government | Women of childbearing age (19-44 years old) | Food Standards Australia and New Zealand (FSANZ) | The authors declare that they have no conflicts of interest | NR | Mandatory fortification | NR | CEA | NR | 2 populations: 19,832,791; 3,244,250 | Societal | Low cost scenario (FSANZ-commissioned estimates); or High cost scenario (industry-commissioned estimates). 1) | 80 years |

| Study ID* (Author Year) | Country | Setting | Population target | Source of funding | Conflicts of interest | Coverage | Regulatory context | Delivery platform | Type of economic evaluation | Model type or study design | Size of population | Perspective† | Types of costs captured | Time horizon |
|---|---|---|---|---|---|---|---|---|---|---|---|---|---|---|
| | | | | | | | | | | | | | Downstream treatment costs incurred by the health sector; 2) Industry fortification upfront costs (including labelling, packaging write off, and equipment); 3) Industry fortification ongoing costs (including folic acid, premix, analytical testing, administration, mill using); 4) Government enforcement costs (including training and awareness, auditing, administration, complaints, and enforcement); 5) Monitoring/evaluation and Public health campaign. | |
| Detzel 2016 | India | National | Children 6-23 months old | NR | NR [author affiliated with Nestle] | NR | Unclear | open market | CEA | NR | NR | Social | intangible costs, production losses | NR |
| Edejer 2005 | Sub-Saharan Africa, South East Asia | 2 multinational WHO regions: Afr-E, Sear-D (both consisting of countries with high rates of child mortality) | Children <5 years old likely to be affected by pneumonia, diarrhea, and measles | None | None declared | 3 levels: 50%, 80%, and 95% | Mandatory fortification | Either locally produced or imported, or whether for industrial or domestic use | CEA | PopMod population model | NR | Societal | Production costs, those incurred during the fortification process | 10 years for costs; related benefits anytime |
| Fiedler 2000 | Philippines | National | Children 12-59 months old | US Agency for International Development under the auspices of the Opportunities for Micronutrient Interventions (OMNI) Project. | NR | Unclear | Government policy | Open market | CEA | NR | NR | NR [presumably Industry/Food fortification provider] | production costs, promotion of the program, internal and external monitoring costs | 1 year |

| Study ID* (Author Year) | Country | Setting | Population target | Source of funding | Conflicts of interest | Coverage | Regulatory context | Delivery platform | Type of economic evaluation | Model type or study design | Size of population | Perspective† | Types of costs captured | Time horizon |
|---|---|---|---|---|---|---|---|---|---|---|---|---|---|---|
| Fiedler 2009 | Afghanistan, Angola, Bangladesh, Bolivia, Brazil, Burkina Faso, Burundi, Cambodia, Cameroon, Chad, China, Dem. Rep. Congo, Cote d'Ivoire, Egypt, Ethiopia, Ghana, Guatemala, Guinea, India, Indonesia, Kenya, Madagascar, Malawi, Mali, Mexico, Morocco, Mozambique, Myanmar, Nepal, Niger, Nigeria, Pakistan, Peru, Philippines, Rwanda, Sierra Leone, South Africa, Sudan, Tanzania, Turkey, Uganda, Uzbekistan, Vietnam, Yemen, Zambia, Zimbabwe | National | general population (for iron); children (for zinc); children and pregnant/lactating women (for vitamin A) | Global Alliance for Improved Nutrition (GAIN), A2Z: USAID Micronutrient Project, the World Bank, and the Micronutrient Initiative. | NR | 25%-100% current consumptions (depending on intervention) | NR | open market | CEA | Model type NR; used algorithm-based spreadsheet tools (production functions) and HarvestPlus DALYs methodology. | NR | Social | Private and public sector costs: production, quality control/regulatory activities, marketing, public health impact evaluation | 10 years for costs & benefits |
| Fiedler 2010 | Uganda | National | Children | US Agency for International Development; | NR | 71% | Mandatory fortification | open market | CEA | NR | NR | Public health sector | private sector: premix, testing, freight, import fees; and/or additional | NR |

| Study ID* (Author Year) | Country | Setting | Population target | Source of funding | Conflicts of interest | Coverage | Regulatory context | Delivery platform | Type of economic evaluation | Model type or study design | Size of population | Perspective† | Types of costs captured | Time horizon |
|---|---|---|---|---|---|---|---|---|---|---|---|---|---|---|
| | | | | A2Z: The Micronutrient and Child Blindness Project | | | | | | | | | production costs, staff, tank, labels, etc. | |
| Fiedler 2012 | India | Gujarat state | Preschool children, pregnant and lactating women; people below the poverty line or with low incomes; school students | NR | NR | Beneficiaries of the SSNP programmes constituted 61.4% of Gujarat population | Collaboration between state, non-government and industry sectors | 3 major social safety net programs | CEA | NR | 3 populations: 1,741,045; 3,935,214; 17,513,420 | NR [presumably Industry/Food fortification provider and Public sectors] | Private sector flour production costs and public sector costs (monitoring and regulating the production) | Unclear |
| Fiedler 2013 | Zambia | National | General population | NR | NR | Coverages: 23% (Maize), 45% (Wheat flour), 59% (Oil), 60% (Sugar); 66% (sugar + oil), 82% (Maize + Wheat flour + Oil + Sugar), 70% (Wheat flour + Sugar), 73% (Oil + Sugar & Wheat flour + Oil + Sugar), 77% (Maize + Oil + Sugar), 80% (Wheat flour + Oil + Sugar) | Mandatory fortification for sugar, voluntary fortification for maize meal and wheat flour, hyphotethical fortification for vegetable oil | Industrial delivery system | CEA | NR | 11,436,380 | NR [presumably Consumer] | Equipment for production of fortified product (including cost of micro feeder/dosifier/mixing tank/blender; cost of laboratory equipment and glassware); cost of premix utilized annually; cost of fortificant utilized annually (including transport); annual cost of operating micro feeder; equipment operating and maintenance, quality assurance/quality control; monitoring and public sector costs | Unclear |
| Fiedler 2014 | Zambia | National | General population | Bill & Melinda Gates Foundation's Nutrition and Economic Research Support to HarvestPlus | NR | Modeled: Maize meal 29%; Wheat flour 57%; Vegetable oil 67%; Sugar 69% | Unclear | Public-private sector | CEA | NR | NR | Societal | NR (total costs) | 30 years for both costs and DALYs |
| Fiedler 2015 | Bangladesh | National | General population | Bill & Melinda Gates Foundation's Nutrition and | NR | National current consumption rates: | Voluntary fortification | NR | CEA | NR | 138,817,749 | NR [presumably Industry/Food fortification provider and Public sectors] | Equipment for production of fortified product; Cost of mixing tank; | NR [possibly 15 years for costs; |

| Study ID* (Author Year) | Country | Setting | Population target | Source of funding | Conflicts of interest | Coverage | Regulatory context | Delivery platform | Type of economic evaluation | Model type or study design | Size of population | Perspective† | Types of costs captured | Time horizon |
|---|---|---|---|---|---|---|---|---|---|---|---|---|---|---|
| | | | | Economic Research Support to HarvestPlus | | Vegetable oil 76%; Wheat flour–containing foods 65%; Either vegetable oil or wheat flour foods 91%; Both vegetable oil and wheat flour foods 50%. | | | | | | | Laboratory equipment and glassware ; Cost of fortificant ; Annual cost fortificant utilized (including transport); Annual cost of operating micro feeder; Equipment operating and maintenance, quality assurance/quality control; monitoring and public sector costs | endline NR for health] |
| GAIN 2017 | Afghanistan | National | General population | USAID | NR | >25-30% of households buy wheat flour nationally | Mandatory fortification | open market | CBA | Spreadsheet based model | 32,564,342 | NR [presumably Societal] | Flour production and fortification costs (initial and ongoing); government costs (regulation, inspection); future earnings losses in children with deficiencies, earnings losses in adults, neonatal and maternal mortality, neural tube defects (mortality, lost productivity, acute and ongoing health care). | 10 years |
| Ghauri 2015 | Pakistan | National | General population | GAIN | NR | Coverage used in model: 45% in 2015 to 56% in 2024 | Mandatory fortification | open market | CBA | NR | 182,520,000 (2015) - 227,942,469 (2024) | Societal | Economic losses due to deficiency/malnutrition relating to neural tube defects, neonatal & maternal mortality, adult and projected children's productivity losses; wheat flour production and fortification costs, government program management costs. | 10 years |
| Ghauri 2016 | Tajikistan | National | General population | USAID | NR | 93% target | Mandatory fortification | open market | CBA | Spreadsheet based model | 5,744,158 (2017) - 6,804,466 (2026) | NR [presumably Societal] | Economic losses due to deficiency/malnutrition relating to neural tube defects, neonatal & maternal mortality, adult and projected | 10 years |

| Study ID* (Author Year) | Country | Setting | Population target | Source of funding | Conflicts of interest | Coverage | Regulatory context | Delivery platform | Type of economic evaluation | Model type or study design | Size of population | Perspective† | Types of costs captured | Time horizon |
|---|---|---|---|---|---|---|---|---|---|---|---|---|---|---|
| | | | | | | | | | | | | | children's productivity losses; wheat flour production and fortification costs, government program management costs. | |
| Gorstein 2020 | Worldwide (>100 countries) | International | Populations of 139 countries | No funding was received. | No competing financial interests exist | 88% in 2019 (low-income countries) | Universal salt iodization | Open market | CEA | Consequence modeling | >7.1 billion | NR [presumably Societal] | Iodine deficiency disorder- and reduced IQ-related productivity deficit (future earnings); regional demographic and labor data | 50 years |
| Grosse 2016 | USA | National | Women of childbearing age | Publication of this article has been sponsored by the Centers for Disease Control and Prevention (CDC), Office of the Associate Director for Policy | No financial disclosures were reported by the authors of this paper. | NR | Mandatory fortification | FDA program | CEA | Unclear [presumably decision tree] | NR | Societal | Production Costs (those costs incurred during the fortification process), unpaid caregiver time costs, lifetime direct cost of a live-born spina bifida case (including medical and special education and developmental services) | Lifetime |
| Hoddinott 2018 | Zambia | National | WRA, pregnant women | Through Nutrition International by a grant provided by the Bill & Melinda Gates Foundation | The author declares no competing interests. | NR | Mandatory fortification | Open market | CEA | NR | 670,000 live births per year | NR [presumably societal] | Production equipment; social marketing and communications; for monitoring and evaluation; program management and implementation; folic acid in premix | 10 years |
| Horton 2003 | Bangladesh, India, Mali, Tanzania, Egypt, Oman, Bolivia, Honduras, Nicaragua | National | General population | Canadian International Development Agency / Micronutrient Initiative | NR | 100% | Mandatory fortification | national iron fortification program | CBA | NR | NR | NR [presumably societal] | Production costs; Productivity losses averted resulting from degraded physical and cognitive performance | Lifetime |
| Horton 2008 | South Asia, Sub-Saharan Africa, CEE/CIS | National | General population | NR | NR | Scale up to: 80% of households | NR | open market | CBA | NR | 380,000,000 (numbers affected) | Societal | Cost of iodisation of costs associated with losses of life, earnings and disability | Lifetime |
| Huang 2020 | China | National | Children 8-10 years old | NR | NR | 100% | Mandatory fortification | State-controlled market | CBA | NR | 19,520,000 | NR | aggregate cost of USI (iodization/production, distribution) | NR |

| Study ID* (Author Year) | Country | Setting | Population target | Source of funding | Conflicts of interest | Coverage | Regulatory context | Delivery platform | Type of economic evaluation | Model type or study design | Size of population | Perspective† | Types of costs captured | Time horizon |
|---|---|---|---|---|---|---|---|---|---|---|---|---|---|---|
| Jentink 2008 | Netherlands | National | Women of childbearing age | NR | Conflicts of interest: None declared. | 11% of unprotected women (i.e. not taking folate supplements) covered by fortification | Theoretical mandatory fortification | open market | CUA | Decision/disease progression tree simulation model | 187,910 newborns | Societal | Total hospital costs; travel costs for parental hospital visits; physiotherapy costs; assistive technology/house adaptations; special education; productivity losses; fortification costs | Lifetime |
| Johnson 2021 | Ethiopia | National | General population | GAIN | NR | NR | Mandatory fortification | open market | CBA | Cost benefit analysis framework & cost modelling tools | 112,859,630 (2019) to 140,822,503 (2028) | Societal | Lost productivity, health care costs; start-up equipment (& training, testing); vitamin A premix paid by the refineries; Government costs (staff training, public education and social marketing, capital improvement for testing, monitoring and enforce quality, baseline survey). | 10 years |
| Kakietek 2018 | Bangladesh | National | General population | UK Aid from the UK government, European Commission (EC) through the South Asia Food and Nutrition Security Initiative (SAFANSI), which is administered by the World Bank. | NR | Coverage to reach 90% over 10 years | Mandatory fortification of oil; Rice NR [hypothetical] | Social safety net program | CEA | NR | NR | Public health sector | Only public sector costs are included here (with majority of costs to be borne by households): waiving duties and tax on fortificant, and quality control | 10 years |
| Kancherla 2021 | Angola, Bangladesh, Benin, China, Côte d'Ivoire, Egypt, Ethiopia, Ghana, India, Liberia, | National | Children and pregnant/WRA | This research received no external funding | The authors declare no conflict of interest | Reach modelled: Angola 9 Million, Bangladesh 144.6 M, Benin 10.3 M, China 1284.9 M, | Modeled mandatory | open market | CBA | NR | 25% of national population | NR [presumably Industry/Food fortification provider and Public sectors] | Production costs; future health and economic impacts (global fortification at the aggregate level was estimated using a global unit cost) | Expected lifespan of a child in each country |

| Study ID* (Author Year) | Country | Setting | Population target | Source of funding | Conflicts of interest | Coverage | Regulatory context | Delivery platform | Type of economic evaluation | Model type or study design | Size of population | Perspective[†] | Types of costs captured | Time horizon |
|---|---|---|---|---|---|---|---|---|---|---|---|---|---|---|
| | Nigeria, Senegal, Tajikistan | | | | | Côte d'Ivoire 12.3 M, Egypt 88.6 M, Ethiopia 84.5 M, Ghana 20 M, India 553.1 M, Liberia 2.8 M, Nigeria 109.4, Senegal 14.2 M, Tajikistan 7.8 M | | | | | | | | |
| Llanos 2007 | Chile | National | Pregnant/WRA | NR | NR | NR | Mandatory wheat flour fortification | NR | CEA & CBA | NR [presumably decision tree] | 117,101 | NR [presumably Healthcare sector] | Fortification and testing costs; and long-term medical,developmental services, and rehabilitation care costs | 22 years for benefits; 1 year for costs |
| Ma 2008 | China | National | Children, adults, pregnant and lactating women, WRA | Ministry of Health and the Ministry of Science and Technology, China | NR | NR [universal] | NR | NR | CEA | Standard World Health Organization ingredients approach | NR | NR | NR [using standard WHO ingredients approach] | NR |
| Mardones-Santander 1991 | Chile | National | Low weight/height pregnant women | FONDECYT-CHILE and Melkunie Holland | NR [1 author affiliated with Melkunie Holland] | 83.5% | Government provision | social safety net program | CEA | NR | 65,123 | NR [presumably Healthcare sector] | Costs of the food product and for healthcare of surviving infants, including: hospital care, nutritional rehabilitation, and ambulatory treatment | 1 year |
| Moges 2020 | Ethiopia | National | Children 6-35 months old; and WRA | Bill & Melinda Gates Foundation | NR | Program reach: oil 59%; flour 22% for WRA, 28% for children | Modeled mandatory fortification | National oil and wheat flour fortification programs | CEA | Lives Saved Tool (LiST) model; cost modeling | Flour: 283,416,764 person-years (from 2012-2020) | NR [presumably Societal] | start-up investments, premix costs, and recurring private and public sector M&E and inspection costs | 10 years |
| MQSUN 2014 | Pakistan | National | WRA (15-49 years); and Children (<5 years) | UKaid / UK Government's Department for International Development | NR | target coverage: 65% in rural areas and 85% in urban areas; current coverage: 0 | Mandatory fortification | open market/private industry | CEA & CBA | Unclear (counterfactual-based analysis) | NR | NR [presumably Industry/Food fortification provider and Public sectors] | 1) fortification costs, 2) production capital costs, 3) in-mill quality control (QC) and other production recurrent costs, 4) public quality assurance, 5) future economic | Unclear for benefits, 5 and 10 years for costs |

| Study ID* (Author Year) | Country | Setting | Population target | Source of funding | Conflicts of interest | Coverage | Regulatory context | Delivery platform | Type of economic evaluation | Model type or study design | Size of population | Perspective[†] | Types of costs captured | Time horizon |
|---|---|---|---|---|---|---|---|---|---|---|---|---|---|---|
| | | | | | | | | | | | | | consequences from morbidity and/or mortality averted. | |
| Niedermaier 2021 | Germany | National | General population | No external funding | No conflict of interest | NR | NR | open market | CEA | NR [conducted literature review] | 83,100,000 | NR [presumably Societal] | Production costs, marketing, education costs, food control, monitoring costs, other program-specific recurrent production costs | NR |
| Niemesh 2015 | USA | National | General population | John E. Rovensky Fellowship, the Vanderbilt University Summer Research Awards Program in Arts and Science, Graduate School Dissertation Enhancement Program, Noel Dissertation Fellowship, and Kirk Dornbush Summer Research Grant | NR | Mandatory fortification (bread). 70-80% (flour, voluntary) | Mandatory fortification | Open market | CBA | NR | NR | NR [presumably Societal] | Costs of production | 10 years |
| Noshirvan 2021 | Cameroon | 2 urban areas: Yaoundé and Douala | Women of reproductive age and children 6-59 months | Bill & Melinda Gates Foundation; UC Davis Medical Student Research Fellowship (AN and BW). | The author(s) declared no potential conflicts of interest with respect to the research, authorship, and/or publication of this article. | 98% | Mandatory wheat flour fortification | NR | CEA | NR [presumably decision tree based on epidemiological population model] | 4,909,950 | NR [presumably Industry/Food fortification provider and Public sectors] | Baseline survey, industrial assessment, revision of standards, equipment for industry, micronutrient premix, equipment for the national lab, training of partners and stakeholders, launch activities, supervision costs, other indirect costs, monitoring and evaluation (incl. start-up monitoring/evaluation, and ongoing monitoring/evaluation) | 13 years |

| Study ID* (Author Year) | Country | Setting | Population target | Source of funding | Conflicts of interest | Coverage | Regulatory context | Delivery platform | Type of economic evaluation | Model type or study design | Size of population | Perspective† | Types of costs captured | Time horizon |
|---|---|---|---|---|---|---|---|---|---|---|---|---|---|---|
| Palacios 2022 | Costa Rica | National | General population | UNDP–UNFPA–UNICEF–WHO–Work Bank Special Programme of Research Development and Research Training in Human Reproduction (HRP) at WHO | The authors declare no competing interests. | mandatory and full-coverage calcium intervention program (100%) | Mandatory fortification | open market | CEA | Novel costing tool | 4,999,443 | Industry/Food fortification provider and Public sectors | Costs of the premix fortifier (that includes the micronutrients needed in the fortification process), the costs of the fortifier salt, the costs of the internal and external quality control processes, wages, marketing and labeling costs, the costs of the logistic supplies, the preventive costs for the installed physical equipment, costs regarding to the calibration of the equipment, and the costs of the final product logistics. | 1 year |
| Pandav 2012 | India | Sikkim state | General population | International Council for Control of Iodine Deficiency Disorders (ICCIDD), the All India Institute of Medical Sciences (AIIMS), New Delhi and United States Agency for Aid in International Development (USAID) through INCLEN Fellowship program | None declared. | 100% | Mandatory (Universal) iodized salt program | Governmental program | CBA | NR [presumably decision tree] | 403,612 | Societal | Salt iodization, monitoring, Communication Campaign | lifetime (60 years) |
| Phillips 1996 | Guatemala | National | Women of child-bearing age and children <6 years old | NR | NR | 90% | Three different doses evaluated: level mandated by | open market | CEA | NR | 9.2 million | NR [presumably Industry/Food fortification provider] | Recurrent and capital costs of operating the program | 1 year |

| Study ID* (Author Year) | Country | Setting | Population target | Source of funding | Conflicts of interest | Coverage | Regulatory context | Delivery platform | Type of economic evaluation | Model type or study design | Size of population | Perspective† | Types of costs captured | Time horizon |
|---|---|---|---|---|---|---|---|---|---|---|---|---|---|---|
| | | | | | | | law; levels from a 1989 study; levels in a 1992/93 INCAP study | | | | | | | |
| Popkin 1980 | Philippines | 4 coastal and squatter areas | Children aged 1-16 years at risk of eye disease/blindness due to Vitamin A deficiency | Office of Nutrition (Philippines); U.S. Agency for International Development; the Philippine National Science Development board; World Health Organization | NR | 100% modelled | Mandatory fortification | Unclear | CBA | NR | 900 | NR [presumably Societal] | Costs of production, costs of health care, costs of lost/gained earnings. | Lifetime |
| Prieto-Patron 2022 | Indonesia | National | Children 6-23 months old | Nestle Infant Nutrition Indonesia and Nestle Research | Prieto Patron and Yulianti Wibowo are employees of Nestlé. Patrick Detzel and Irene were previous employee of Nestlé when the manuscript was drafted. Nestlé is a company selling Fortified Infant Cereals. | 74% | Current voluntary fortification level | open market | CEA | Simulation model | 3,923,000 | Societal | Lowered future income from impaired cognitive or physical development | 71 years |
| Qureshy 2023 | India | National | General population with mild/moderate anemia (excluding young children and pregnant/lactating women) | Government of Japan through the Japan Trust Fund for Scaling Up Nutrition - Bill & Melinda Gates Foundation through | NR | NR | Voluntary fortification | social safety net program | CBA | Benefit cost model | 548,859,082 (Midday Meal scheme + Public distribution system group) | NR [presumably Societal] | Fortification cost, blending, and depreciation, opportunity cost of raising revenue for financing the fortification (deadweight loss) | 40 years |

| Study ID* (Author Year) | Country | Setting | Population target | Source of funding | Conflicts of interest | Coverage | Regulatory context | Delivery platform | Type of economic evaluation | Model type or study design | Size of population | Perspective† | Types of costs captured | Time horizon |
|---|---|---|---|---|---|---|---|---|---|---|---|---|---|---|
| | | | | Partnerships and Opportunities to Strengthen and Harmonize Actions Against Malnutrition in India (POSHAN), led by the International Food Policy Research Institute (IFPRI) | | | | | | | | | | |
| Rabovskaja 2013 | Australia | National | General population | Supported by the Faculty of Business, University of Technology Sydney | Author disclosures: V. Rabovskaja, B. Parkinson, and S. Goodall, no conflicts of interest | NR | Mandatory fortification | Open market | CUA | Decision tree model | NR | Societal | Industry and regulatory costs to the government; health system costs of neonatal death, NTDs, spina bifida, anencephaly, and vitamin B-12 deficiency and its neurologic consequences | Lifetime |
| Rajkumar 2012 | Ethiopia | National | General population | World Bank (preparation & publication) | NR | Modeled: sugar 41%; salt 99.9% | Unclear | Open market | CEA & CBA | NR | 78,000,000 | NR [presumably Societal] | Production, social costs/gains | Unclear |
| Rochau 2019 | Germany | National | Hypothetical population with moderate iodine deficiency | NR | NR | NR | NR | NR | CUA | Markov model: state-transition open cohort model | 82,792,351 | NR [presumably Societal] | Direct medical costs, direct non-medical costs. [comment: Indirect costs were considered in an additional analysis] | Lifetime |
| Rodrigues 2023 | Brazil | Ministry of Health's outpatient and hospital information system databases | Women of reproductive age | No funding received | The authors have declared that no competing interests exist. | NR | Mandatory folic acid fortification | NR | CEA | primary study (observational study) | NR | Healthcare sector | outpatient and hospital care | 10 years |
| Sablah 2012 | West African Economic and Monetary Union (UEMOA): Benin, | Multinational initiative: "Faire Tache d'Huile en Afrique de l'Ouest" | Infants and children <5 years of age not already being protected by vitamin A supplements | The major funding agencies for the Tache d'Huile initiative are the Global Alliance for Improved | The authors confirm that they do not have any potential financial or any other | 70% | Mandatory fortification | Open market | CEA | NR | 16,600,000 | NR [presumably Industry/Food fortification provider and Public sectors] | private sector, public sector and other stakeholders' costs | Unclear [life expectancy for benefits] |

| Study ID* (Author Year) | Country | Setting | Population target | Source of funding | Conflicts of interest | Coverage | Regulatory context | Delivery platform | Type of economic evaluation | Model type or study design | Size of population | Perspective† | Types of costs captured | Time horizon |
|---|---|---|---|---|---|---|---|---|---|---|---|---|---|---|
| | Burkina Faso, Côte d'Ivoire, Guinea-Bissau, Mali, Niger, Senegal, Togo | | | Nutrition (GAIN), the Michael & Susan Dell Foundation, the Micronutrient Initiative, the Government of Taiwan, UNICEF, and the US Agency for International Development. implementation agencies include Helen Keller International, UEMOA, the West Africa Health Organization, UNICEF, the Professional Association of Cooking Oil Industries of the West African Economic and Monetary Union (AIFO-UEMOA), and the technical ministries of the governments of the member countries of UEMOA. | conflicts of interest in publishing this manuscript. | | | | | | | | | |
| Sandmann 2017 | Germany | National | Elderly women (>65 years old) | This research received no specific grant from any funding agency in the public, commercial or not- for-profit sectors. | Conflict of interest: None. | 82% | NR | open market | CEA & CBA | Spreadsheet model [presumably decision tree] | NR | Societal | Inpatient costs, outpatient costs, home care costs, long-term care costs, fortification, marketing and education costs | 36 years |

| Study ID* (Author Year) | Country | Setting | Population target | Source of funding | Conflicts of interest | Coverage | Regulatory context | Delivery platform | Type of economic evaluation | Model type or study design | Size of population | Perspective† | Types of costs captured | Time horizon |
|---|---|---|---|---|---|---|---|---|---|---|---|---|---|---|
| Sayed 2008 | South Africa | 12 sentinel public hospitals in 4 provinces | Pregnant women and new births | University of Cape Town, South African Medical Research Council, South African Department of Health | Competing Interests: None. | ~90% | National Food Fortification Program | Open market | CBA | primary study (observational study) | 1.05 million births | NR [presumably Healthcare sector] | Direct cost of fortification; direct cost of minimal medical intervention in the short term (3 years) | 3 years |
| van Stuijvenberg 2001 | South Africa | Rural primary schools in KwaZulu-Natal | Children 5-11 years old | Palm Oil Research Institute of Malaysia (PORIM) | NR | 100% | NR | School distribution | CEA | primary study (RCT) | 267 | NR [presumably Consumer] | production costs for biscuits | 3 months |
| Vosti 2023 | Cameroon | National | Zinc: Children; Vitamin A: Children; Folic acid: Women of reproductive age | Bill and Melinda Gates Foundation; Mars Inc. gift to UC Davis for interdisciplinary research | The authors declare no competing interests. | Effective coverage: Zinc flour 15-27%; Zinc bouillon 9%; Vitamin A oil 15-18%; Vitamin A bouillon 19%; Folic acid flour 28-44%; Folic acid bouillon 60%. | Mixed (depending on intervention program) | NR | CEA | Micronutrient Intervention Modeling (MINIMOD) tool, economic optimization model (mixed integer linear programming) | Effective coverage: 4,567,575 (zinc); 14,233,468 (vitamin A); 17,096,476 (folic acid). | NR [presumably Industry/Food fortification provider and Public sectors] | costs of the specific fortificants or compounds included in the premix, plus the factory and public sector costs associated with quality assurance and regulation | 10 years |
| Walters 2019 | Tanzania | National | Children 6-59 months old and their lactating mothers | Global Affairs Canada; International Development Research Centre (IDRC) | The authors declare no conflicts of interest in relation to this study. | NR | Mandatory fortification | NR | CEA | NR [Differences-in-differences (DIDs) regression analysis conducted] | NR | Societal | Private sector start-up and recurrent costs, and public sector costs for regulation, social marketing, monitoring, and programme management. | 1 year |
| Wei 2023 | China | Deqing County, Zhejiang Province | Women 15-54 years old | NR | NR | Purchase rate amongst study participants: 76% | NR | Open market | CEA & CBA | NR | 585 | NR | Iron-fortified soy sauce purchase costs; start-up, publicity, coordination, lectures, training, supervision and other work, as well as the cost of investment by iron-fortified soy sauce production enterprises; annual income loss | 15 months |

*See the 'List of Included Studies' IDs' above for the full bibliography.

†The point of view of an economic study: a societal perspective considers costs and benefits for society; and a healthcare perspective includes costs and effects on the health sector.

Abbreviations: CBA=cost-benefit analysis; CE= cost-effectiveness; CEA= cost-effectiveness analysis; CUA= cost-utility analysis; DALY=disability-adjusted life year; ICER=incremental cost-effectiveness ratio; N/A=not applicable; NR=not reported; NTDs=neural tube defects; QALY=quality-adjusted life year; WRA=women of reproductive age.

*Table S2*: **Results of each individual study and evaluation, grouped by Micronutrient (then by Country, followed by Health outcome type)**

| Study ID* (Author Year) | INTERVENTION-Micronutrient(s) | INTERVENTION-Food Vehicle(s) | Dose | Other Specs | Compar-ator | Country | Curr-ency | Price Year | HEALTH Outcome Type | Health Result - Difference | Costs - Difference *as REPORTED* | Incremental CE Ratio *as REPORTED* | CONVERTED ICER (using 2022 US$)† | Cost-Benefit Analysis | Dis-count Rate | Uncertainty & Sensitivity Analyses |
|---|---|---|---|---|---|---|---|---|---|---|---|---|---|---|---|---|
| Fiedler 2009 | Vitamin A | Sugar | 7.5-20 mg retinol (RE)/kg | Retinyl palmitate; Industrial | Pre-fortification | Afghanistan | US$ | 2009 | DALY saved | NR | 38.27 million | 42 | 45 | NR | 3% for DALYs; 0% for costs | NR |
| Edejer 2005 | Vitamin A | Sugar | 80% coverage; dose NR | NR | No fortification | Afr-E (sub-Saharan Africa) region | international dollars ($Int) | 2000 | DALYs averted yearly (millions) | 0.38 | 13,000,000 yearly | ICER $ 34/DALY saved | 137 | NR | 3% for both costs and benefits | Removal of age weighting and discounting for DALYs makes the interventions more cost effective. Varying geographical coverage from 80% to: 50% or 95% gave ICERs of I$ 41 or 32 per DALY saved (resp'ly). |
| Fiedler 2009 | Vitamin A | Vegetable oil | 1.5-4.0 mg retinol (RE)/kg | Retinyl palmitate; Industrial | Pre-fortification | Angola | US$ | 2009 | DALY saved | NR | 39.32 million | 110 | 148 | NR | 3% for DALYs; 0% for costs | NR |
| Fiedler 2009 | Vitamin A | Sugar | 7.5-20 mg retinol (RE)/kg | Retinyl palmitate; Industrial | Pre-fortification | Bangladesh | US$ | 2009 | DALY saved | NR | 205.45 million | 419 | 565 | NR | 3% for DALYs; 0% for costs | NR |
| Fiedler 2015 | Vitamin A | Vegetable oil | 15 mg per kg | retinyl palmitate 1.7 mIU/g | No fortification | Bangladesh | US$ | 2012 (unclear) | DALYs saved | 406,877 | 1,270,000 annually | ICER $ 3.12/ DALY saved | 6 | NR | Unclear (costs & benefits were discounted) | NR |
| Fiedler 2009 | Vitamin A | Sugar | 7.5-20 mg retinol (RE)/kg | Retinyl palmitate; Industrial | Pre-fortification | Bolivia | US$ | 2009 | DALY saved | NR | 32.59 million | 658 | 1,057 | NR | 3% for DALYs; 0% for costs | NR |
| Fiedler 2009 | Vitamin A | Vegetable oil | 1.5-4.0 mg retinol (RE)/kg | Retinyl palmitate te; | Pre-fortification | Bolivia | US$ | 2009 | DALY saved | NR | 9.57 million | 196 | 315 | NR | 3% for DALYs; 0% | NR |

| Study ID* (Author Year) | INTERVENTION-Micronutrient(s) | INTERVENTION-Food Vehicle(s) | Dose | Other Specs | Compar-ator | Country | Curr-ency | Price Year | HEALTH Outcome Type | Health Result - Difference | Costs - Difference *as REPORTED* | Incremental CE Ratio *as REPORTED* | CONVERTED ICER (using 2022 US$)† | Cost-Benefit Analysis | Dis-count Rate | Uncertainty & Sensitivity Analyses |
|---|---|---|---|---|---|---|---|---|---|---|---|---|---|---|---|---|
| | | | | Industrial | | | | | | | | | | | for costs | |
| Fiedler 2009 | Vitamin A | Sugar | 7.5-20 mg retinol (RE)/kg | Retinyl palmitate; Industrial | Pre-fortification | Burkina Faso | US$ | 2009 | DALY saved | NR | 15.47 million | 81 | 81 | NR | 3% for DALYs; 0% for costs | NR |
| Fiedler 2009 | Vitamin A | Vegetable oil | 1.5-4.0 mg retinol (RE)/kg | Retinyl palmitate; Industrial | Pre-fortification | Burkina Faso | US$ | 2009 | DALY saved | NR | 34.66 million | 182 | 183 | NR | 3% for DALYs; 0% for costs | NR |
| Fiedler 2009 | Vitamin A | Vegetable oil | 1.5-4.0 mg retinol (RE)/kg | Retinyl palmitate; Industrial | Pre-fortification | Burundi | US$ | 2009 | DALY saved | NR | 5.50 million | 106 | 178 | NR | 3% for DALYs; 0% for costs | NR |
| Fiedler 2009 | Vitamin A | Sugar | 7.5-20 mg retinol (RE)/kg | Retinyl palmitate; Industrial | Pre-fortification | Cameroon | US$ | 2009 | DALY saved | NR | 43.10 million | 420 | 402 | NR | 3% for DALYs; 0% for costs | NR |
| Fiedler 2009 | Vitamin A | Vegetable oil | 1.5-4.0 mg retinol (RE)/kg | Retinyl palmitate; Industrial | Pre-fortification | Cameroon | US$ | 2009 | DALY saved | NR | 39.05 million | 233 | 223 | NR | 3% for DALYs; 0% for costs | NR |
| Vosti 2023 | Vitamin A | Bouillon cubes | 80 mg/kg | NR | No fortification | Cameroon | US$ | 2019 | Child lives saved | 11,070 | Total program costs US $ 9,802,000 | 885 | 966 | NR | NR | The program selection of "high" or "low" effectiveness results did not influence the key policy messages. |
| Vosti 2023 | Vitamin A | Edible oils | 9 mg/kg | NR | No fortification | Cameroon | US$ | 2019 | Child lives saved | 11,619 | Total program costs US $ 4,455,000 | 383 | 393 | NR | NR | The program selection of "high" or "low" effectiveness results did not influence the key policy messages. |
| Vosti 2023 | Vitamin A | Edible oils | 12 mg/kg | NR | No fortification | Cameroon | US$ | 2019 | Child lives saved | 13,428 | Total program costs US $ 7,306,000 | 544 | 558 | NR | NR | The program selection of "high" or "low" effectiveness results did not influence the |

| Study ID* (Author Year) | INTERVENTION-Micronutrient(s) | INTERVENTION-Food Vehicle(s) | Dose | Other Specs | Compar-ator | Country | Curr-ency | Price Year | HEALTH Outcome Type | Health Result - Difference | Costs - Difference *as REPORTED* | Incremental CE Ratio *as REPORTED* | CONVERTED ICER (using 2022 US$)† | Cost-Benefit Analysis | Dis-count Rate | Uncertainty & Sensitivity Analyses |
|---|---|---|---|---|---|---|---|---|---|---|---|---|---|---|---|---|
| | | | | | | | | | | | | | | | | key policy messages. |
| Adams 2022/ Vosti 2023 | Vitamin A | Refined oil + Bouillon cube | Refined oil (12 mg/kg) + bouillon (80 mg/kg) | NR | No fortification /interventio n | Cameroon | US$ | 2019 | Effective coverage: those at risk of deficiency (inadequate intake) who then received sufficient additional intake from intervention | 7,812 | 15,740 | $ 2.01 per child-year effectively covered | 2 | NR | NR | NR |
| Adams 2022/ Vosti 2023 | Vitamin A | Wheat flour | 5.9 mg/kg | NR | No fortification /interventio n | Cameroon | US$ | 2019 | Effective coverage: those at risk of deficiency (inadequate intake) who then received sufficient additional intake from intervention | 4,142 | 22,243 | $ 5.37 per child-year effectively covered | 6 | NR | NR | NR |
| Fiedler 2009 | Vitamin A | Sugar | 7.5-20 mg retinol (RE)/kg | Retinyl palmita te; Industr ial | Pre-fortification | Chad | US$ | 2009 | DALY saved | NR | 15.47 million | 107 | 107 | NR | 3% for DALY s; 0% for costs | NR |
| Asian Develop. Bank 2004 | Vitamin A | Oil | 50 IU (15 mcg) per gram | Mixing tank method . Retinol palmita te compo und | Pre-fortification | China | US$ | NR | Deaths averted | 14,000 | 67,364,000 | NR | N/A | BCR: 1.7 | NR [Disco unted costs and benefit s] | NR |
| Fiedler 2009 | Vitamin A | Sugar | 7.5-20 mg retinol (RE)/kg | Retinyl palmita te; Industr ial | Pre-fortification | Congo, Dem. Rep. | US$ | 2009 | DALY saved | NR | 43.10 million | 37 | 56 | NR | 3% for DALY s; 0% for costs | NR |

| Study ID* (Author Year) | INTERVENTION-Micronutrient(s) | INTERVENTION-Food Vehicle(s) | Dose | Other Specs | Compar-ator | Country | Curr-ency | Price Year | HEALTH Outcome Type | Health Result - Difference | Costs - Difference *as REPORTED* | Incremental CE Ratio *as REPORTED* | CONVERTED ICER (using 2022 US$)† | Cost-Benefit Analysis | Dis-count Rate | Uncertainty & Sensitivity Analyses |
|---|---|---|---|---|---|---|---|---|---|---|---|---|---|---|---|---|
| Fiedler 2009 | Vitamin A | Vegetable oil | 1.5-4.0 mg retinol (RE)/kg | Retinyl palmitate; Industrial | Pre-fortification | Congo, Dem. Rep. | US$ | 2009 | DALY saved | NR | 23.89 million | 43 | 65 | NR | 3% for DALYs; 0% for costs | NR |
| Fiedler 2009 | Vitamin A | Sugar | 7.5-20 mg retinol (RE)/kg | Retinyl palmitate; Industrial | Pre-fortification | Côte d'Ivoire | US$ | 2009 | DALY saved | NR | 93.89 million | 448 | 439 | NR | 3% for DALYs; 0% for costs | NR |
| Fiedler 2009 | Vitamin A | Vegetable oil | 1.5-4.0 mg retinol (RE)/kg | Retinyl palmitate; Industrial | Pre-fortification | Côte d'Ivoire | US$ | 2009 | DALY saved | NR | 6.94 million | 48 | 47 | NR | 3% for DALYs; 0% for costs | NR |
| Fiedler 2009 | Vitamin A | Sugar | 7.5-20 mg retinol (RE)/kg | Retinyl palmitate; Industrial | Pre-fortification | Egypt | US$ | 2009 | DALY saved | NR | 126.94 million | 399 | 525 | NR | 3% for DALYs; 0% for costs | NR |
| Fiedler 2009 | Vitamin A | Vegetable oil | 1.5-4.0 mg retinol (RE)/kg | Retinyl palmitate; Industrial | Pre-fortification | Egypt | US$ | 2009 | DALY saved | NR | 195.31 million | 667 | 878 | NR | 3% for DALYs; 0% for costs | NR |
| Fiedler 2009 | Vitamin A | Vegetable oil | 1.5-4.0 mg retinol (RE)/kg | Retinyl palmitate; Industrial | Pre-fortification | Ethiopia | US$ | 2009 | DALY saved | NR | 21.18 million | 43 | 59 | NR | 3% for DALYs; 0% for costs | NR |
| Moges 2020 | Vitamin A | Edible oil | 2000 mcg retinol/100 g oil | NR | Pre-fortification | Ethiopia | US$ | 2011 | Child deaths averted (among non-breastfed) | 3713-5071 | 3,657,327 | US$721-US$985 per child death averted | 1,175-1,605 | NR | NR | NR |
| Rajkumar 2012 | Vitamin A | Sugar | * | NR | Unfortified | Ethiopia | US$ | 2007 | Deaths averted | NR | Cost per beneficiary $0.10 | $ 102.48 per death averted | 175 | BCR: 15.32 | 5% | NR |
| Fiedler 2009 | Vitamin A | Vegetable oil | 1.5-4.0 mg retinol (RE)/kg | Retinyl palmitate; Industrial | Pre-fortification | Ghana | US$ | 2009 | DALY saved | NR | 42.01 million | 251 | 341 | NR | 3% for DALYs; 0% for costs | NR |
| Fiedler 2009 | Vitamin A | Vegetable oil | 1.5-4.0 mg retinol (RE)/kg | Retinyl palmitate; | Pre-fortification | Guatemala | US$ | 2009 | DALY saved | NR | 31.04 million | 913 | 1,494 | NR | 3% for DALYs; 0% | NR |

| Study ID* (Author Year) | INTERVENTION-Micronutrient(s) | INTERVENTION-Food Vehicle(s) | Dose | Other Specs | Compar-ator | Country | Curr-ency | Price Year | HEALTH Outcome Type | Health Result - Difference | Costs - Difference *as REPORTED* | Incremental CE Ratio *as REPORTED* | CONVERTED ICER (using 2022 US$)† | Cost-Benefit Analysis | Dis-count Rate | Uncertainty & Sensitivity Analyses |
|---|---|---|---|---|---|---|---|---|---|---|---|---|---|---|---|---|
| | | | | Industrial | | | | | | | | | | | for costs | |
| Phillips 1996 | Vitamin A | Sugar | Per gram sugar: 4.71 mcg vitamin A in urban areas; 2.51 mcg vitamin A in rural areas | NR | Pre-fortification | Guatemala | US$ | 1991 | Number of person years of vitamin A gap reduction per year for the high risk population | 664,069 | NR | $ 3.583 per high-risk person year of adequacy achieved | 12 | NR | 10% | NR |
| Phillips 1996 | Vitamin A | Sugar | Per gram sugar: 7.3 mcg vitamin A in urban areas; 6.0 mcg vitamin A in rural areas | NR | Pre-fortification | Guatemala | US$ | 1991 | Number of person years of vitamin A gap reduction per year for the high risk population | 2,418,362 | 2,379,278 annually | $ 0.984 per high-risk person year of adequacy achieved | 3 | NR | 10% | NR |
| Phillips 1996 | Vitamin A | Sugar | Per gram sugar: 15 mcg vitamin A | NR | Pre-fortification | Guatemala | US$ | 1991 | Number of person years of vitamin A gap reduction per year for the high risk population | 2,418,362 | NR | $ 0.984 per high-risk person year of adequacy achieved | 3 | NR | 10% | NR |
| Fiedler 2009 | Vitamin A | Sugar | 7.5-20 mg retinol (RE)/kg | Retinyl palmitate; Industrial | Pre-fortification | Guinea | US$ | 2009 | DALY saved | NR | 15.47 million | 150 | 167 | NR | 3% for DALYs; 0% for costs | NR |
| Fiedler 2009 | Vitamin A | Vegetable oil | 1.5-4.0 mg retinol (RE)/kg | Retinyl palmitate; Industrial | Pre-fortification | Guinea | US$ | 2009 | DALY saved | NR | 17.64 million | 134 | 150 | NR | 3% for DALYs; 0% for costs | NR |
| Chow 2010 | Vitamin A | Mustard oil | NR | Bottling (vitamin A) in opaque | No fortification | India | US$ | 2005 | DALY averted | 10.8 million (95% CI, 10.5-11.1 million) | 3,177 million | NR [calculated as: $294/DALY averted] | 457 | Internal rate of return, IRR: 6% (95% CI, 5-7%) | 3% for both DALYs and costs | Varying mortality rate reduction with an optimistic rate (high efficacy) gave |

| Study ID* (Author Year) | INTERVENTION-Micronutrient(s) | INTERVENTION-Food Vehicle(s) | Dose | Other Specs | Compar-ator | Country | Curr-ency | Price Year | HEALTH Outcome Type | Health Result - Difference | Costs - Difference *as REPORTED* | Incremental CE Ratio *as REPORTED* | CONVERTED ICER (using 2022 US$)† | Cost-Benefit Analysis | Dis-count Rate | Uncertainty & Sensitivity Analyses |
|---|---|---|---|---|---|---|---|---|---|---|---|---|---|---|---|---|
| | | | | containers of oil mustard | | | | | | | | | | | | IRR= 22% (95% CI: 21-22). |
| Fiedler 2009 | Vitamin A | Sugar | 7.5-20 mg retinol (RE)/kg | Retinyl palmitate; Industrial | Pre-fortification | India | US$ | 2009 | DALY saved | NR | 14092.43 million | 1803 | 2,271 | NR | 3% for DALYs; 0% for costs | NR |
| Fiedler 2009 | Vitamin A | Vegetable oil | 1.5-4.0 mg retinol (RE)/kg | Retinyl palmitate; Industrial | Pre-fortification | India | US$ | 2009 | DALY saved | NR | 1814.76 million | 218 | 275 | NR | 3% for DALYs; 0% for costs | NR |
| Fiedler 2009 | Vitamin A | Sugar | 7.5-20 mg retinol (RE)/kg | Retinyl palmitate; Industrial | Pre-fortification | Indonesia | US$ | 2009 | DALY saved | NR | 692.12 million | 1212 | 1,635 | NR | 3% for DALYs; 0% for costs | NR |
| Fiedler 2009 | Vitamin A | Vegetable oil | 1.5-4.0 mg retinol (RE)/kg | Retinyl palmitate; Industrial | Pre-fortification | Indonesia | US$ | 2009 | DALY saved | NR | 241.11 million | 427 | 576 | NR | 3% for DALYs; 0% for costs | NR |
| Asian Develop. Bank 2004 | Vitamin A | Oil (Palm oil) | 50 IU (15 mcg) per gram | Mixing tank method. Retinol palmitate compound | Pre-fortification | Indonesia | US$ | NR | Deaths averted | 44,000 | 60,372,000 | NR | N/A | BCR: 11.0 | NR [Discounted costs and benefits] | NR |
| Fiedler 2009 | Vitamin A | Sugar | 7.5-20 mg retinol (RE)/kg | Retinyl palmitate; Industrial | Pre-fortification | Kenya | US$ | 2009 | DALY saved | NR | 61.75 million | 262 | 382 | NR | 3% for DALYs; 0% for costs | NR |
| Fiedler 2009 | Vitamin A | Vegetable oil | 1.5-4.0 mg retinol (RE)/kg | Retinyl palmitate; Industrial | Pre-fortification | Kenya | US$ | 2009 | DALY saved | NR | 28.36 million | 161 | 235 | NR | 3% for DALYs; 0% for costs | NR |
| Fiedler 2009 | Vitamin A | Sugar | 7.5-20 mg retinol (RE)/kg | Retinyl palmitate; | Pre-fortification | Madagascar | US$ | 2009 | DALY saved | NR | 47.83 million | 235 | 274 | NR | 3% for DALYs; 0% | NR |

| Study ID* (Author Year) | INTERVENTION-Micronutrient(s) | INTERVENTION-Food Vehicle(s) | Dose | Other Specs | Compar-ator | Country | Curr-ency | Price Year | HEALTH Outcome Type | Health Result - Difference | Costs - Difference *as REPORTED* | Incremental CE Ratio *as REPORTED* | CONVERTED ICER (using 2022 US$)† | Cost-Benefit Analysis | Dis-count Rate | Uncertainty & Sensitivity Analyses |
|---|---|---|---|---|---|---|---|---|---|---|---|---|---|---|---|---|
| | | | | Industrial | | | | | | | | | | | for costs | |
| Fiedler 2009 | Vitamin A | Vegetable oil | 1.5-4.0 mg retinol (RE)/kg | Retinyl palmitate; Industrial | Pre-fortification | Madagascar | US$ | 2009 | DALY saved | NR | 24.64 million | 98 | 114 | NR | 3% for DALYs; 0% for costs | NR |
| Fiedler 2009 | Vitamin A | Sugar | 7.5-20 mg retinol (RE)/kg | Retinyl palmitate; Industrial | Pre-fortification | Malawi | US$ | 2009 | DALY saved | NR | 29.30 million | 223 | 193 | NR | 3% for DALYs; 0% for costs | NR |
| Fiedler 2009 | Vitamin A | Vegetable oil | 1.5-4.0 mg retinol (RE)/kg | Retinyl palmitate; Industrial | Pre-fortification | Malawi | US$ | 2009 | DALY saved | NR | 9.74 million | 105 | 91 | NR | 3% for DALYs; 0% for costs | NR |
| Fiedler 2009 | Vitamin A | Sugar | 7.5-20 mg retinol (RE)/kg | Retinyl palmitate; Industrial | Pre-fortification | Mali | US$ | 2009 | DALY saved | NR | 32.34 million | 146 | 166 | NR | 3% for DALYs; 0% for costs | NR |
| Fiedler 2009 | Vitamin A | Vegetable oil | 1.5-4.0 mg retinol (RE)/kg | Retinyl palmitate; Industrial | Pre-fortification | Mali | US$ | 2009 | DALY saved | NR | 222.25 million | 2696 | 3,065 | NR | 3% for DALYs; 0% for costs | NR |
| Fiedler 2009 | Vitamin A | Sugar | 7.5-20 mg retinol (RE)/kg | Retinyl palmitate; Industrial | Pre-fortification | Mexico | US$ | 2009 | DALY saved | NR | 377.56 million | 5194 | 6,383 | NR | 3% for DALYs; 0% for costs | NR |
| Fiedler 2009 | Vitamin A | Vegetable oil | 1.5-4.0 mg retinol (RE)/kg | Retinyl palmitate; Industrial | Pre-fortification | Mexico | US$ | 2009 | DALY saved | NR | 290.30 million | 4251 | 5,224 | NR | 3% for DALYs; 0% for costs | NR |
| Fiedler 2009 | Vitamin A | Sugar | 7.5-20 mg retinol (RE)/kg | Retinyl palmitate; Industrial | Pre-fortification | Morocco | US$ | 2009 | DALY saved | NR | 205.88 million | 2711 | 2,456 | NR | 3% for DALYs; 0% for costs | NR |
| Fiedler 2009 | Vitamin A | Sugar | 7.5-20 mg retinol (RE)/kg | Retinyl palmitate; Industrial | Pre-fortification | Mozambique | US$ | 2009 | DALY saved | NR | 124.50 million | 1545 | 1,269 | NR | 3% for DALYs; 0% for costs | NR |

| Study ID* (Author Year) | INTERVENTION-Micronutrient(s) | INTERVENTION-Food Vehicle(s) | Dose | Other Specs | Compar-ator | Country | Curr-ency | Price Year | HEALTH Outcome Type | Health Result - Difference | Costs - Difference *as REPORTED* | Incremental CE Ratio *as REPORTED* | CONVERTED ICER (using 2022 US$)† | Cost-Benefit Analysis | Dis-count Rate | Uncertainty & Sensitivity Analyses |
|---|---|---|---|---|---|---|---|---|---|---|---|---|---|---|---|---|
| Fiedler 2009 | Vitamin A | Vegetable oil | 1.5-4.0 mg retinol (RE)/kg | Retinyl palmitate; Industrial | Pre-fortification | Mozambique | US$ | 2009 | DALY saved | NR | 29.45 million | 301 | 247 | NR | 3% for DALYs; 0% for costs | NR |
| Fiedler 2009 | Vitamin A | Sugar | 7.5-20 mg retinol (RE)/kg | Retinyl palmitate; Industrial | Pre-fortification | Myanmar | US$ | 2009 | DALY saved | NR | 377.70 million | 1422 | 10 | NR | 3% for DALYs; 0% for costs | NR |
| Fiedler 2009 | Vitamin A | Vegetable oil | 1.5-4.0 mg retinol (RE)/kg | Retinyl palmitate; Industrial | Pre-fortification | Myanmar | US$ | 2009 | DALY saved | NR | 98.07 million | 318 | 2 | NR | 3% for DALYs; 0% for costs | NR |
| Fiedler 2009 | Vitamin A | Sugar | 7.5-20 mg retinol (RE)/kg | Retinyl palmitate; Industrial | Pre-fortification | Nepal | US$ | 2009 | DALY saved | NR | 126.57 million | 454 | 799 | NR | 3% for DALYs; 0% for costs | NR |
| Fiedler 2009 | Vitamin A | Vegetable oil | 1.5-4.0 mg retinol (RE)/kg | Retinyl palmitate; Industrial | Pre-fortification | Nepal | US$ | 2009 | DALY saved | NR | 11.30 million | 39 | 69 | NR | 3% for DALYs; 0% for costs | NR |
| Fiedler 2009 | Vitamin A | Sugar | 7.5-20 mg retinol (RE)/kg | Retinyl palmitate; Industrial | Pre-fortification | Niger | US$ | 2009 | DALY saved | NR | 15.27 million | 52 | 51 | NR | 3% for DALYs; 0% for costs | NR |
| Fiedler 2009 | Vitamin A | Vegetable oil | 1.5-4.0 mg retinol (RE)/kg | Retinyl palmitate; Industrial | Pre-fortification | Niger | US$ | 2009 | DALY saved | NR | 15.89 million | 47 | 46 | NR | 3% for DALYs; 0% for costs | NR |
| Fiedler 2009 | Vitamin A | Sugar | 7.5-20 mg retinol (RE)/kg | Retinyl palmitate; Industrial | Pre-fortification | Nigeria | US$ | 2009 | DALY saved | NR | 95.63 million | 50 | 54 | NR | 3% for DALYs; 0% for costs | NR |
| Fiedler 2009 | Vitamin A | Vegetable oil | 1.5-4.0 mg retinol (RE)/kg | Retinyl palmitate; Industrial | Pre-fortification | Nigeria | US$ | 2009 | DALY saved | NR | 179.15 million | 81 | 88 | NR | 3% for DALYs; 0% for costs | NR |

| Study ID* (Author Year) | INTERVENTION-Micronutrient(s) | INTERVENTION-Food Vehicle(s) | Dose | Other Specs | Compar-ator | Country | Curr-ency | Price Year | HEALTH Outcome Type | Health Result - Difference | Costs - Difference *as REPORTED* | Incremental CE Ratio *as REPORTED* | CONVERTED ICER (using 2022 US$)† | Cost-Benefit Analysis | Dis-count Rate | Uncertainty & Sensitivity Analyses |
|---|---|---|---|---|---|---|---|---|---|---|---|---|---|---|---|---|
| Fiedler 2009 | Vitamin A | Sugar | 7.5-20 mg retinol (RE)/kg | Retinyl palmitate; Industrial | Pre-fortification | Pakistan | US$ | 2009 | DALY saved | NR | 880.75 million | 616 | 687 | NR | 3% for DALYs; 0% for costs | NR |
| Fiedler 2009 | Vitamin A | Vegetable oil | 1.5-4.0 mg retinol (RE)/kg | Retinyl palmitate; Industrial | Pre-fortification | Pakistan | US$ | 2009 | DALY saved | NR | 379.87 million | 289 | 322 | NR | 3% for DALYs; 0% for costs | NR |
| Asian Develop. Bank 2004 | Vitamin A | Oil (Vegetable ghee and Edible oil) | NR | Mixing tank method. Retinol palmitate compound | Pre-fortification | Pakistan | US$ | NR | Deaths averted | 42,000 | 35,833,000 | NR | N/A | BCR: 3.9 | NR [Discounted costs and benefits] | NR |
| Fiedler 2009 | Vitamin A | Sugar | 7.5-20 mg retinol (RE)/kg | Retinyl palmitate; Industrial | Pre-fortification | Peru | US$ | 2009 | DALY saved | NR | 111.44 million | 985 | 1,229 | NR | 3% for DALYs; 0% for costs | NR |
| Fiedler 2009 | Vitamin A | Vegetable oil | 1.5-4.0 mg retinol (RE)/kg | Retinyl palmitate; Industrial | Pre-fortification | Peru | US$ | 2009 | DALY saved | NR | 79.52 million | 1528 | 1,907 | NR | 3% for DALYs; 0% for costs | NR |
| Fiedler 2009 | Vitamin A | Sugar | 7.5-20 mg retinol (RE)/kg | Retinyl palmitate; Industrial | Pre-fortification | Philippines | US$ | 2009 | DALY saved | NR | 315.10 million | 980 | 1,176 | NR | 3% for DALYs; 0% for costs | NR |
| Fiedler 2009 | Vitamin A | Vegetable oil | 1.5-4.0 mg retinol (RE)/kg | Retinyl palmitate; Industrial | Pre-fortification | Philippines | US$ | 2009 | DALY saved | NR | 111.41 million | 350 | 420 | NR | 3% for DALYs; 0% for costs | NR |
| Popkin 1980 | Vitamin A | MSG (in rural coastal barrios) | 15,000 IU per 2.4 mg packet | Vitamin A combined with MSG in a large mixer | Pre-fortification | Philippines | US$ | 1980 | Benefit-cost ratio | NR | NR | NR | N/A | BCR: 3.8 | 15% | Varying discount rate to 8%, gives BCR: 8.4 |

| Study ID* (Author Year) | INTERVENTION-Micronutrient(s) | INTERVENTION-Food Vehicle(s) | Dose | Other Specs | Compar-ator | Country | Curr-ency | Price Year | HEALTH Outcome Type | Health Result - Difference | Costs - Difference *as REPORTED* | Incremental CE Ratio *as REPORTED* | CONVERTED ICER (using 2022 US$)† | Cost-Benefit Analysis | Dis-count Rate | Uncertainty & Sensitivity Analyses |
|---|---|---|---|---|---|---|---|---|---|---|---|---|---|---|---|---|
| | | | | along with silicic acid, a flow-enhancing agent. | | | | | | | | | | | | |
| Popkin 1980 | Vitamin A | MSG (in urban squatter areas) | 15,000 IU per 2.4 mg packet | Vitamin A combined with MSG in a large mixer along with silicic acid, a flow-enhancing agent. | Pre-fortification | Philippines | US$ | 1980 | Benefit-cost ratio | NR | NR | NR | N/A | BCR: 2.4 | 15% | Varying discount rate to 8%, gives BCR: 6.2 |
| Fiedler 2000 | Vitamin A | All wheat flour | 490 REs/100g flour | retinol palmitate type 250 SD | Unfortified flour | Philippines | Philippine pesos | 1998 | Vitamin A (VA) gap reduction per year | 1,028,000 person years | P 227,195,000 | Cost per person-year of adequate VA intake achieved: 221 | 10 | NR | NR | NR |
| Fiedler 2000 | Vitamin A | Hard wheat flour | 490 REs/100g flour | retinol palmitate type 250 SD | Unfortified flour | Philippines | Philippine pesos | 1998 | Vitamin A (VA) gap reduction per year | 982,000 person years | P 149,244,000 | Cost per person-year of adequate VA intake achieved: 152 | 7 | NR | NR | NR |
| Fiedler 2000 | Vitamin A | Hard wheat flour | 710 REs/100g flour | retinol palmitate type 250 SD | Unfortified flour | Philippines | Philippine pesos | 1998 | Vitamin A (VA) gap reduction per year | 1,687,000 person years | P 221,465,000 | Cost per person-year of adequate VA intake achieved: 131 | 6 | NR | NR | With a dose of 600 REs/100 g in hard flour resulted in: P 141 per person-year of adequate VA intake achieved. |
| Fiedler 2000 | Vitamin A | Hard wheat flour | 820 REs/100g flour | retinol palmitate type 250 SD | Unfortified flour | Philippines | Philippine pesos | 1998 | Vitamin A (VA) gap reduction per year | 1,890,000 person years | P 270,143,000 | Cost per person-year of adequate VA intake achieved: 143 | 6 | NR | NR | NR |
| Fiedler 2009 | Vitamin A | Sugar | 7.5-20 mg retinol (RE)/kg | Retinyl palmitate; Industrial | Pre-fortification | Rwanda | US$ | 2009 | DALY saved | NR | 29.29 million | 225 | 228 | NR | 3% for DALYs; 0% for costs | NR |

| Study ID* (Author Year) | INTERVENTION-Micronutrient(s) | INTERVENTION-Food Vehicle(s) | Dose | Other Specs | Compar-ator | Country | Curr-ency | Price Year | HEALTH Outcome Type | Health Result - Difference | Costs - Difference *as REPORTED* | Incremental CE Ratio *as REPORTED* | CONVERTED ICER (using 2022 US$)† | Cost-Benefit Analysis | Dis-count Rate | Uncertainty & Sensitivity Analyses |
|---|---|---|---|---|---|---|---|---|---|---|---|---|---|---|---|---|
| Fiedler 2009 | Vitamin A | Vegetable oil | 1.5-4.0 mg retinol (RE)/kg | Retinyl palmitate; Industrial | Pre-fortification | Rwanda | US$ | 2009 | DALY saved | NR | 6.62 million | 68 | 69 | NR | 3% for DALYs; 0% for costs | NR |
| Edejer 2005 | Vitamin A | Sugar | 80% coverage; dose NR | NR | No fortification | Sear-D (South East Asia) region | international dollars ($Int) | 2000 | DALYs averted yearly (millions) | 0.10 | 25,000,000 yearly | ICER $ 244/DALY saved | 366 | NR | 3% for both costs and benefits | Removal of age weighting and discounting for DALYs makes the interventions more cost effective. Varying geographical coverage from 80% to: 50% or 95% gave ICERs of I$ 277 or 237 per DALY saved (resp'ly). |
| Fiedler 2009 | Vitamin A | Vegetable oil | 1.5-4.0 mg retinol (RE)/kg | Retinyl palmitate; Industrial | Pre-fortification | Sierra Leone | US$ | 2009 | DALY saved | NR | 10.60 million | 69 | 70 | NR | 3% for DALYs; 0% for costs | NR |
| Fiedler 2009 | Vitamin A | Sugar | 7.5-20 mg retinol (RE)/kg | Retinyl palmitate; Industrial | Pre-fortification | South Africa | US$ | 2009 | DALY saved | NR | 174.67 million | 933 | 962 | NR | 3% for DALYs; 0% for costs | NR |
| Fiedler 2009 | Vitamin A | Vegetable oil | 1.5-4.0 mg retinol (RE)/kg | Retinyl palmitate; Industrial | Pre-fortification | South Africa | US$ | 2009 | DALY saved | NR | 113.09 million | 734 | 756 | NR | 3% for DALYs; 0% for costs | NR |
| van Stuijvenberg 2001 | Vitamin A | Biscuit | 1.17 mg per 45 g serving | synthetic beta-carotene | Unfortified biscuit | South Africa | US$ | 1999 | Decrease in prevalence of low serum retinol level (<15 mcg/dl) | 8.7% | 0.61 per child per school year | NR | N/A | NR | NA | NR |
| Fiedler 2009 | Vitamin A | Sugar | 7.5-20 mg retinol (RE)/kg | Retinyl palmitate; Industrial | Pre-fortification | Sudan | US$ | 2009 | DALY saved | NR | 47.83 million | 613 | 656 | NR | 3% for DALYs; 0% for costs | NR |

| Study ID* (Author Year) | INTERVENTION-Micronutrient(s) | INTERVENTION-Food Vehicle(s) | Dose | Other Specs | Compar-ator | Country | Curr-ency | Price Year | HEALTH Outcome Type | Health Result - Difference | Costs - Difference *as REPORTED* | Incremental CE Ratio *as REPORTED* | CONVERTED ICER (using 2022 US$)† | Cost-Benefit Analysis | Dis-count Rate | Uncertainty & Sensitivity Analyses |
|---|---|---|---|---|---|---|---|---|---|---|---|---|---|---|---|---|
| Fiedler 2009 | Vitamin A | Vegetable oil | 1.5-4.0 mg retinol (RE)/kg | Retinyl palmitate; Industrial | Pre-fortification | Sudan | US$ | 2009 | DALY saved | NR | 50.94 million | 496 | 531 | NR | 3% for DALYs; 0% for costs | NR |
| Fiedler 2009 | Vitamin A | Sugar | 7.5-20 mg retinol (RE)/kg | Retinyl palmitate; Industrial | Pre-fortification | Tanzania | US$ | 2009 | DALY saved | NR | 69.47 million | 227 | 281 | NR | 3% for DALYs; 0% for costs | NR |
| Fiedler 2009 | Vitamin A | Vegetable oil | 1.5-4.0 mg retinol (RE)/kg | Retinyl palmitate; Industrial | Pre-fortification | Tanzania | US$ | 2009 | DALY saved | NR | 49.41 million | 106 | 131 | NR | 3% for DALYs; 0% for costs | NR |
| Walters 2019 | Vitamin A | Sunflower oil (by large-scale producer) | 36.6–73.3 mg/kg retinol (equivalent to 20–40 mg/kg of retinyl palmitate) | | unfortified oil | Tanzania | US$ | 2017 | DALYs averted per year | 26,105 nationally (including all scales of producers) | 0.13 per capita per year | ICER USD 281/DALY averted | 314 | NR | NR | Using a high estimate of VAD prevalence reduction (instead of conservative as extracted) would result in 30,996 DALYs averted per year (nationally across all producers). |
| Walters 2019 | Vitamin A | Sunflower oil (by medium-scale producer) | 36.6–73.3 mg/kg retinol (equivalent to 20–40 mg/kg of retinyl palmitate) | | unfortified oil | Tanzania | US$ | 2017 | DALYs averted per year | 26,105 nationally (including all scales of producers) | 1.36 per capita per year | ICER USD 2,892/DALY averted | 3,229 | NR | NR | If enable ideal enterprise conditions and government policy changes, then ICER estimate is much lower: $ 626 per DALY averted for medium-scale producers. |
| Walters 2019 | Vitamin A | Sunflower oil (by small-scale producer) | 36.6–73.3 mg/kg retinol (equivalent to 20–40 mg/kg of retinyl palmitate) | | unfortified oil | Tanzania | US$ | 2017 | DALYs averted per year | 26,105 nationally (including all scales of producers) | 2.09 per capita per year | ICER USD 4,442/DALY averted | 4,960 | NR | NR | If enable ideal enterprise conditions and government policy changes, then ICER estimate is much lower: $ 1,507 per DALY averted for small-scale producers. |
| Fiedler 2010 | Vitamin A | Sugar | 15 mg/kg | Retinol palmitate | Unfortified food | Uganda | US$ | 2008 | DALYs averted | NR | 2,644,765 annually | 82 | 127 | NR | Discounted | NR |

| Study ID* (Author Year) | INTERVENTION-Micronutrient(s) | INTERVENTION-Food Vehicle(s) | Dose | Other Specs | Compar-ator | Country | Curr-ency | Price Year | HEALTH Outcome Type | Health Result - Difference | Costs - Difference *as REPORTED* | Incremental CE Ratio *as REPORTED* | CONVERTED ICER (using 2022 US$)† | Cost-Benefit Analysis | Dis-count Rate | Uncertainty & Sensitivity Analyses |
|---|---|---|---|---|---|---|---|---|---|---|---|---|---|---|---|---|
| | | | | 250,000 IU/g | | | | | | | | | | | (details NR) | |
| Fiedler 2010 | Vitamin A | Vegetable oil | 35 mg/kg | Retinol palmitate 1,000,000 IU/g | Unfortified food | Uganda | US$ | 2008 | DALYs averted | NR | 555,668 annually | 18 | 28 | NR | Discounted (details NR) | NR |
| Fiedler 2009 | Vitamin A | Vegetable oil | 1.5-4.0 mg retinol (RE)/kg | Retinyl palmitate; Industrial | Pre-fortification | Uzbekistan | US$ | 2009 | DALY saved | NR | 102.59 million | 966 | 1,283 | NR | 3% for DALYs; 0% for costs | NR |
| Fiedler 2009 | Vitamin A | Sugar | 7.5-20 mg retinol (RE)/kg | Retinyl palmitate; Industrial | Pre-fortification | Vietnam | US$ | 2009 | DALY saved | NR | 503.81 million | 6219 | 11,133 | NR | 3% for DALYs; 0% for costs | NR |
| Fiedler 2009 | Vitamin A | Vegetable oil | 1.5-4.0 mg retinol (RE)/kg | Retinyl palmitate; Industrial | Pre-fortification | Vietnam | US$ | 2009 | DALY saved | NR | 43.43 million | 514 | 920 | NR | 3% for DALYs; 0% for costs | NR |
| Sablah 2012 | vitamin A | Cooking oil | Unclear (Mali 25 IU/g, Côte d'Ivoire 4 mcg/g, both as retinyl palmitate) | NR | Pre-fortification | West African Economic and Monetary Union (UEMOA): Benin, Burkina Faso, Côte d'Ivoire, Guinea-Bissau, Mali, Niger, Senegal, Togo | US$ | NR | DALYs saved per year | 740,000 | NR | $3.79/DALY saved | 4 | NR | NR | NR |
| Fiedler 2009 | Vitamin A | Vegetable oil | 1.5-4.0 mg retinol (RE)/kg | Retinyl palmitate; Industrial | Pre-fortification | Yemen | US$ | 2009 | DALY saved | NR | 48.17 million | 215 | 341 | NR | 3% for DALYs; 0% for costs | NR |
| Fiedler 2014 | Vitamin A | Maize meal | 1 mcg/g Retinol Activity Equivalent (RAE) | 1 IU retinol = 0.3003 mcg retinol. | Unfortified food | Zambia | US$ | 2013 | DALYs saved | 400,000 [roughly extracted from a chart] | NR | ICER $ 130/DALY saved [roughly extracted from a chart] | 102 | NR | 3% for both costs and DALYs | Using the year 2013 instead of 30-year total found a slightly lower ICER, as did using 10- or 20-year |

| Study ID* (Author Year) | INTERVENTION-Micronutrient(s) | INTERVENTION-Food Vehicle(s) | Dose | Other Specs | Compar-ator | Country | Curr-ency | Price Year | HEALTH Outcome Type | Health Result - Difference | Costs - Difference *as REPORTED* | Incremental CE Ratio *as REPORTED* | CONVERTED ICER (using 2022 US$)† | Cost-Benefit Analysis | Dis-count Rate | Uncertainty & Sensitivity Analyses |
|---|---|---|---|---|---|---|---|---|---|---|---|---|---|---|---|---|
| | | | | Maize as breakfast meal and roller meal. | | | | | | | | | | | | accounting periods instead. |
| Fiedler 2014 | Vitamin A | Sugar | 10 mcg/g RAE | 1 IU retinol = 0.3003 mcg retinol | Unfortified food | Zambia | US$ | 2013 | DALYs saved | 1,400,000 [roughly extracted from a chart] | Approx. 23,000,000 [roughly extracted from a chart] | ICER $ 19/DALY saved [roughly extracted from a chart] | 15 | NR | 3% for both costs and DALYs | Using the year 2013 instead of 30-year total found similar ICERs, as did using 10- or 20-year accounting periods. |
| Fiedler 2013 | Vitamin A | Sugar | Vitamin A (palmitate 250 CWS/CWD) minimum 10 mg /kg | NR | Unfortified food | Zambia | US$ | 2006 | DALYs saved | 7,337 | 873,065 per year | ICER $ 119/ DALY saved | 122 | NR | NR | Changes in 'prevalence' of inadequate intakes used to calculate DALYs lost were extracted here. In all instances, "efficiency-based" estimates were more cost-effective than prevalence-based ICERs. |
| Fiedler 2009 | Vitamin A | Sugar | 7.5-20 mg retinol (RE)/kg | Retinyl palmitate; Industrial | Pre-fortification | Zambia | US$ | 2009 | DALY saved | NR | 29.32 million | 218 | 237 | NR | 3% for DALYs; 0% for costs | NR |
| Fiedler 2014 | Vitamin A | Sugar & Vegetable oil | mcg/g RAE = 10 (Sugar), mcg/g RAE = 30 (Vegetable oil) | 1 IU retinol = 0.3003 mcg retinol | Unfortified food | Zambia | US$ | 2013 | DALYs saved | 1,900,000 [roughly extracted from a chart] | Approx. 30,000,000 [roughly extracted from a chart] | ICER $ 18/DALY saved [roughly extracted from a chart] | 14 | NR | 3% for both costs and DALYs | NR |
| Fiedler 2014 | Vitamin A | Sugar & Vegetable oil & Wheat flour | mcg/g RAE = 10 (Sugar), mcg/g RAE = 30 (Vegetable oil), mcg/g RAE = 5.9 (Wheat flour) | 1 IU retinol = 0.3003 mcg retinol | Unfortified food | Zambia | US$ | 2013 | DALYs saved | NR | NR | ICER $ 32/DALY saved [roughly extracted from a chart] | 25 | NR | 3% for both costs and DALYs | NR |

| Study ID* (Author Year) | INTERVENTION- Micronutrient(s) | INTERVENTION- Food Vehicle(s) | Dose | Other Specs | Compar- ator | Country | Curr- ency | Price Year | HEALTH Outcome Type | Health Result - Difference | Costs - Difference *as REPORTED* | Incremental CE Ratio *as REPORTED* | CONVERTED ICER (using 2022 US$)† | Cost- Benefit Analysis | Dis- count Rate | Uncertainty & Sensitivity Analyses |
|---|---|---|---|---|---|---|---|---|---|---|---|---|---|---|---|---|
| Fiedler 2014 | Vitamin A | Sugar & Wheat flour | mcg/g RAE = 10 (Sugar), mcg/g RAE = 5.9 (Wheat flour) | 1 IU retinol = 0.3003 mcg retinol | Unfortified food | Zambia | US$ | 2013 | DALYs saved | NR | NR | ICER $ 37/DALY saved [roughly extracted from a chart] | 29 | NR | 3% for both costs and DALYs | NR |
| Fiedler 2013 | Vitamin A | Sugar + Vegetable oil | Sugar: Vitamin A (palmitate 250 CWS/CWD) minimum 10 mg /kg; Vegetable oil: Vitamin A (retinol) 30 mg/kg | NR | Unfortified food | Zambia | US$ | 2006 | DALYs saved | 26,216 | 1,115,465 per year | ICER $ 43/ DALY saved | 44 | NR | NR | Changes in 'prevalence' of inadequate intakes used to calculate DALYs lost were extracted here. In all instances, "efficiency-based" estimates were more cost-effective than prevalence-based ICERs. |
| Fiedler 2014 | Vitamin A | Vegetable oil | 30 mcg/g RAE | 1 IU retinol = 0.3003 mcg retinol | Unfortified food | Zambia | US$ | 2013 | DALYs saved | 1,700,000 [roughly extracted from a chart] | Approx. 6,000,000 [roughly extracted from a chart] | ICER $ 5/DALY saved [roughly extracted from a chart] | 4 | NR | 3% for both costs and DALYs | Using the year 2013 instead of 30-year total found similar ICERs, as did using 10- or 20-year accounting periods. |
| Fiedler 2013 | Vitamin A | Vegetable oil | Vitamin A (retinol) 30 mg/kg | NR | Unfortified food | Zambia | US$ | 2006 | DALYs saved | 17,170 | 242,400 per year | ICER $ 14/ DALY saved | 14 | NR | NR | Changes in 'prevalence' of inadequate intakes used to calculate DALYs lost were extracted here. In all instances, "efficiency-based" estimates were more cost-effective than prevalence-based ICERs. |
| Fiedler 2014 | Vitamin A | Wheat flour | 5.9 mcg/g RAE | 1 IU retinol = 0.3003 mcg retinol | Unfortified food | Zambia | US$ | 2013 | DALYs saved | 500,000 [roughly extracted from a chart] | NR | ICER $ 35/DALY saved [roughly extracted from a chart] | 27 | NR | 3% for both costs and DALYs | Using the year 2013 instead of 30-year total found similar ICERs, as did using 10- or 20-year accounting periods. |
| Fiedler 2014 | Vitamin A | Wheat flour & Vegetable oil | mcg/g RAE = 5.9 (Wheat flour), | 1 IU retinol = 0.3003 | Unfortified food | Zambia | US$ | 2013 | DALYs saved | NR | NR | ICER $ 22/DALY saved [roughly | 17 | NR | 3% for both costs and | NR |

| Study ID* (Author Year) | INTERVENTION-Micronutrient(s) | INTERVENTION-Food Vehicle(s) | Dose | Other Specs | Compar-ator | Country | Curr-ency | Price Year | HEALTH Outcome Type | Health Result - Difference | Costs - Difference *as REPORTED* | Incremental CE Ratio *as REPORTED* | CONVERTED ICER (using 2022 US$)† | Cost-Benefit Analysis | Dis-count Rate | Uncertainty & Sensitivity Analyses |
|---|---|---|---|---|---|---|---|---|---|---|---|---|---|---|---|---|
| | | | mcg/g RAE = 30 (Vegetable oil) | mcg retinol | | | | | | | | extracted from a chart] | | | DALYs | |
| Fiedler 2009 | Vitamin A | Sugar | 7.5-20 mg retinol (RE)/kg | Retinyl palmitate; Industrial | Pre-fortification | Zimbabwe | US$ | 2009 | DALY saved | NR | 103.27 million | 33 | 39 | NR | 3% for DALYs; 0% for costs | NR |
| Fiedler 2009 | Vitamin A | Vegetable oil | 1.5-4.0 mg retinol (RE)/kg | Retinyl palmitate; Industrial | Pre-fortification | Zimbabwe | US$ | 2009 | DALY saved | NR | 24.39 million | 487 | 570 | NR | 3% for DALYs; 0% for costs | NR |
| Horton 2003 | Vitamins A (maize flour only) + B1 + B2 + B3 + Iron | Maize and wheat flour | NR | NR | No mandatory fortification | Bangladesh, India, Mali, Tanzania, Egypt, Oman, Bolivia, Honduras, Nicaragua (individually) | US$ | 1994 | Benefit-cost ratio | NR | 0.12 per person annually | NR | N/A | BCR: median 8.7 (for all modeled countries, BCR was >1) | 3% | If cognitive improvements are excluded, the model using physical productivity alone gives a median BCR of value is 6:1 among the study countries. |
| Fiedler 2015 | Vitamins A + B1 + B2 + B3 + B6 + B12 + Folic acid + Calcium + Iron + Zinc | Wheat flour | Fe: 55 mg/kg + Zn: 27 mg/kg + Vit. A: 4 IU/g + Ca: 53 mg/kg + Vit. B1: 6 mg/kg + Vit. B2: 4 mg/kg + Vit. B3: 15 mg/kg + Vit. B6: 5 mg/kg + Folic acid: 2 mg/kg + Vit. B12: 0.010 mg/kg | Iron as NaFeEDTA; + Zinc oxide + Vit. A (dry) 250,000 IU/g + Di-Calcium phosphate 2H2O + Thiamine mononitrate + Riboflavin + Niacinamide | No fortification | Bangladesh | US$ | 2012 (unclear) | DALYs saved | 137,461 | 2,548,044 annually | ICER $ 18.54/ DALY saved | 33 | NR | Unclear (costs & benefits were discounted) | NR |

| Study ID* (Author Year) | INTERVENTION-Micronutrient(s) | INTERVENTION-Food Vehicle(s) | Dose | Other Specs | Compar-ator | Country | Curr-ency | Price Year | HEALTH Outcome Type | Health Result - Difference | Costs - Difference *as REPORTED* | Incremental CE Ratio *as REPORTED* | CONVERTED ICER (using 2022 US$)† | Cost-Benefit Analysis | Dis-count Rate | Uncertainty & Sensitivity Analyses |
|---|---|---|---|---|---|---|---|---|---|---|---|---|---|---|---|---|
| | | | | + Pyridoxine hydrochloride + Folic acid + Vit. B12 0.1% water soluble | | | | | | | | | | | | |
| Fiedler 2015 | Vitamins A + B1 + B2 + B3 + B6 + B12 + Folic acid + Calcium + Iron + Zinc | Wheat flour | Fe: 55 mg/kg + Zn: 27 mg/kg + Vit. A: 4 IU/g + Ca: 53 mg/kg + Vit. B1: 6 mg/kg + Vit. B2: 4 mg/kg + Vit. B3: 15 mg/kg + Vit. B6: 5 mg/kg + Folic acid: 2 mg/kg + Vit. B12: 0.010 mg/kg | Iron as Ferrous fumarate; + Zinc oxide + Vit. A (dry) 250,000 IU/g + Di-Calcium phosphate 2H2O + Thiamine mononitrate + Riboflavin + Niacinamide + Pyridoxine hydrochloride + Folic acid + Vit. B12 0.1% | No fortification | Bangladesh | US$ | 2012 (unclear) | DALYs saved | 129,212 | 1,905,369 annually | ICER $ 14.75/ DALY saved | 26 | NR | Unclear (costs & benefits were discounted) | NR |

| Study ID* (Author Year) | INTERVENTION-Micronutrient(s) | INTERVENTION-Food Vehicle(s) | Dose | Other Specs | Compar-ator | Country | Curr-ency | Price Year | HEALTH Outcome Type | Health Result - Difference | Costs - Difference *as REPORTED* | Incremental CE Ratio *as REPORTED* | CONVERTED ICER (using 2022 US$)† | Cost-Benefit Analysis | Dis-count Rate | Uncertainty & Sensitivity Analyses |
|---|---|---|---|---|---|---|---|---|---|---|---|---|---|---|---|---|
| | | | | water soluble | | | | | | | | | | | | |
| Fiedler 2015 | Vitamins A + B1 + B2 + B3 + B6 + B12 + Folic acid + Calcium + Iron + Zinc | Wheat flour + Vegetable oil | Fe: 55 mg/kg + Zn: 27 mg/kg + Vit. A: flour (4 IU/g) & oil (15 mg/kg); + Ca: 53 mg/kg + Vit. B1: 6 mg/kg + Vit. B2: 4 mg/kg + Vit. B3: 15 mg/kg + Vit. B6: 5 mg/kg + Folic acid: 2 mg/kg + Vit. B12: 0.010 mg/kg | Iron as NaFeEDTA; + Zinc oxide + Vit. A: flour (250,000 IU/g, dry) & oil (retinyl palmitate 1.7 mIU/g); + Di-Calcium phosphate 2H2O + Thiamine mononitrate + Riboflavin + Niacinamide + Pyridoxine hydrochloride + Folic acid + Vit. B12 0.1% water soluble | No fortification | Bangladesh | US$ | 2012 (unclear) | DALYs saved | 479,898 | 3,816,160 annually | ICER $ 7.95/ DALY saved | 14 | NR | Unclear (costs & benefits were discounted) | NR |

| Study ID* (Author Year) | INTERVENTION-Micronutrient(s) | INTERVENTION-Food Vehicle(s) | Dose | Other Specs | Compar-ator | Country | Curr-ency | Price Year | HEALTH Outcome Type | Health Result - Difference | Costs - Difference *as REPORTED* | Incremental CE Ratio *as REPORTED* | CONVERTED ICER (using 2022 US$)† | Cost-Benefit Analysis | Dis-count Rate | Uncertainty & Sensitivity Analyses |
|---|---|---|---|---|---|---|---|---|---|---|---|---|---|---|---|---|
| Fiedler 2015 | Vitamins A + B1 + B2 + B3 + B6 + B12 + Folic acid + Calcium + Iron + Zinc | Wheat flour + Vegetable oil | Fe: 55 mg/kg + Zn: 27 mg/kg + Vit. A: flour (4 IU/g) & oil (15 mg/kg); + Ca: 53 mg/kg + Vit. B1: 6 mg/kg + Vit. B2: 4 mg/kg + Vit. B3: 15 mg/kg + Vit. B6: 5 mg/kg + Folic acid: 2 mg/kg + Vit. B12: 0.010 mg/kg | Iron as Ferrous fumarate; + Zinc oxide + Vit. A: flour (250,000 IU/g, dry) & oil (retinyl palmitate 1.7 mIU/g); + Di-Calcium phosphate 2H2O + Thiamine mononitrate + Riboflavin + Niacinamide + Pyridoxine hydrochloride + Folic acid + Vit. B12 0.1% water soluble | No fortification | Bangladesh | US$ | 2012 (unclear) | DALYs saved | 471,599 | 3,173,484 annually | ICER $ 6.73/ DALY saved | 12 | NR | Unclear (costs & benefits were discounted) | NR |
| Fiedler 2013 | Vitamins A + B1 + B2 + B3 + B6 + B12 + Folic acid + | Sugar + Maize meal + Wheat flour | Sugar: Vitamin A (palmitate | Maize as breakfa | Unfortified food | Zambia | US$ | 2006 | DALYs saved | 15,795 | 4,067,202 per year | ICER $ 257/ DALY saved | 263 | NR | NR | Changes in 'prevalence' of inadequate intakes |

| Study ID* (Author Year) | INTERVENTION-Micronutrient(s) | INTERVENTION-Food Vehicle(s) | Dose | Other Specs | Compar-ator | Country | Curr-ency | Price Year | HEALTH Outcome Type | Health Result - Difference | Costs - Difference *as REPORTED* | Incremental CE Ratio *as REPORTED* | CONVERTED ICER (using 2022 US$)† | Cost-Benefit Analysis | Dis-count Rate | Uncertainty & Sensitivity Analyses |
|---|---|---|---|---|---|---|---|---|---|---|---|---|---|---|---|---|
| | Calcium + Iron + Zinc | | 250 CWS/CWD : minimum 10 mg /kg); Maize meal: Vitamin A (retinol 3333IU/kg) + Vitamin B1 (2.0 mg/kg) + Vitamin B2 (2.5 mg/kg) + Vitamin B3 (20.0 mg/kg) + Vitamin B6 (2.5 mg/kg) + Folic acid (1.0 mg/kg) + Vitamin B12 (0.005 mg/kg) + Iron (10.0 mg/kg) + Zinc (15.0 mg/kg); Wheat Flour: Vitamin B1 (4.5-5.5 mg/kg) + Vitamin B2 (2.7-3.5 mg/kg) + Vitamin B3 (35.5-44.4 mg/kg) + Iron (28.9-36.7 mg/kg) + Calcium (1,111–1,444 mg/kg) | st meal and roller meal | | | | | | | | | | | | used to calculate DALYs lost were extracted here. In all instances, "efficiency-based" estimates were more cost-effective than prevalence-based ICERs. |

| Study ID* (Author Year) | INTERVENTION- Micronutrient(s) | INTERVENTION- Food Vehicle(s) | Dose | Other Specs | Compar-ator | Country | Curr-ency | Price Year | HEALTH Outcome Type | Health Result - Difference | Costs - Difference *as REPORTED* | Incremental CE Ratio *as REPORTED* | CONVERTED ICER (using 2022 US$)† | Cost-Benefit Analysis | Dis-count Rate | Uncertainty & Sensitivity Analyses |
|---|---|---|---|---|---|---|---|---|---|---|---|---|---|---|---|---|
| Fiedler 2013 | Vitamins A + B1 + B2 + B3 + B6 + B12 + Folic acid + Calcium + Iron + Zinc | Sugar + Vegetable oil + Maize meal + Wheat flour | Sugar: Vitamin A (palmitate 250 CWS/CWD : minimum 10 mg /kg); Vegetable oil: Vitamin A (retinol 30 mg/kg); Maize meal: Vitamin A (retinol 3333IU/kg) + Vitamin B1 (2.0 mg/kg) + Vitamin B2 (2.5 mg/kg) + Vitamin B3 (20.0 mg/kg) + Vitamin B6 (2.5 mg/kg) + Folic acid (1.0 mg/kg) + Vitamin B12 (0.005 mg/kg) + Iron (10.0 mg/kg) + Zinc (15.0 mg/kg); Wheat Flour: Vitamin B1 (4.5-5.5 mg/kg) + Vitamin B2 (2.7-3.5 mg/kg) + Vitamin B3 (35.5-44.4 mg/kg) + | Maize as breakfast meal and roller meal | Unfortified food | Zambia | US$ | 2006 | DALYs saved | 33,156 | 4,309,602 per year | ICER $ 130/ DALY saved | 133 | NR | NR | Changes in 'prevalence' of inadequate intakes used to calculate DALYs lost were extracted here. In all instances, "efficiency-based" estimates were more cost-effective than prevalence-based ICERs. |

| Study ID* (Author Year) | INTERVENTION-Micronutrient(s) | INTERVENTION-Food Vehicle(s) | Dose | Other Specs | Compar-ator | Country | Curr-ency | Price Year | HEALTH Outcome Type | Health Result - Difference | Costs - Difference *as REPORTED* | Incremental CE Ratio *as REPORTED* | CONVERTED ICER (using 2022 US$)† | Cost-Benefit Analysis | Dis-count Rate | Uncertainty & Sensitivity Analyses |
|---|---|---|---|---|---|---|---|---|---|---|---|---|---|---|---|---|
| | | | Iron (28.9-36.7 mg/kg) + Calcium (1,111–1,444 mg/kg) | | | | | | | | | | | | | |
| Fiedler 2009 | Vitamins A + B1 + B2 + B3 + B6 + B12 + Folic acid + Iron + Zinc | Wheat flour | Vitamin A (Retinyl palmitate 250,000 IU/g) 1.5-4.0 mg RE/kg; Vitamin B1 2-6 mg/kg ; Vitamin B2 3-5 mg/kg; Vitamin B3 5-60 mg/kg; Vitamin B6 2-6 mg/kg; Folic acid 1.5-4 mg/kg; Vitamin B12 0.005-0.020; Iron NaFeEDTA 20 mg/kg or Ferrous fumarate 45 mg/kg; Zinc oxide 10-30 mg/kg | Industrial | Pre-fortification | Afghanistan | US$ | 2009 | DALY saved | NR | 2.0 million | 4.17 | 4 | NR | 3% for DALYs; 0% for costs | NR |
| Fiedler 2009 | Vitamins A + B1 + B2 + B3 + B6 + B12 + Folic acid + Iron + Zinc | Maize flour | Vitamin A (Retinyl palmitate 250,000 IU/g) 1.5-4.0 mg RE/kg; Vitamin B1 2-6 mg/kg ; Vitamin B2 | Industrial | Pre-fortification | Angola | US$ | 2009 | DALY saved | NR | 66.7 million | 985 | 1,329 | NR | 3% for DALYs; 0% for costs | NR |

| Study ID* (Author Year) | INTERVENTION- Micronutrient(s) | INTERVENTION- Food Vehicle(s) | Dose | Other Specs | Compar- ator | Country | Curr- ency | Price Year | HEALTH Outcome Type | Health Result - Difference | Costs - Difference *as REPORTED* | Incremental CE Ratio *as REPORTED* | CONVERTED ICER (using 2022 US$)† | Cost- Benefit Analysis | Dis- count Rate | Uncertainty & Sensitivity Analyses |
|---|---|---|---|---|---|---|---|---|---|---|---|---|---|---|---|---|
| | | | 3-5 mg/kg; Vitamin B3 5-60 mg/kg; Vitamin B6 2-6 mg/kg; Folic acid 1.5-4 mg/kg; Vitamin B12 0.005-0.020; Iron NaFeEDTA 20 mg/kg or Ferrous fumarate 45 mg/kg; Zinc oxide 10-30 mg/kg | | | | | | | | | | | | | |
| Fiedler 2009 | Vitamins A + B1 + B2 + B3 + B6 + B12 + Folic acid + Iron + Zinc | Wheat flour | Vitamin A (Retinyl palmitate 250,000 IU/g) 1.5-4.0 mg RE/kg; Vitamin B1 2-6 mg/kg ; Vitamin B2 3-5 mg/kg; Vitamin B3 5-60 mg/kg; Vitamin B6 2-6 mg/kg; Folic acid 1.5-4 mg/kg; Vitamin B12 0.005-0.020; Iron NaFeEDTA 20 mg/kg or Ferrous fumarate 45 mg/kg; | Industrial | Pre-fortification | Angola | US$ | 2009 | DALY saved | NR | 2.0 million | 5.14 | 7 | NR | 3% for DALYs; 0% for costs | NR |

| Study ID* (Author Year) | INTERVENTION-Micronutrient(s) | INTERVENTION-Food Vehicle(s) | Dose | Other Specs | Compar-ator | Country | Curr-ency | Price Year | HEALTH Outcome Type | Health Result - Difference | Costs - Difference *as REPORTED* | Incremental CE Ratio *as REPORTED* | CONVERTED ICER (using 2022 US$)† | Cost-Benefit Analysis | Dis-count Rate | Uncertainty & Sensitivity Analyses |
|---|---|---|---|---|---|---|---|---|---|---|---|---|---|---|---|---|
| | | | Zinc oxide 10-30 mg/kg | | | | | | | | | | | | | |
| Fiedler 2009 | Vitamins A + B1 + B2 + B3 + B6 + B12 + Folic acid + Iron + Zinc | Wheat flour | Vitamin A (Retinyl palmitate 250,000 IU/g) 1.5-4.0 mg RE/kg; Vitamin B1 2-6 mg/kg ; Vitamin B2 3-5 mg/kg; Vitamin B3 5-60 mg/kg; Vitamin B6 2-6 mg/kg; Folic acid 1.5-4 mg/kg; Vitamin B12 0.005-0.020; Iron NaFeEDTA 20 mg/kg or Ferrous fumarate 45 mg/kg; Zinc oxide 10-30 mg/kg | Industrial | Pre-fortification | Bangladesh | US$ | 2009 | DALY saved | NR | 16.0 million | 19.01 | 36 | NR | 3% for DALYs; 0% for costs | NR |
| Fiedler 2009 | Vitamins A + B1 + B2 + B3 + B6 + B12 + Folic acid + Iron + Zinc | Maize flour | Vitamin A (Retinyl palmitate 250,000 IU/g) 1.5-4.0 mg RE/kg; Vitamin B1 2-6 mg/kg ; Vitamin B2 3-5 mg/kg; Vitamin B3 5-60 mg/kg; | Industrial | Pre-fortification | Bolivia | US$ | 2009 | DALY saved | NR | 153.0 million | 4466 | 7,177 | NR | 3% for DALYs; 0% for costs | NR |

| Study ID* (Author Year) | INTERVENTION-Micronutrient(s) | INTERVENTION-Food Vehicle(s) | Dose | Other Specs | Compar-ator | Country | Curr-ency | Price Year | HEALTH Outcome Type | Health Result - Difference | Costs - Difference *as REPORTED* | Incremental CE Ratio *as REPORTED* | CONVERTED ICER (using 2022 US$)† | Cost-Benefit Analysis | Dis-count Rate | Uncertainty & Sensitivity Analyses |
|---|---|---|---|---|---|---|---|---|---|---|---|---|---|---|---|---|
| | | | Vitamin B6 2-6 mg/kg; Folic acid 1.5-4 mg/kg; Vitamin B12 0.005-0.020; Iron NaFeEDTA 20 mg/kg or Ferrous fumarate 45 mg/kg; Zinc oxide 10-30 mg/kg | | | | | | | | | | | | | |
| Fiedler 2009 | Vitamins A + B1 + B2 + B3 + B6 + B12 + Folic acid + Iron + Zinc | Wheat flour | Vitamin A (Retinyl palmitate 250,000 IU/g) 1.5-4.0 mg RE/kg; Vitamin B1 2-6 mg/kg ; Vitamin B2 3-5 mg/kg; Vitamin B3 5-60 mg/kg; Vitamin B6 2-6 mg/kg; Folic acid 1.5-4 mg/kg; Vitamin B12 0.005-0.020; Iron NaFeEDTA 20 mg/kg or Ferrous fumarate 45 mg/kg; Zinc oxide 10-30 mg/kg | Industrial | Pre-fortification | Brazil | US$ | 2009 | DALY saved | NR | 22.8 million | 28.10 | 27 | NR | 3% for DALYs; 0% for costs | NR |

| Study ID* (Author Year) | INTERVENTION-Micronutrient(s) | INTERVENTION-Food Vehicle(s) | Dose | Other Specs | Compar-ator | Country | Curr-ency | Price Year | HEALTH Outcome Type | Health Result - Difference | Costs - Difference *as REPORTED* | Incremental CE Ratio *as REPORTED* | CONVERTED ICER (using 2022 US$)† | Cost-Benefit Analysis | Dis-count Rate | Uncertainty & Sensitivity Analyses |
|---|---|---|---|---|---|---|---|---|---|---|---|---|---|---|---|---|
| Fiedler 2009 | Vitamins A + B1 + B2 + B3 + B6 + B12 + Folic acid + Iron + Zinc | Wheat flour | Vitamin A (Retinyl palmitate 250,000 IU/g) 1.5-4.0 mg RE/kg; Vitamin B1 2-6 mg/kg ; Vitamin B2 3-5 mg/kg; Vitamin B3 5-60 mg/kg; Vitamin B6 2-6 mg/kg; Folic acid 1.5-4 mg/kg; Vitamin B12 0.005-0.020; Iron NaFeEDTA 20 mg/kg or Ferrous fumarate 45 mg/kg; Zinc oxide 10-30 mg/kg | Industrial | Pre-fortification | Burkina Faso | US$ | 2009 | DALY saved | NR | 1.9 million | 6.06 | 6 | NR | 3% for DALYs; 0% for costs | NR |
| Fiedler 2009 | Vitamins A + B1 + B2 + B3 + B6 + B12 + Folic acid + Iron + Zinc | Wheat flour | Vitamin A (Retinyl palmitate 250,000 IU/g) 1.5-4.0 mg RE/kg; Vitamin B1 2-6 mg/kg ; Vitamin B2 3-5 mg/kg; Vitamin B3 5-60 mg/kg; Vitamin B6 2-6 mg/kg; Folic acid | Industrial | Pre-fortification | Cambodia | US$ | 2009 | DALY saved | NR | 1.7 million | 19.57 | 27 | NR | 3% for DALYs; 0% for costs | NR |

| Study ID* (Author Year) | INTERVENTION-Micronutrient(s) | INTERVENTION-Food Vehicle(s) | Dose | Other Specs | Compar-ator | Country | Curr-ency | Price Year | HEALTH Outcome Type | Health Result - Difference | Costs - Difference *as REPORTED* | Incremental CE Ratio *as REPORTED* | CONVERTED ICER (using 2022 US$)† | Cost-Benefit Analysis | Dis-count Rate | Uncertainty & Sensitivity Analyses |
|---|---|---|---|---|---|---|---|---|---|---|---|---|---|---|---|---|
| | | | 1.5-4 mg/kg; Vitamin B12 0.005-0.020; Iron NaFeEDTA 20 mg/kg or Ferrous fumarate 45 mg/kg; Zinc oxide 10-30 mg/kg | | | | | | | | | | | | | |
| Fiedler 2009 | Vitamins A + B1 + B2 + B3 + B6 + B12 + Folic acid + Iron + Zinc | Wheat flour | Vitamin A (Retinyl palmitate 250,000 IU/g) 1.5-4.0 mg RE/kg; Vitamin B1 2-6 mg/kg ; Vitamin B2 3-5 mg/kg; Vitamin B3 5-60 mg/kg; Vitamin B6 2-6 mg/kg; Folic acid 1.5-4 mg/kg; Vitamin B12 0.005-0.020; Iron NaFeEDTA 20 mg/kg or Ferrous fumarate 45 mg/kg; Zinc oxide 10-30 mg/kg | Industrial | Pre-fortification | Cameroon | US$ | 2009 | DALY saved | NR | 1.6 million | 6.88 | 7 | NR | 3% for DALYs; 0% for costs | NR |
| Fiedler 2009 | Vitamins A + B1 + B2 + B3 + B6 + B12 + Folic acid + Iron + Zinc | Wheat flour | Vitamin A (Retinyl palmitate 250,000 | Industrial | Pre-fortification | China | US$ | 2009 | DALY saved | NR | 907.7 million | 412.99 | 608 | NR | 3% for DALYs; 0% | NR |

| Study ID* (Author Year) | INTERVENTION-Micronutrient(s) | INTERVENTION-Food Vehicle(s) | Dose | Other Specs | Compar-ator | Country | Curr-ency | Price Year | HEALTH Outcome Type | Health Result - Difference | Costs - Difference *as REPORTED* | Incremental CE Ratio *as REPORTED* | CONVERTED ICER (using 2022 US$)† | Cost-Benefit Analysis | Dis-count Rate | Uncertainty & Sensitivity Analyses |
|---|---|---|---|---|---|---|---|---|---|---|---|---|---|---|---|---|
| | | | IU/g) 1.5-4.0 mg RE/kg; Vitamin B1 2-6 mg/kg ; Vitamin B2 3-5 mg/kg; Vitamin B3 5-60 mg/kg; Vitamin B6 2-6 mg/kg; Folic acid 1.5-4 mg/kg; Vitamin B12 0.005-0.020; Iron NaFeEDTA 20 mg/kg or Ferrous fumarate 45 mg/kg; Zinc oxide 10-30 mg/kg | | | | | | | | | | | | for costs | |
| Fiedler 2009 | Vitamins A + B1 + B2 + B3 + B6 + B12 + Folic acid + Iron + Zinc | Wheat flour | Vitamin A (Retinyl palmitate 250,000 IU/g) 1.5-4.0 mg RE/kg; Vitamin B1 2-6 mg/kg ; Vitamin B2 3-5 mg/kg; Vitamin B3 5-60 mg/kg; Vitamin B6 2-6 mg/kg; Folic acid 1.5-4 mg/kg; Vitamin B12 0.005- | Industrial | Pre-fortification | Congo, Dem. Rep. | US$ | 2009 | DALY saved | NR | 1.8 million | 0.94 | 2 | NR | 3% for DALYs; 0% for costs | NR |

| Study ID* (Author Year) | INTERVENTION-Micronutrient(s) | INTERVENTION-Food Vehicle(s) | Dose | Other Specs | Compar-ator | Country | Curr-ency | Price Year | HEALTH Outcome Type | Health Result - Difference | Costs - Difference *as REPORTED* | Incremental CE Ratio *as REPORTED* | CONVERTED ICER (using 2022 US$)† | Cost-Benefit Analysis | Dis-count Rate | Uncertainty & Sensitivity Analyses |
|---|---|---|---|---|---|---|---|---|---|---|---|---|---|---|---|---|
| | | | 0.020; Iron NaFeEDTA 20 mg/kg or Ferrous fumarate 45 mg/kg; Zinc oxide 10-30 mg/kg | | | | | | | | | | | | | |
| Fiedler 2009 | Vitamins A + B1 + B2 + B3 + B6 + B12 + Folic acid + Iron + Zinc | Maize flour | Vitamin A (Retinyl palmitate 250,000 IU/g) 1.5-4.0 mg RE/kg; Vitamin B1 2-6 mg/kg ; Vitamin B2 3-5 mg/kg; Vitamin B3 5-60 mg/kg; Vitamin B6 2-6 mg/kg; Folic acid 1.5-4 mg/kg; Vitamin B12 0.005-0.020; Iron NaFeEDTA 20 mg/kg or Ferrous fumarate 45 mg/kg; Zinc oxide 10-30 mg/kg | Industrial | Pre-fortification | Côte d'Ivoire | US$ | 2009 | DALY saved | NR | 44.9 million | 232 | 227 | NR | 3% for DALYs; 0% for costs | NR |
| Fiedler 2009 | Vitamins A + B1 + B2 + B3 + B6 + B12 + Folic acid + Iron + Zinc | Wheat flour | Vitamin A (Retinyl palmitate 250,000 IU/g) 1.5-4.0 mg RE/kg; Vitamin B1 | Industrial | Pre-fortification | Côte d'Ivoire | US$ | 2009 | DALY saved | NR | 2.0 million | 3.89 | 4 | NR | 3% for DALYs; 0% for costs | NR |

| Study ID* (Author Year) | INTERVENTION-Micronutrient(s) | INTERVENTION-Food Vehicle(s) | Dose | Other Specs | Compar-ator | Country | Curr-ency | Price Year | HEALTH Outcome Type | Health Result - Difference | Costs - Difference *as REPORTED* | Incremental CE Ratio *as REPORTED* | CONVERTED ICER (using 2022 US$)† | Cost-Benefit Analysis | Dis-count Rate | Uncertainty & Sensitivity Analyses |
|---|---|---|---|---|---|---|---|---|---|---|---|---|---|---|---|---|
| | | | 2-6 mg/kg ; Vitamin B2 3-5 mg/kg; Vitamin B3 5-60 mg/kg; Vitamin B6 2-6 mg/kg; Folic acid 1.5-4 mg/kg; Vitamin B12 0.005-0.020; Iron NaFeEDTA 20 mg/kg or Ferrous fumarate 45 mg/kg; Zinc oxide 10-30 mg/kg | | | | | | | | | | | | | |
| Fiedler 2009 | Vitamins A + B1 + B2 + B3 + B6 + B12 + Folic acid + Iron + Zinc | Wheat flour | Vitamin A (Retinyl palmitate 250,000 IU/g) 1.5-4.0 mg RE/kg; Vitamin B1 2-6 mg/kg ; Vitamin B2 3-5 mg/kg; Vitamin B3 5-60 mg/kg; Vitamin B6 2-6 mg/kg; Folic acid 1.5-4 mg/kg; Vitamin B12 0.005-0.020; Iron NaFeEDTA 20 mg/kg or Ferrous | Industrial | Pre-fortification | Ethiopia | US$ | 2009 | DALY saved | NR | 9.3 million | 12.68 | 18 | NR | 3% for DALYs; 0% for costs | NR |

| Study ID* (Author Year) | INTERVENTION-Micronutrient(s) | INTERVENTION-Food Vehicle(s) | Dose | Other Specs | Compar-ator | Country | Curr-ency | Price Year | HEALTH Outcome Type | Health Result - Difference | Costs - Difference *as REPORTED* | Incremental CE Ratio *as REPORTED* | CONVERTED ICER (using 2022 US$)† | Cost-Benefit Analysis | Dis-count Rate | Uncertainty & Sensitivity Analyses |
|---|---|---|---|---|---|---|---|---|---|---|---|---|---|---|---|---|
| | | | fumarate 45 mg/kg; Zinc oxide 10-30 mg/kg | | | | | | | | | | | | | |
| Fiedler 2009 | Vitamins A + B1 + B2 + B3 + B6 + B12 + Folic acid + Iron + Zinc | Maize flour | Vitamin A (Retinyl palmitate 250,000 IU/g) 1.5-4.0 mg RE/kg; Vitamin B1 2-6 mg/kg ; Vitamin B2 3-5 mg/kg; Vitamin B3 5-60 mg/kg; Vitamin B6 2-6 mg/kg; Folic acid 1.5-4 mg/kg; Vitamin B12 0.005-0.020; Iron NaFeEDTA 20 mg/kg or Ferrous fumarate 45 mg/kg; Zinc oxide 10-30 mg/kg | Industrial | Pre-fortification | Ghana | US$ | 2009 | DALY saved | NR | 69.0 million | 491 | 666 | NR | 3% for DALYs; 0% for costs | NR |
| Fiedler 2009 | Vitamins A + B1 + B2 + B3 + B6 + B12 + Folic acid + Iron + Zinc | Wheat flour | Vitamin A (Retinyl palmitate 250,000 IU/g) 1.5-4.0 mg RE/kg; Vitamin B1 2-6 mg/kg ; Vitamin B2 3-5 mg/kg; Vitamin B3 | Industrial | Pre-fortification | Ghana | US$ | 2009 | DALY saved | NR | 2.6 million | 5.88 | 8 | NR | 3% for DALYs; 0% for costs | NR |

| Study ID* (Author Year) | INTERVENTION-Micronutrient(s) | INTERVENTION-Food Vehicle(s) | Dose | Other Specs | Compar-ator | Country | Curr-ency | Price Year | HEALTH Outcome Type | Health Result - Difference | Costs - Difference *as REPORTED* | Incremental CE Ratio *as REPORTED* | CONVERTED ICER (using 2022 US$)† | Cost-Benefit Analysis | Dis-count Rate | Uncertainty & Sensitivity Analyses |
|---|---|---|---|---|---|---|---|---|---|---|---|---|---|---|---|---|
| | | | 5-60 mg/kg; Vitamin B6 2-6 mg/kg; Folic acid 1.5-4 mg/kg; Vitamin B12 0.005-0.020; Iron NaFeEDTA 20 mg/kg or Ferrous fumarate 45 mg/kg; Zinc oxide 10-30 mg/kg | | | | | | | | | | | | | |
| Fiedler 2009 | Vitamins A + B1 + B2 + B3 + B6 + B12 + Folic acid + Iron + Zinc | Maize flour | Vitamin A (Retinyl palmitate 250,000 IU/g) 1.5-4.0 mg RE/kg; Vitamin B1 2-6 mg/kg ; Vitamin B2 3-5 mg/kg; Vitamin B3 5-60 mg/kg; Vitamin B6 2-6 mg/kg; Folic acid 1.5-4 mg/kg; Vitamin B12 0.005-0.020; Iron NaFeEDTA 20 mg/kg or Ferrous fumarate 45 mg/kg; Zinc oxide | Industrial | Pre-fortification | Guatemala | US$ | 2009 | DALY saved | NR | 776.9 million | 11364 | 18,592 | NR | 3% for DALYs; 0% for costs | NR |

| Study ID* (Author Year) | INTERVENTION-Micronutrient(s) | INTERVENTION-Food Vehicle(s) | Dose | Other Specs | Compar-ator | Country | Curr-ency | Price Year | HEALTH Outcome Type | Health Result - Difference | Costs - Difference *as REPORTED* | Incremental CE Ratio *as REPORTED* | CONVERTED ICER (using 2022 US$)† | Cost-Benefit Analysis | Dis-count Rate | Uncertainty & Sensitivity Analyses |
|---|---|---|---|---|---|---|---|---|---|---|---|---|---|---|---|---|
| | | | 10-30 mg/kg | | | | | | | | | | | | | |
| Fiedler 2009 | Vitamins A + B1 + B2 + B3 + B6 + B12 + Folic acid + Iron + Zinc | Wheat flour | Vitamin A (Retinyl palmitate 250,000 IU/g) 1.5-4.0 mg RE/kg; Vitamin B1 2-6 mg/kg ; Vitamin B2 3-5 mg/kg; Vitamin B3 5-60 mg/kg; Vitamin B6 2-6 mg/kg; Folic acid 1.5-4 mg/kg; Vitamin B12 0.005-0.020; Iron NaFeEDTA 20 mg/kg or Ferrous fumarate 45 mg/kg; Zinc oxide 10-30 mg/kg | Industrial | Pre-fortification | Guatemala | US$ | 2009 | DALY saved | NR | 3.4 million | 28.15 | 46 | NR | 3% for DALYs; 0% for costs | NR |
| Fiedler 2009 | Vitamins A + B1 + B2 + B3 + B6 + B12 + Folic acid + Iron + Zinc | Wheat flour | Vitamin A (Retinyl palmitate 250,000 IU/g) 1.5-4.0 mg RE/kg; Vitamin B1 2-6 mg/kg ; Vitamin B2 3-5 mg/kg; Vitamin B3 5-60 mg/kg; Vitamin B6 | Industrial | Pre-fortification | Guinea | US$ | 2009 | DALY saved | NR | 1.6 million | 6.54 | 7 | NR | 3% for DALYs; 0% for costs | NR |

| Study ID* (Author Year) | INTERVENTION-Micronutrient(s) | INTERVENTION-Food Vehicle(s) | Dose | Other Specs | Compar-ator | Country | Curr-ency | Price Year | HEALTH Outcome Type | Health Result - Difference | Costs - Difference *as REPORTED* | Incremental CE Ratio *as REPORTED* | CONVERTED ICER (using 2022 US$)† | Cost-Benefit Analysis | Dis-count Rate | Uncertainty & Sensitivity Analyses |
|---|---|---|---|---|---|---|---|---|---|---|---|---|---|---|---|---|
| | | | 2-6 mg/kg; Folic acid 1.5-4 mg/kg; Vitamin B12 0.005-0.020; Iron NaFeEDTA 20 mg/kg or Ferrous fumarate 45 mg/kg; Zinc oxide 10-30 mg/kg | | | | | | | | | | | | | |
| Fiedler 2009 | Vitamins A + B1 + B2 + B3 + B6 + B12 + Folic acid + Iron + Zinc | Wheat flour | Vitamin A (Retinyl palmitate 250,000 IU/g) 1.5-4.0 mg RE/kg; Vitamin B1 2-6 mg/kg ; Vitamin B2 3-5 mg/kg; Vitamin B3 5-60 mg/kg; Vitamin B6 2-6 mg/kg; Folic acid 1.5-4 mg/kg; Vitamin B12 0.005-0.020; Iron NaFeEDTA 20 mg/kg or Ferrous fumarate 45 mg/kg; Zinc oxide 10-30 mg/kg | Industrial | Pre-fortification | India | US$ | 2009 | DALY saved | NR | 161.1 million | 15.76 | 20 | NR | 3% for DALYs; 0% for costs | NR |
| Fiedler 2009 | Vitamins A + B1 + B2 + B3 + B6 + B12 | Wheat flour | Vitamin A (Retinyl | Industrial | Pre-fortification | Indonesia | US$ | 2009 | DALY saved | NR | 3.0 million | 3.45 | 5 | NR | 3% for DALY | NR |

| Study ID* (Author Year) | INTERVENTION- Micronutrient(s) | INTERVENTION- Food Vehicle(s) | Dose | Other Specs | Compar- ator | Country | Curr- ency | Price Year | HEALTH Outcome Type | Health Result - Difference | Costs - Difference *as REPORTED* | Incremental CE Ratio *as REPORTED* | CONVERTED ICER (using 2022 US$)† | Cost- Benefit Analysis | Dis- count Rate | Uncertainty & Sensitivity Analyses |
|---|---|---|---|---|---|---|---|---|---|---|---|---|---|---|---|---|
| | + Folic acid + Iron + Zinc | | palmitate 250,000 IU/g) 1.5-4.0 mg RE/kg; Vitamin B1 2-6 mg/kg ; Vitamin B2 3-5 mg/kg; Vitamin B3 5-60 mg/kg; Vitamin B6 2-6 mg/kg; Folic acid 1.5-4 mg/kg; Vitamin B12 0.005-0.020; Iron NaFeEDTA 20 mg/kg or Ferrous fumarate 45 mg/kg; Zinc oxide 10-30 mg/kg | | | | | | | | | | | | s; 0% for costs | |
| Fiedler 2009 | Vitamins A + B1 + B2 + B3 + B6 + B12 + Folic acid + Iron + Zinc | Maize flour | Vitamin A (Retinyl palmitate 250,000 IU/g) 1.5-4.0 mg RE/kg; Vitamin B1 2-6 mg/kg ; Vitamin B2 3-5 mg/kg; Vitamin B3 5-60 mg/kg; Vitamin B6 2-6 mg/kg; Folic acid 1.5-4 mg/kg; | Industrial | Pre-fortification | Kenya | US$ | 2009 | DALY saved | NR | 312.8 million | 1252 | 1,825 | NR | 3% for DALYs; 0% for costs | NR |

| Study ID* (Author Year) | INTERVENTION- Micronutrient(s) | INTERVENTION- Food Vehicle(s) | Dose | Other Specs | Compar-ator | Country | Curr-ency | Price Year | HEALTH Outcome Type | Health Result - Difference | Costs - Difference *as REPORTED* | Incremental CE Ratio *as REPORTED* | CONVERTED ICER (using 2022 US$)† | Cost-Benefit Analysis | Dis-count Rate | Uncertainty & Sensitivity Analyses |
|---|---|---|---|---|---|---|---|---|---|---|---|---|---|---|---|---|
| | | | Vitamin B12 0.005-0.020; Iron NaFeEDTA 20 mg/kg or Ferrous fumarate 45 mg/kg; Zinc oxide 10-30 mg/kg | | | | | | | | | | | | | |
| Fiedler 2009 | Vitamins A + B1 + B2 + B3 + B6 + B12 + Folic acid + Iron + Zinc | Wheat flour | Vitamin A (Retinyl palmitate 250,000 IU/g) 1.5-4.0 mg RE/kg; Vitamin B1 2-6 mg/kg ; Vitamin B2 3-5 mg/kg; Vitamin B3 5-60 mg/kg; Vitamin B6 2-6 mg/kg; Folic acid 1.5-4 mg/kg; Vitamin B12 0.005-0.020; Iron NaFeEDTA 20 mg/kg or Ferrous fumarate 45 mg/kg; Zinc oxide 10-30 mg/kg | Industrial | Pre-fortification | Kenya | US$ | 2009 | DALY saved | NR | 2.4 million | 7.53 | 11 | NR | 3% for DALYs; 0% for costs | NR |
| Fiedler 2009 | Vitamins A + B1 + B2 + B3 + B6 + B12 + Folic acid + Iron + Zinc | Wheat flour | Vitamin A (Retinyl palmitate 250,000 IU/g) 1.5-4.0 mg | Industrial | Pre-fortification | Madagascar | US$ | 2009 | DALY saved | NR | 1.7 million | 7.03 | 8 | NR | 3% for DALYs; 0% for costs | NR |

| Study ID* (Author Year) | INTERVENTION-Micronutrient(s) | INTERVENTION-Food Vehicle(s) | Dose | Other Specs | Compar-ator | Country | Curr-ency | Price Year | HEALTH Outcome Type | Health Result - Difference | Costs - Difference *as REPORTED* | Incremental CE Ratio *as REPORTED* | CONVERTED ICER (using 2022 US$)† | Cost-Benefit Analysis | Dis-count Rate | Uncertainty & Sensitivity Analyses |
|---|---|---|---|---|---|---|---|---|---|---|---|---|---|---|---|---|
| | | | RE/kg; Vitamin B1 2-6 mg/kg ; Vitamin B2 3-5 mg/kg; Vitamin B3 5-60 mg/kg; Vitamin B6 2-6 mg/kg; Folic acid 1.5-4 mg/kg; Vitamin B12 0.005-0.020; Iron NaFeEDTA 20 mg/kg or Ferrous fumarate 45 mg/kg; Zinc oxide 10-30 mg/kg | | | | | | | | | | | | | |
| Fiedler 2009 | Vitamins A + B1 + B2 + B3 + B6 + B12 + Folic acid + Iron + Zinc | Maize flour | Vitamin A (Retinyl palmitate 250,000 IU/g) 1.5-4.0 mg RE/kg; Vitamin B1 2-6 mg/kg ; Vitamin B2 3-5 mg/kg; Vitamin B3 5-60 mg/kg; Vitamin B6 2-6 mg/kg; Folic acid 1.5-4 mg/kg; Vitamin B12 0.005-0.020; Iron NaFeEDTA | Industrial | Pre-fortification | Malawi | US$ | 2009 | DALY saved | NR | 62.1 million | 120 | 104 | NR | 3% for DALYs; 0% for costs | NR |

| Study ID* (Author Year) | INTERVENTION-Micronutrient(s) | INTERVENTION-Food Vehicle(s) | Dose | Other Specs | Compar-ator | Country | Curr-ency | Price Year | HEALTH Outcome Type | Health Result - Difference | Costs - Difference *as REPORTED* | Incremental CE Ratio *as REPORTED* | CONVERTED ICER (using 2022 US$)† | Cost-Benefit Analysis | Dis-count Rate | Uncertainty & Sensitivity Analyses |
|---|---|---|---|---|---|---|---|---|---|---|---|---|---|---|---|---|
| | | | 20 mg/kg or Ferrous fumarate 45 mg/kg; Zinc oxide 10-30 mg/kg | | | | | | | | | | | | | |
| Fiedler 2009 | Vitamins A + B1 + B2 + B3 + B6 + B12 + Folic acid + Iron + Zinc | Wheat flour | Vitamin A (Retinyl palmitate 250,000 IU/g) 1.5-4.0 mg RE/kg; Vitamin B1 2-6 mg/kg ; Vitamin B2 3-5 mg/kg; Vitamin B3 5-60 mg/kg; Vitamin B6 2-6 mg/kg; Folic acid 1.5-4 mg/kg; Vitamin B12 0.005-0.020; Iron NaFeEDTA 20 mg/kg or Ferrous fumarate 45 mg/kg; Zinc oxide 10-30 mg/kg | Industrial | Pre-fortification | Malawi | US$ | 2009 | DALY saved | NR | 1.9 million | 10.81 | 9 | NR | 3% for DALYs; 0% for costs | NR |
| Fiedler 2009 | Vitamins A + B1 + B2 + B3 + B6 + B12 + Folic acid + Iron + Zinc | Wheat flour | Vitamin A (Retinyl palmitate 250,000 IU/g) 1.5-4.0 mg RE/kg; Vitamin B1 2-6 mg/kg ; Vitamin B2 | Industrial | Pre-fortification | Mali | US$ | 2009 | DALY saved | NR | 1.8 million | 4.68 | 5 | NR | 3% for DALYs; 0% for costs | NR |

| Study ID* (Author Year) | INTERVENTION-Micronutrient(s) | INTERVENTION-Food Vehicle(s) | Dose | Other Specs | Compar-ator | Country | Curr-ency | Price Year | HEALTH Outcome Type | Health Result - Difference | Costs - Difference *as REPORTED* | Incremental CE Ratio *as REPORTED* | CONVERTED ICER (using 2022 US$)† | Cost-Benefit Analysis | Dis-count Rate | Uncertainty & Sensitivity Analyses |
|---|---|---|---|---|---|---|---|---|---|---|---|---|---|---|---|---|
| | | | 3-5 mg/kg; Vitamin B3 5-60 mg/kg; Vitamin B6 2-6 mg/kg; Folic acid 1.5-4 mg/kg; Vitamin B12 0.005-0.020; Iron NaFeEDTA 20 mg/kg or Ferrous fumarate 45 mg/kg; Zinc oxide 10-30 mg/kg | | | | | | | | | | | | | |
| Fiedler 2009 | Vitamins A + B1 + B2 + B3 + B6 + B12 + Folic acid + Iron + Zinc | Maize flour | Vitamin A (Retinyl palmitate 250,000 IU/g) 1.5-4.0 mg RE/kg; Vitamin B1 2-6 mg/kg ; Vitamin B2 3-5 mg/kg; Vitamin B3 5-60 mg/kg; Vitamin B6 2-6 mg/kg; Folic acid 1.5-4 mg/kg; Vitamin B12 0.005-0.020; Iron NaFeEDTA 20 mg/kg or Ferrous fumarate 45 mg/kg; | Industrial | Pre-fortification | Mexico | US$ | 2009 | DALY saved | NR | 8273.7 million | 18048 | 22,180 | NR | 3% for DALYs; 0% for costs | NR |

| Study ID* (Author Year) | INTERVENTION-Micronutrient(s) | INTERVENTION-Food Vehicle(s) | Dose | Other Specs | Compar-ator | Country | Curr-ency | Price Year | HEALTH Outcome Type | Health Result - Difference | Costs - Difference *as REPORTED* | Incremental CE Ratio *as REPORTED* | CONVERTED ICER (using 2022 US$)† | Cost-Benefit Analysis | Dis-count Rate | Uncertainty & Sensitivity Analyses |
|---|---|---|---|---|---|---|---|---|---|---|---|---|---|---|---|---|
| | | | Zinc oxide 10-30 mg/kg | | | | | | | | | | | | | |
| Fiedler 2009 | Vitamins A + B1 + B2 + B3 + B6 + B12 + Folic acid + Iron + Zinc | Wheat flour | Vitamin A (Retinyl palmitate 250,000 IU/g) 1.5-4.0 mg RE/kg; Vitamin B1 2-6 mg/kg ; Vitamin B2 3-5 mg/kg; Vitamin B3 5-60 mg/kg; Vitamin B6 2-6 mg/kg; Folic acid 1.5-4 mg/kg; Vitamin B12 0.005-0.020; Iron NaFeEDTA 20 mg/kg or Ferrous fumarate 45 mg/kg; Zinc oxide 10-30 mg/kg | Industrial | Pre-fortification | Mexico | US$ | 2009 | DALY saved | NR | 25.7 million | 56.06 | 69 | NR | 3% for DALYs; 0% for costs | NR |
| Fiedler 2009 | Vitamins A + B1 + B2 + B3 + B6 + B12 + Folic acid + Iron + Zinc | Wheat flour | Vitamin A (Retinyl palmitate 250,000 IU/g) 1.5-4.0 mg RE/kg; Vitamin B1 2-6 mg/kg ; Vitamin B2 3-5 mg/kg; Vitamin B3 5-60 mg/kg; | Industrial | Pre-fortification | Mozambique | US$ | 2009 | DALY saved | NR | 4.6 million | 22.38 | 18 | NR | 3% for DALYs; 0% for costs | NR |

| Study ID* (Author Year) | INTERVENTION-Micronutrient(s) | INTERVENTION-Food Vehicle(s) | Dose | Other Specs | Compar-ator | Country | Curr-ency | Price Year | HEALTH Outcome Type | Health Result - Difference | Costs - Difference *as REPORTED* | Incremental CE Ratio *as REPORTED* | CONVERTED ICER (using 2022 US$)† | Cost-Benefit Analysis | Dis-count Rate | Uncertainty & Sensitivity Analyses |
|---|---|---|---|---|---|---|---|---|---|---|---|---|---|---|---|---|
| | | | Vitamin B6 2-6 mg/kg; Folic acid 1.5-4 mg/kg; Vitamin B12 0.005-0.020; Iron NaFeEDTA 20 mg/kg or Ferrous fumarate 45 mg/kg; Zinc oxide 10-30 mg/kg | | | | | | | | | | | | | |
| Fiedler 2009 | Vitamins A + B1 + B2 + B3 + B6 + B12 + Folic acid + Iron + Zinc | Wheat flour | Vitamin A (Retinyl palmitate 250,000 IU/g) 1.5-4.0 mg RE/kg; Vitamin B1 2-6 mg/kg ; Vitamin B2 3-5 mg/kg; Vitamin B3 5-60 mg/kg; Vitamin B6 2-6 mg/kg; Folic acid 1.5-4 mg/kg; Vitamin B12 0.005-0.020; Iron NaFeEDTA 20 mg/kg or Ferrous fumarate 45 mg/kg; Zinc oxide 10-30 mg/kg | Industrial | Pre-fortification | Myanmar | US$ | 2009 | DALY saved | NR | 1.7 million | 3.03 | 1 | NR | 3% for DALYs; 0% for costs | NR |

| Study ID* (Author Year) | INTERVENTION-Micronutrient(s) | INTERVENTION-Food Vehicle(s) | Dose | Other Specs | Compar-ator | Country | Curr-ency | Price Year | HEALTH Outcome Type | Health Result - Difference | Costs - Difference *as REPORTED* | Incremental CE Ratio *as REPORTED* | CONVERTED ICER (using 2022 US$)[†] | Cost-Benefit Analysis | Dis-count Rate | Uncertainty & Sensitivity Analyses |
|---|---|---|---|---|---|---|---|---|---|---|---|---|---|---|---|---|
| Fiedler 2009 | Vitamins A + B1 + B2 + B3 + B6 + B12 + Folic acid + Iron + Zinc | Wheat flour | Vitamin A (Retinyl palmitate 250,000 IU/g) 1.5-4.0 mg RE/kg; Vitamin B1 2-6 mg/kg ; Vitamin B2 3-5 mg/kg; Vitamin B3 5-60 mg/kg; Vitamin B6 2-6 mg/kg; Folic acid 1.5-4 mg/kg; Vitamin B12 0.005-0.020; Iron NaFeEDTA 20 mg/kg or Ferrous fumarate 45 mg/kg; Zinc oxide 10-30 mg/kg | Industrial | Pre-fortification | Nepal | US$ | 2009 | DALY saved | NR | 3.3 million | 8.73 | 15 | NR | 3% for DALYs; 0% for costs | NR |
| Fiedler 2009 | Vitamins A + B1 + B2 + B3 + B6 + B12 + Folic acid + Iron + Zinc | Wheat flour | Vitamin A (Retinyl palmitate 250,000 IU/g) 1.5-4.0 mg RE/kg; Vitamin B1 2-6 mg/kg ; Vitamin B2 3-5 mg/kg; Vitamin B3 5-60 mg/kg; Vitamin B6 2-6 mg/kg; Folic acid | Industrial | Pre-fortification | Niger | US$ | 2009 | DALY saved | NR | 1.8 million | 7.36 | 7 | NR | 3% for DALYs; 0% for costs | NR |

| Study ID* (Author Year) | INTERVENTION-Micronutrient(s) | INTERVENTION-Food Vehicle(s) | Dose | Other Specs | Compar-ator | Country | Curr-ency | Price Year | HEALTH Outcome Type | Health Result - Difference | Costs - Difference *as REPORTED* | Incremental CE Ratio *as REPORTED* | CONVERTED ICER (using 2022 US$)† | Cost-Benefit Analysis | Dis-count Rate | Uncertainty & Sensitivity Analyses |
|---|---|---|---|---|---|---|---|---|---|---|---|---|---|---|---|---|
| | | | 1.5-4 mg/kg; Vitamin B12 0.005-0.020; Iron NaFeEDTA 20 mg/kg or Ferrous fumarate 45 mg/kg; Zinc oxide 10-30 mg/kg | | | | | | | | | | | | | |
| Fiedler 2009 | Vitamins A + B1 + B2 + B3 + B6 + B12 + Folic acid + Iron + Zinc | Wheat flour | Vitamin A (Retinyl palmitate 250,000 IU/g) 1.5-4.0 mg RE/kg; Vitamin B1 2-6 mg/kg ; Vitamin B2 3-5 mg/kg; Vitamin B3 5-60 mg/kg; Vitamin B6 2-6 mg/kg; Folic acid 1.5-4 mg/kg; Vitamin B12 0.005-0.020; Iron NaFeEDTA 20 mg/kg or Ferrous fumarate 45 mg/kg; Zinc oxide 10-30 mg/kg | Industrial | Pre-fortification | Nigeria | US$ | 2009 | DALY saved | NR | 3.3 million | 0.85 | 1 | NR | 3% for DALYs; 0% for costs | NR |
| Fiedler 2009 | Vitamins A + B1 + B2 + B3 + B6 + B12 + Folic acid + Iron + Zinc | Wheat flour | Vitamin A (Retinyl palmitate 250,000 | Industrial | Pre-fortification | Pakistan | US$ | 2009 | DALY saved | NR | 68.8 million | 97.87 | 109 | NR | 3% for DALYs; 0% | NR |

| Study ID* (Author Year) | INTERVENTION- Micronutrient(s) | INTERVENTION- Food Vehicle(s) | Dose | Other Specs | Compar- ator | Country | Curr- ency | Price Year | HEALTH Outcome Type | Health Result - Difference | Costs - Difference *as REPORTED* | Incremental CE Ratio *as REPORTED* | CONVERTED ICER (using 2022 US$)† | Cost- Benefit Analysis | Dis- count Rate | Uncertainty & Sensitivity Analyses |
|---|---|---|---|---|---|---|---|---|---|---|---|---|---|---|---|---|
| | | | IU/g) 1.5-4.0 mg RE/kg; Vitamin B1 2-6 mg/kg ; Vitamin B2 3-5 mg/kg; Vitamin B3 5-60 mg/kg; Vitamin B6 2-6 mg/kg; Folic acid 1.5-4 mg/kg; Vitamin B12 0.005-0.020; Iron NaFeEDTA 20 mg/kg or Ferrous fumarate 45 mg/kg; Zinc oxide 10-30 mg/kg | | | | | | | | | | | | for costs | |
| Fiedler 2009 | Vitamins A + B1 + B2 + B3 + B6 + B12 + Folic acid + Iron + Zinc | Wheat flour | Vitamin A (Retinyl palmitate 250,000 IU/g) 1.5-4.0 mg RE/kg; Vitamin B1 2-6 mg/kg ; Vitamin B2 3-5 mg/kg; Vitamin B3 5-60 mg/kg; Vitamin B6 2-6 mg/kg; Folic acid 1.5-4 mg/kg; Vitamin B12 0.005- | Industrial | Pre-fortification | Peru | US$ | 2009 | DALY saved | NR | 2.9 million | 18.31 | 23 | NR | 3% for DALYs; 0% for costs | NR |

| Study ID* (Author Year) | INTERVENTION-Micronutrient(s) | INTERVENTION-Food Vehicle(s) | Dose | Other Specs | Compar-ator | Country | Curr-ency | Price Year | HEALTH Outcome Type | Health Result - Difference | Costs - Difference *as REPORTED* | Incremental CE Ratio *as REPORTED* | CONVERTED ICER (using 2022 US$)† | Cost-Benefit Analysis | Dis-count Rate | Uncertainty & Sensitivity Analyses |
|---|---|---|---|---|---|---|---|---|---|---|---|---|---|---|---|---|
| | | | 0.020; Iron NaFeEDTA 20 mg/kg or Ferrous fumarate 45 mg/kg; Zinc oxide 10-30 mg/kg | | | | | | | | | | | | | |
| Fiedler 2009 | Vitamins A + B1 + B2 + B3 + B6 + B12 + Folic acid + Iron + Zinc | Wheat flour | Vitamin A (Retinyl palmitate 250,000 IU/g) 1.5-4.0 mg RE/kg; Vitamin B1 2-6 mg/kg ; Vitamin B2 3-5 mg/kg; Vitamin B3 5-60 mg/kg; Vitamin B6 2-6 mg/kg; Folic acid 1.5-4 mg/kg; Vitamin B12 0.005-0.020; Iron NaFeEDTA 20 mg/kg or Ferrous fumarate 45 mg/kg; Zinc oxide 10-30 mg/kg | Industrial | Pre-fortification | Philippines | US$ | 2009 | DALY saved | NR | 4.7 million | 16.05 | 19 | NR | 3% for DALYs; 0% for costs | NR |
| Fiedler 2009 | Vitamins A + B1 + B2 + B3 + B6 + B12 + Folic acid + Iron + Zinc | Maize flour | Vitamin A (Retinyl palmitate 250,000 IU/g) 1.5-4.0 mg RE/kg; Vitamin B1 | Industrial | Pre-fortification | South Africa | US$ | 2009 | DALY saved | NR | 1913.2 million | 8243 | 8,495 | NR | 3% for DALYs; 0% for costs | NR |

| Study ID* (Author Year) | INTERVENTION-Micronutrient(s) | INTERVENTION-Food Vehicle(s) | Dose | Other Specs | Compar-ator | Country | Curr-ency | Price Year | HEALTH Outcome Type | Health Result - Difference | Costs - Difference *as REPORTED* | Incremental CE Ratio *as REPORTED* | CONVERTED ICER (using 2022 US$)† | Cost-Benefit Analysis | Dis-count Rate | Uncertainty & Sensitivity Analyses |
|---|---|---|---|---|---|---|---|---|---|---|---|---|---|---|---|---|
| | | | 2-6 mg/kg ; Vitamin B2 3-5 mg/kg; Vitamin B3 5-60 mg/kg; Vitamin B6 2-6 mg/kg; Folic acid 1.5-4 mg/kg; Vitamin B12 0.005-0.020; Iron NaFeEDTA 20 mg/kg or Ferrous fumarate 45 mg/kg; Zinc oxide 10-30 mg/kg | | | | | | | | | | | | | |
| Fiedler 2009 | Vitamins A + B1 + B2 + B3 + B6 + B12 + Folic acid + Iron + Zinc | Wheat flour | Vitamin A (Retinyl palmitate 250,000 IU/g) 1.5-4.0 mg RE/kg; Vitamin B1 2-6 mg/kg ; Vitamin B2 3-5 mg/kg; Vitamin B3 5-60 mg/kg; Vitamin B6 2-6 mg/kg; Folic acid 1.5-4 mg/kg; Vitamin B12 0.005-0.020; Iron NaFeEDTA 20 mg/kg or Ferrous | Industrial | Pre-fortification | South Africa | US$ | 2009 | DALY saved | NR | 3.3 million | 21.07 | 22 | NR | 3% for DALYs; 0% for costs | NR |

| Study ID* (Author Year) | INTERVENTION-Micronutrient(s) | INTERVENTION-Food Vehicle(s) | Dose | Other Specs | Compar-ator | Country | Curr-ency | Price Year | HEALTH Outcome Type | Health Result - Difference | Costs - Difference *as REPORTED* | Incremental CE Ratio *as REPORTED* | CONVERTED ICER (using 2022 US$)† | Cost-Benefit Analysis | Dis-count Rate | Uncertainty & Sensitivity Analyses |
|---|---|---|---|---|---|---|---|---|---|---|---|---|---|---|---|---|
| | | | fumarate 45 mg/kg; Zinc oxide 10-30 mg/kg | | | | | | | | | | | | | |
| Fiedler 2009 | Vitamins A + B1 + B2 + B3 + B6 + B12 + Folic acid + Iron + Zinc | Wheat flour | Vitamin A (Retinyl palmitate 250,000 IU/g) 1.5-4.0 mg RE/kg; Vitamin B1 2-6 mg/kg ; Vitamin B2 3-5 mg/kg; Vitamin B3 5-60 mg/kg; Vitamin B6 2-6 mg/kg; Folic acid 1.5-4 mg/kg; Vitamin B12 0.005-0.020; Iron NaFeEDTA 20 mg/kg or Ferrous fumarate 45 mg/kg; Zinc oxide 10-30 mg/kg | Industrial | Pre-fortification | Sudan | US$ | 2009 | DALY saved | NR | 5.0 million | 18.94 | 20 | NR | 3% for DALYs; 0% for costs | NR |
| Fiedler 2009 | Vitamins A + B1 + B2 + B3 + B6 + B12 + Folic acid + Iron + Zinc | Maize flour | Vitamin A (Retinyl palmitate 250,000 IU/g) 1.5-4.0 mg RE/kg; Vitamin B1 2-6 mg/kg ; Vitamin B2 3-5 mg/kg; Vitamin B3 | Industrial | Pre-fortification | Tanzania | US$ | 2009 | DALY saved | NR | 156.3 million | 201 | 256 | NR | 3% for DALYs; 0% for costs | NR |

| Study ID* (Author Year) | INTERVENTION-Micronutrient(s) | INTERVENTION-Food Vehicle(s) | Dose | Other Specs | Compar-ator | Country | Curr-ency | Price Year | HEALTH Outcome Type | Health Result - Difference | Costs - Difference *as REPORTED* | Incremental CE Ratio *as REPORTED* | CONVERTED ICER (using 2022 US$)† | Cost-Benefit Analysis | Dis-count Rate | Uncertainty & Sensitivity Analyses |
|---|---|---|---|---|---|---|---|---|---|---|---|---|---|---|---|---|
| | | | 5-60 mg/kg; Vitamin B6 2-6 mg/kg; Folic acid 1.5-4 mg/kg; Vitamin B12 0.005-0.020; Iron NaFeEDTA 20 mg/kg or Ferrous fumarate 45 mg/kg; Zinc oxide 10-30 mg/kg | | | | | | | | | | | | | |
| Fiedler 2009 | Vitamins A + B1 + B2 + B3 + B6 + B12 + Folic acid + Iron + Zinc | Wheat flour | Vitamin A (Retinyl palmitate 250,000 IU/g) 1.5-4.0 mg RE/kg; Vitamin B1 2-6 mg/kg ; Vitamin B2 3-5 mg/kg; Vitamin B3 5-60 mg/kg; Vitamin B6 2-6 mg/kg; Folic acid 1.5-4 mg/kg; Vitamin B12 0.005-0.020; Iron NaFeEDTA 20 mg/kg or Ferrous fumarate 45 mg/kg; Zinc oxide | Industrial | Pre-fortification | Tanzania | US$ | 2009 | DALY saved | NR | 2.2 million | 5.61 | 7 | NR | 3% for DALYs; 0% for costs | NR |

| Study ID* (Author Year) | INTERVENTION-Micronutrient(s) | INTERVENTION-Food Vehicle(s) | Dose | Other Specs | Compar-ator | Country | Curr-ency | Price Year | HEALTH Outcome Type | Health Result - Difference | Costs - Difference *as REPORTED* | Incremental CE Ratio *as REPORTED* | CONVERTED ICER (using 2022 US$)† | Cost-Benefit Analysis | Dis-count Rate | Uncertainty & Sensitivity Analyses |
|---|---|---|---|---|---|---|---|---|---|---|---|---|---|---|---|---|
| | | | 10-30 mg/kg | | | | | | | | | | | | | |
| Fiedler 2009 | Vitamins A + B1 + B2 + B3 + B6 + B12 + Folic acid + Iron + Zinc | Wheat flour | Vitamin A (Retinyl palmitate 250,000 IU/g) 1.5-4.0 mg RE/kg; Vitamin B1 2-6 mg/kg ; Vitamin B2 3-5 mg/kg; Vitamin B3 5-60 mg/kg; Vitamin B6 2-6 mg/kg; Folic acid 1.5-4 mg/kg; Vitamin B12 0.005-0.020; Iron NaFeEDTA 20 mg/kg or Ferrous fumarate 45 mg/kg; Zinc oxide 10-30 mg/kg | Industrial | Pre-fortification | Turkey | US$ | 2009 | DALY saved | NR | 71.2 million | 168.28 | 111 | NR | 3% for DALYs; 0% for costs | NR |
| Fiedler 2009 | Vitamins A + B1 + B2 + B3 + B6 + B12 + Folic acid + Iron + Zinc | Maize flour | Vitamin A (Retinyl palmitate 250,000 IU/g) 1.5-4.0 mg RE/kg; Vitamin B1 2-6 mg/kg ; Vitamin B2 3-5 mg/kg; Vitamin B3 5-60 mg/kg; Vitamin B6 | Industrial | Pre-fortification | Uganda | US$ | 2009 | DALY saved | NR | 57.9 million | 197 | 194 | NR | 3% for DALYs; 0% for costs | NR |

| Study ID* (Author Year) | INTERVENTION-Micronutrient(s) | INTERVENTION-Food Vehicle(s) | Dose | Other Specs | Compar-ator | Country | Curr-ency | Price Year | HEALTH Outcome Type | Health Result - Difference | Costs - Difference *as REPORTED* | Incremental CE Ratio *as REPORTED* | CONVERTED ICER (using 2022 US$)† | Cost-Benefit Analysis | Dis-count Rate | Uncertainty & Sensitivity Analyses |
|---|---|---|---|---|---|---|---|---|---|---|---|---|---|---|---|---|
| | | | 2-6 mg/kg; Folic acid 1.5-4 mg/kg; Vitamin B12 0.005-0.020; Iron NaFeEDTA 20 mg/kg or Ferrous fumarate 45 mg/kg; Zinc oxide 10-30 mg/kg | | | | | | | | | | | | | |
| Fiedler 2009 | Vitamins A + B1 + B2 + B3 + B6 + B12 + Folic acid + Iron + Zinc | Wheat flour | Vitamin A (Retinyl palmitate 250,000 IU/g) 1.5-4.0 mg RE/kg; Vitamin B1 2-6 mg/kg ; Vitamin B2 3-5 mg/kg; Vitamin B3 5-60 mg/kg; Vitamin B6 2-6 mg/kg; Folic acid 1.5-4 mg/kg; Vitamin B12 0.005-0.020; Iron NaFeEDTA 20 mg/kg or Ferrous fumarate 45 mg/kg; Zinc oxide 10-30 mg/kg | Industrial | Pre-fortification | Uzbekistan | US$ | 2009 | DALY saved | NR | 4.8 million | 24.96 | 33 | NR | 3% for DALYs; 0% for costs | NR |
| Fiedler 2009 | Vitamins A + B1 + B2 + B3 + B6 + B12 | Wheat flour | Vitamin A (Retinyl | Industrial | Pre-fortification | Vietnam | US$ | 2009 | DALY saved | NR | 3.5 million | 30.52 | 55 | NR | 3% for DALY | NR |

| Study ID* (Author Year) | INTERVENTION- Micronutrient(s) | INTERVENTION- Food Vehicle(s) | Dose | Other Specs | Compar-ator | Country | Curr-ency | Price Year | HEALTH Outcome Type | Health Result - Difference | Costs - Difference *as REPORTED* | Incremental CE Ratio *as REPORTED* | CONVERTED ICER (using 2022 US$)† | Cost-Benefit Analysis | Dis-count Rate | Uncertainty & Sensitivity Analyses |
|---|---|---|---|---|---|---|---|---|---|---|---|---|---|---|---|---|
| | + Folic acid + Iron + Zinc | | palmitate 250,000 IU/g) 1.5-4.0 mg RE/kg; Vitamin B1 2-6 mg/kg ; Vitamin B2 3-5 mg/kg; Vitamin B3 5-60 mg/kg; Vitamin B6 2-6 mg/kg; Folic acid 1.5-4 mg/kg; Vitamin B12 0.005-0.020; Iron NaFeEDTA 20 mg/kg or Ferrous fumarate 45 mg/kg; Zinc oxide 10-30 mg/kg | | | | | | | | | | | | s; 0% for costs | |
| Fiedler 2009 | Vitamins A + B1 + B2 + B3 + B6 + B12 + Folic acid + Iron + Zinc | Wheat flour | Vitamin A (Retinyl palmitate 250,000 IU/g) 1.5-4.0 mg RE/kg; Vitamin B1 2-6 mg/kg ; Vitamin B2 3-5 mg/kg; Vitamin B3 5-60 mg/kg; Vitamin B6 2-6 mg/kg; Folic acid 1.5-4 mg/kg; | Industrial | Pre-fortification | Yemen | US$ | 2009 | DALY saved | NR | 3.2 million | 16.80 | 27 | NR | 3% for DALYs; 0% for costs | NR |

| Study ID* (Author Year) | INTERVENTION-Micronutrient(s) | INTERVENTION-Food Vehicle(s) | Dose | Other Specs | Compar-ator | Country | Curr-ency | Price Year | HEALTH Outcome Type | Health Result - Difference | Costs - Difference *as REPORTED* | Incremental CE Ratio *as REPORTED* | CONVERTED ICER (using 2022 US$)† | Cost-Benefit Analysis | Dis-count Rate | Uncertainty & Sensitivity Analyses |
|---|---|---|---|---|---|---|---|---|---|---|---|---|---|---|---|---|
| | | | Vitamin B12 0.005-0.020; Iron NaFeEDTA 20 mg/kg or Ferrous fumarate 45 mg/kg; Zinc oxide 10-30 mg/kg | | | | | | | | | | | | | |
| Fiedler 2009 | Vitamins A + B1 + B2 + B3 + B6 + B12 + Folic acid + Iron + Zinc | Maize flour | Vitamin A (Retinyl palmitate 250,000 IU/g) 1.5-4.0 mg RE/kg; Vitamin B1 2-6 mg/kg ; Vitamin B2 3-5 mg/kg; Vitamin B3 5-60 mg/kg; Vitamin B6 2-6 mg/kg; Folic acid 1.5-4 mg/kg; Vitamin B12 0.005-0.020; Iron NaFeEDTA 20 mg/kg or Ferrous fumarate 45 mg/kg; Zinc oxide 10-30 mg/kg | Industrial | Pre-fortification | Zambia | US$ | 2009 | DALY saved | NR | 73.5 million | 659 | 717 | NR | 3% for DALYs; 0% for costs | NR |
| Fiedler 2013 | Vitamins A + B1 + B2 + B3 + B6 + B12 + Folic acid + Iron + Zinc | Maize meal | Vitamin A (retinol 3333IU/kg) + Vitamin B1 (2.0 mg/kg) + | Maize as breakfast meal and | Unfortified food | Zambia | US$ | 2006 | DALYs saved | 5,657 | 2,267,525 per year | ICER $ 401/ DALY saved | 411 | NR | NR | Changes in 'prevalence' of inadequate intakes used to calculate DALYs lost were extracted here. In all |

| Study ID* (Author Year) | INTERVENTION-Micronutrient(s) | INTERVENTION-Food Vehicle(s) | Dose | Other Specs | Compar-ator | Country | Curr-ency | Price Year | HEALTH Outcome Type | Health Result - Difference | Costs - Difference *as REPORTED* | Incremental CE Ratio *as REPORTED* | CONVERTED ICER (using 2022 US$)† | Cost-Benefit Analysis | Dis-count Rate | Uncertainty & Sensitivity Analyses |
|---|---|---|---|---|---|---|---|---|---|---|---|---|---|---|---|---|
| | | | Vitamin B2 (2.5 mg/kg) + Vitamin B3 (20.0 mg/kg) + Vitamin B6 (2.5 mg/kg) + Folic acid (1.0 mg/kg) + Vitamin B12 (0.005 mg/kg) + Iron (10.0 mg/kg) + Zinc (15.0 mg/kg) | roller meal | | | | | | | | | | | | instances, "efficiency-based" estimates were more cost-effective than prevalence-based ICERs. |
| Fiedler 2013 | Vitamins A + B1 + B2 + B3 + B6 + B12 + Folic acid + Iron + Zinc | Sugar + Maize meal | Sugar: Vitamin A (palmitate 250 CWS/CWD : minimum 10 mg /kg); Maize meal: Vitamin A (retinol 3333IU/kg) + Vitamin B1 (2.0 mg/kg) + Vitamin B2 (2.5 mg/kg) + Vitamin B3 (20.0 mg/kg) + Vitamin B6 (2.5 mg/kg) + Folic acid (1.0 mg/kg) + Vitamin B12 (0.005 mg/kg) + Iron (10.0 mg/kg) + Zinc (15.0 mg/kg) | Maize as breakfast meal and roller meal | Unfortified food | Zambia | US$ | 2006 | DALYs saved | 14,864 | 3,140,590 per year | ICER $ 211/ DALY saved | 216 | NR | NR | Changes in 'prevalence' of inadequate intakes used to calculate DALYs lost were extracted here. In all instances, "efficiency-based" estimates were more cost-effective than prevalence-based ICERs. |

| Study ID* (Author Year) | INTERVENTION- Micronutrient(s) | INTERVENTION- Food Vehicle(s) | Dose | Other Specs | Compar-ator | Country | Curr-ency | Price Year | HEALTH Outcome Type | Health Result - Difference | Costs - Difference *as REPORTED* | Incremental CE Ratio *as REPORTED* | CONVERTED ICER (using 2022 US$)† | Cost-Benefit Analysis | Dis-count Rate | Uncertainty & Sensitivity Analyses |
|---|---|---|---|---|---|---|---|---|---|---|---|---|---|---|---|---|
| Fiedler 2013 | Vitamins A + B1 + B2 + B3 + B6 + B12 + Folic acid + Iron + Zinc | Sugar + Vegetable oil + Maize meal | Sugar: Vitamin A (palmitate 250 CWS/CWD : minimum 10 mg /kg); Vegetable oil: Vitamin A (retinol 30 mg/kg); Maize meal: Vitamin A (retinol 3333IU/kg) + Vitamin B1 (2.0 mg/kg) + Vitamin B2 (2.5 mg/kg) + Vitamin B3 (20.0 mg/kg) + Vitamin B6 (2.5 mg/kg) + Folic acid (1.0 mg/kg) + Vitamin B12 (0.005 mg/kg) + Iron (10.0 mg/kg) + Zinc (15.0 mg/kg) | Maize as breakfast meal and roller meal | Unfortified food | Zambia | US$ | 2006 | DALYs saved | 32,225 | 3,382,990 per year | ICER $ 105/ DALY saved | 108 | NR | NR | Changes in 'prevalence' of inadequate intakes used to calculate DALYs lost were extracted here. In all instances, "efficiency-based" estimates were more cost-effective than prevalence-based ICERs. |
| Fiedler 2009 | Vitamins A + B1 + B2 + B3 + B6 + B12 + Folic acid + Iron + Zinc | Wheat flour | Vitamin A (Retinyl palmitate 250,000 IU/g) 1.5-4.0 mg RE/kg; Vitamin B1 2-6 mg/kg ; Vitamin B2 3-5 mg/kg; | Industrial | Pre-fortification | Zambia | US$ | 2009 | DALY saved | NR | 1.9 million | 12.10 | 13 | NR | 3% for DALYs; 0% for costs | NR |

| Study ID* (Author Year) | INTERVENTION-Micronutrient(s) | INTERVENTION-Food Vehicle(s) | Dose | Other Specs | Compar-ator | Country | Curr-ency | Price Year | HEALTH Outcome Type | Health Result - Difference | Costs - Difference *as REPORTED* | Incremental CE Ratio *as REPORTED* | CONVERTED ICER (using 2022 US$)† | Cost-Benefit Analysis | Dis-count Rate | Uncertainty & Sensitivity Analyses |
|---|---|---|---|---|---|---|---|---|---|---|---|---|---|---|---|---|
| | | | Vitamin B3 5-60 mg/kg; Vitamin B6 2-6 mg/kg; Folic acid 1.5-4 mg/kg; Vitamin B12 0.005-0.020; Iron NaFeEDTA 20 mg/kg or Ferrous fumarate 45 mg/kg; Zinc oxide 10-30 mg/kg | | | | | | | | | | | | | |
| Fiedler 2009 | Vitamins A + B1 + B2 + B3 + B6 + B12 + Folic acid + Iron + Zinc | Maize flour | Vitamin A (Retinyl palmitate 250,000 IU/g) 1.5-4.0 mg RE/kg; Vitamin B1 2-6 mg/kg ; Vitamin B2 3-5 mg/kg; Vitamin B3 5-60 mg/kg; Vitamin B6 2-6 mg/kg; Folic acid 1.5-4 mg/kg; Vitamin B12 0.005-0.020; Iron NaFeEDTA 20 mg/kg or Ferrous fumarate 45 mg/kg; Zinc oxide | Industrial | Pre-fortification | Zimbabwe | US$ | 2009 | DALY saved | NR | 82.9 million | 1060 | 1,240 | NR | 3% for DALYs; 0% for costs | NR |

| Study ID* (Author Year) | INTERVENTION-Micronutrient(s) | INTERVENTION-Food Vehicle(s) | Dose | Other Specs | Compar-ator | Country | Curr-ency | Price Year | HEALTH Outcome Type | Health Result - Difference | Costs - Difference *as REPORTED* | Incremental CE Ratio *as REPORTED* | CONVERTED ICER (using 2022 US$)† | Cost-Benefit Analysis | Dis-count Rate | Uncertainty & Sensitivity Analyses |
|---|---|---|---|---|---|---|---|---|---|---|---|---|---|---|---|---|
| | | | 10-30 mg/kg | | | | | | | | | | | | | |
| Fiedler 2009 | Vitamins A + B1 + B2 + B3 + B6 + B12 + Folic acid + Iron + Zinc | Wheat flour | Vitamin A (Retinyl palmitate 250,000 IU/g) 1.5-4.0 mg RE/kg; Vitamin B1 2-6 mg/kg ; Vitamin B2 3-5 mg/kg; Vitamin B3 5-60 mg/kg; Vitamin B6 2-6 mg/kg; Folic acid 1.5-4 mg/kg; Vitamin B12 0.005-0.020; Iron NaFeEDTA 20 mg/kg or Ferrous fumarate 45 mg/kg; Zinc oxide 10-30 mg/kg | Industrial | Pre-fortification | Zimbabwe | US$ | 2009 | DALY saved | NR | 1.8 million | 31.78 | 37 | NR | 3% for DALYs; 0% for costs | NR |
| Fiedler 2012 | Vitamins A + B1 + B2 + B3 + C + Folic acid + Calcium + Iron | Wheat flour (in Gujarat's Integrated Child Development Scheme) | Per 50 g flour (2011 levels): Vit. A (mcg) 150, Iron (mg) 7.5, Calcium (mg) 250, Vit. B2 (mg) 0.33, Vit. C (mg) 10, Folic acid (mcg) 50, Vit. B3 (mg) 3.5, | NR | Unfortified food | India | US$ | Unclear | DALYs saved | 27,427 | 605,616 annually | Cost per DALY averted US$ 22.1 | 24 | NR | NR | Estimate for the 3 programs combined is $ 27.6 /DALY saved but this considers each of the SSNPs independently, not considering that some individuals may benefit from more than one of the programs. |

| Study ID* (Author Year) | INTERVENTION- Micronutrient(s) | INTERVENTION- Food Vehicle(s) | Dose | Other Specs | Compar-ator | Country | Curr-ency | Price Year | HEALTH Outcome Type | Health Result - Difference | Costs - Difference *as REPORTED* | Incremental CE Ratio *as REPORTED* | CONVERTED ICER (using 2022 US$)† | Cost-Benefit Analysis | Dis-count Rate | Uncertainty & Sensitivity Analyses |
|---|---|---|---|---|---|---|---|---|---|---|---|---|---|---|---|---|
| | | | Vit. B1 (mg) 0.3 | | | | | | | | | | | | | |
| Fiedler 2012 | Vitamins A + B1 + B2 + B3 + C + Folic acid + Calcium + Iron | Wheat flour (in Gujarat's Mid-Day Meal School Program) | Per 50 g flour (2011 levels): Vit. A (mcg) 150, Iron (mg) 7.5, Calcium (mg) 250, Vit. B2 (mg) 0.33, Vit. C (mg) 10, Folic acid (mcg) 50, Vit. B3 (mg) 3.5, Vit. B1 (mg) 0.3 | NR | Unfortified food | India | US$ | Unclea r | DALYs saved | 4,185 | 1,009,360 annually | Cost per DALY averted US$ 241.2 | 259 | NR | NR | Estimate for the 3 programs combined is $ 27.6 /DALY saved but this considers each of the SSNPs independently, not considering that some individuals may benefit from more than one of the programs. |
| Fiedler 2013 | Vitamins A + B1 + B2 + B3 + Calcium + Iron | Sugar + Vegetable oil + Wheat flour | Sugar: Vitamin A (palmitate 250 CWS/CWD : minimum 10 mg /kg); Vegetable oil: Vitamin A (retinol 30 mg/kg); Wheat Flour: Vitamin B1 (4.5-5.5 mg/kg) + Vitamin B2 (2.7-3.5 mg/kg) + Vitamin B3 (35.5-44.4 mg/kg) + Iron (28.9-36.7 mg/kg) + Calcium (1,111– | NR | Unfortified food | Zambia | US$ | 2006 | DALYs saved | 27,142 | 2,042,077 per year | ICER $ 75/ DALY saved | 77 | NR | NR | Changes in 'prevalence' of inadequate intakes used to calculate DALYs lost were extracted here. In all instances, "efficiency-based" estimates were more cost-effective than prevalence-based ICERs. |

| Study ID* (Author Year) | INTERVENTION-Micronutrient(s) | INTERVENTION-Food Vehicle(s) | Dose | Other Specs | Compar-ator | Country | Curr-ency | Price Year | HEALTH Outcome Type | Health Result - Difference | Costs - Difference *as REPORTED* | Incremental CE Ratio *as REPORTED* | CONVERTED ICER (using 2022 US$)† | Cost-Benefit Analysis | Dis-count Rate | Uncertainty & Sensitivity Analyses |
|---|---|---|---|---|---|---|---|---|---|---|---|---|---|---|---|---|
| | | | 1,444 mg/kg) | | | | | | | | | | | | | |
| Fiedler 2013 | Vitamins A + B1 + B2 + B3 + Calcium + Iron | Sugar + Wheat flour | Sugar: Vitamin A (palmitate 250 CWS/CWD : minimum 10 mg /kg); Wheat Flour: Vitamin B1 (4.5-5.5 mg/kg) + Vitamin B2 (2.7-3.5 mg/kg) + Vitamin B3 (35.5-44.4 mg/kg) + Iron (28.9-36.7 mg/kg) + Calcium (1,111–1,444 mg/kg) | NR | Unfortified food | Zambia | US$ | 2006 | DALYs saved | 8,262 | 1,799,677 per year | ICER $ 218/ DALY saved | 224 | NR | NR | Changes in 'prevalence' of inadequate intakes used to calculate DALYs lost were extracted here. In all instances, "efficiency-based" estimates were more cost-effective than prevalence-based ICERs. |
| Kakietek 2018 | Vitamins A + B1 + B2 + Folic acid + Iron + Zinc | Rice | NR | NR | Unfortified | Bangladesh | US$ | 2015 | DALYs averted | 188,218 | 74,391,248 | ICER $395 per DALY averted | 561 | NR | Unclea r | NR |
| Fiedler 2012 | Vitamins A + B2 + B12 + C + Folic acid + Calcium + Iodine + Iron + Zinc | Wheat flour (in Gujarat's Integrated Child Development Scheme) | Per 50 g flour (initial levels): Vit. (mcg) 200, Iron (mg) 7.5, Zinc (mg) 5, Calcium (mg) 225, Iodine (mcg) 50, Vit. B2 (mg) 0.5, Vit. C (mg) 20, Folic acid (mcg) | NR | Unfortified food | India | US$ | Unclea r | DALYs saved | 66,013 | 605,616 annually | Cost per DALY averted US$ 9.2 | 10 | NR | NR | Estimate for the 3 programs combined is $ 17.9 /DALY saved but this considers each of the SSNPs independently, not considering that some individuals may benefit from more than one of the programs. |

| Study ID* (Author Year) | INTERVENTION-Micronutrient(s) | INTERVENTION-Food Vehicle(s) | Dose | Other Specs | Compar-ator | Country | Curr-ency | Price Year | HEALTH Outcome Type | Health Result - Difference | Costs - Difference *as REPORTED* | Incremental CE Ratio *as REPORTED* | CONVERTED ICER (using 2022 US$)† | Cost-Benefit Analysis | Dis-count Rate | Uncertainty & Sensitivity Analyses |
|---|---|---|---|---|---|---|---|---|---|---|---|---|---|---|---|---|
| | | | 20, Vit. B12 (mcg) 0.5 | | | | | | | | | | | | | |
| Fiedler 2012 | Vitamins A + B2 + B12 + C + Folic acid + Calcium + Iodine + Iron + Zinc | Wheat flour (in Gujarat's Mid-Day Meal School Program) | Per 50 g flour (initial levels): Vit. (mcg) 200, Iron (mg) 7.5, Zinc (mg) 5, Calcium (mg) 225, Iodine (mcg) 50, Vit. B2 (mg) 0.5, Vit. C (mg) 20, Folic acid (mcg) 20, Vit. B12 (mcg) 0.5 | NR | Unfortified food | India | US$ | Unclear | DALYs saved | 4,185 | 1,009,360 annually | Cost per DALY averted US$ 241.2 | 259 | NR | NR | Estimate for the 3 programs combined is $ 17.9 /DALY saved but this considers each of the SSNPs independently, not considering that some individuals may benefit from more than one of the programs. |
| Mardones-Santander 1991 | Vitamins A + B3 + B6 + C + D + E + Folic acid + Copper + Iodine + Iron + Magnesium + Zinc | Powdered milk-based product | Additional amounts to Comparator (not total dose from milk; from 1988 cited article), per 100g: Vit. A 0.83 mg, Vit. B6 0.22mg, Vit. C 330mg, Vit. D (cholecalciferol) 0.0105mg, Vit. E 1.75mg, Vit. B3 (niacin) 2.47mg, Folic acid | NR | Unfortified milk powder containing 26% fat | Chile | US$ | 1989 | Infant mortality rate | -2.59% | -293,115 | Dominant | Dominant | NR | 10% | NR |

| Study ID* (Author Year) | INTERVENTION-Micronutrient(s) | INTERVENTION-Food Vehicle(s) | Dose | Other Specs | Compar-ator | Country | Curr-ency | Price Year | HEALTH Outcome Type | Health Result - Difference | Costs - Difference *as REPORTED* | Incremental CE Ratio *as REPORTED* | CONVERTED ICER (using 2022 US$)† | Cost-Benefit Analysis | Dis-count Rate | Uncertainty & Sensitivity Analyses |
|---|---|---|---|---|---|---|---|---|---|---|---|---|---|---|---|---|
| | | | 0.35g, Mg 25mg, Zn 3.0mg, Fe 42.5mg (providing 4.475mg bioavailabl e Fe), Cu 0.26mg, Iodine 0.015mg | | | | | | | | | | | | | |
| Johnson 2021 | Vitamins A + D | Vegetable oil | Vitamins A 20mg/100g, D3 0.0167 mg/100g | Retinyl Palmita te | No fortification | Ethiopia | US$ | NR | Benefit-cost ratio | NR | 21,351,611 | NR | N/A | BCR: 2.75 | NR | NR |
| Fiedler 2015 | Vitamin A + Iron + Zinc | Wheat flour | Fe: 55 mg/kg + Zn: 27 mg/kg + Vit. A: 4 IU/g | Iron as NaFeE DTA; + Zinc oxide + Vit. A (dry) 250,00 0 IU/g | No fortification | Bangladesh | US$ | 2012 (unclea r) | DALYs saved | 137,461 | 1,641,825 annually | ICER $ 11.94/ DALY saved | 21 | NR | Unclea r (costs & benefit s were discou nted) | NR |
| Fiedler 2015 | Vitamin A + Iron + Zinc | Wheat flour | Fe: 55 mg/kg + Zn: 27 mg/kg + Vit. A: 4 IU/g | Iron as Ferrous fumara te; + Zinc oxide + Vit. A (dry) 250,00 0 IU/g | No fortification | Bangladesh | US$ | 2012 (unclea r) | DALYs saved | 129,212 | 999,771 annually | ICER $ 7.74/ DALY saved | 14 | NR | Unclea r (costs & benefit s were discou nted) | NR |
| Fiedler 2015 | Vitamin A + Iron + Zinc | Wheat flour + Vegetable oil | Fe: 55 mg/kg + Zn: 27 mg/kg + Vit. A: flour (4 IU/g) & oil (15 mg/kg) | Iron as NaFeE DTA; + Zinc oxide + Vit. A: flour (250,0 00 IU/g, dry) & oil (retinyl | No fortification | Bangladesh | US$ | 2012 (unclea r) | DALYs saved | 479,898 | 2,909,941 annually | ICER $ 6.06/ DALY saved | 11 | NR | Unclea r (costs & benefit s were discou nted) | NR |

| Study ID* (Author Year) | INTERVENTION-Micronutrient(s) | INTERVENTION-Food Vehicle(s) | Dose | Other Specs | Compar-ator | Country | Curr-ency | Price Year | HEALTH Outcome Type | Health Result - Difference | Costs - Difference *as REPORTED* | Incremental CE Ratio *as REPORTED* | CONVERTED ICER (using 2022 US$)† | Cost-Benefit Analysis | Dis-count Rate | Uncertainty & Sensitivity Analyses |
|---|---|---|---|---|---|---|---|---|---|---|---|---|---|---|---|---|
| | | | | palmitate 1.7 mIU/g) | | | | | | | | | | | | |
| Fiedler 2015 | Vitamin A + Iron + Zinc | Wheat flour + Vegetable oil | Fe: 55 mg/kg + Zn: 27 mg/kg + Vit. A: flour (4 IU/g) & oil (15 mg/kg) | Iron as Ferrous fumarate; + Zinc oxide + Vit. A: flour (250,000 IU/g, dry) & oil (retinyl palmitate 1.7 mIU/g) | No fortification | Bangladesh | US$ | 2012 (unclear) | DALYs saved | 471,599 | 2,267,886 annually | ICER $ 4.81/ DALY saved | 9 | NR | Unclear (costs & benefits were discounted) | NR |
| Edejer 2005 | Vitamin A + Zinc | Sugar (VA) & Wheat (Zinc) | 80% coverage; dose NR | Zinc oxide | No fortification | Afr-E (sub-Saharan Africa) region | international dollars ($Int) | 2000 | DALYs averted yearly (millions) | 1.01 | 20,000,000 yearly | ICER $ 20/DALY saved | 102 | NR | 3% for both costs and benefits | Removal of age weighting and discounting for DALYs makes the interventions more cost effective. Varying geographical coverage from 80% to: 50% or 95% gave ICERs of I$ 24 or 19 per DALY saved (resp'ly). |
| Edejer 2005 | Vitamin A + Zinc | Sugar (VA) & Wheat (Zinc) | 80% coverage; dose NR | Zinc oxide | No fortification | Sear-D (South East Asia) region | international dollars ($Int) | 2000 | DALYs averted yearly (millions) | 1.20 | 42,000,000 yearly | ICER $ 35/DALY saved | 52 | NR | 3% for both costs and benefits | Removal of age weighting and discounting for DALYs makes the interventions more cost effective. Varying geographical coverage from 80% to: 50% or 95% gave ICERs of I$ 39 or 35 per DALY saved (resp'ly). |

| Study ID* (Author Year) | INTERVENTION-Micronutrient(s) | INTERVENTION-Food Vehicle(s) | Dose | Other Specs | Compar-ator | Country | Curr-ency | Price Year | HEALTH Outcome Type | Health Result - Difference | Costs - Difference *as REPORTED* | Incremental CE Ratio *as REPORTED* | CONVERTED ICER (using 2022 US$)† | Cost-Benefit Analysis | Dis-count Rate | Uncertainty & Sensitivity Analyses |
|---|---|---|---|---|---|---|---|---|---|---|---|---|---|---|---|---|
| Connelly 1996 | Vitamin B1 | Bread-flour | 0.64 mg/100 g | Milling | No fortification /Intervention | Australia | AUD Dollars (A$) | 1992 | Wernicke's encephalopathy cases averted | NR (Condition 2 assumption: No malabsorption of thiamin when consumed in bread) | 4,821,121 | ICER A$ 17,927/Case averted | 27,215 | NR | 10% | Varying discount rates from between 0% to 20% found results were similar. Using 20-year time horizon had very similar results. |
| Connelly 1996 | Vitamin B1 | Bread-flour | 0.64 mg/100 g | Milling | No fortification /Intervention | Australia | AUD Dollars (A$) | 1992 | Wernicke's encephalopathy cases averted | NR (Condition 3 assumption: Malabsorption of thiamin in heavy alcohol drinkers) | 4,821,121 | ICER A$ 35,855/Case averted | 54,431 | NR | 10% | Varying discount rates from between 0% to 20% found consistent results. |
| Asian Develop. Bank 2004 | Vitamin B1 + Folic acid + Calcium + Iron | Complementary food | Per 100 kcal: Calcium 40.00 mg; Iron 5.30 mg; Vitamin B1 0.08 mg; Folic Acid 11.00 mcg | Ground rice kernels | Unfortified food | Thailand | US$ | NR | Benefit-cost ratio | NR | 3,500,789 | NR | N/A | BCR: nearly 4 | NR [Discounted costs and benefits] | NR |
| Johnson 2021 | Vitamins B1 + B2 + B3 + B6 + B12 + Folic acid + Zinc | Wheat flour | Vitamins 9 (Folate) 2.0 ppm, B1 9.0 ppm, B26.0 ppm, B350.0 ppm, B6 6.0 ppm, B12 0.02 ppm, Zinc 80 ppm | Folic acid, Thiamine Mononitrate, Riboflavin, Niacinamide, Pyridoxine, Vitamin B12, Zinc Oxide | No fortification | Ethiopia | US$ | NR | Benefit-cost ratio | NR | 36,200,000 | NR | N/A | BCR: 13.0 | NR | NR |

| Study ID* (Author Year) | INTERVENTION-Micronutrient(s) | INTERVENTION-Food Vehicle(s) | Dose | Other Specs | Compar-ator | Country | Curr-ency | Price Year | HEALTH Outcome Type | Health Result - Difference | Costs - Difference *as REPORTED* | Incremental CE Ratio *as REPORTED* | CONVERTED ICER (using 2022 US$)† | Cost-Benefit Analysis | Dis-count Rate | Uncertainty & Sensitivity Analyses |
|---|---|---|---|---|---|---|---|---|---|---|---|---|---|---|---|---|
| Fiedler 2013 | Vitamins B1 + B2 + B3 + Calcium + Iron | Wheat flour | Vitamin B1 (4.5-5.5 mg/kg) + Vitamin B2 (2.7-3.5 mg/kg) + Vitamin B3 (35.5-44.4 mg/kg) + Iron (28.9-36.7 mg/kg) + Calcium (1,111–1,444 mg/kg) | NR | Unfortified food | Zambia | US$ | 2006 | DALYs saved | 926 | 926,612 per year | ICER $ 1,001/ DALY saved | 1,026 | NR | NR | Changes in 'prevalence' of inadequate intakes used to calculate DALYs lost were extracted here. In all instances, "efficiency-based" estimates were more cost-effective than prevalence-based ICERs. |
| Niemesh 2015 | Vitamins B1 + B3 + Iron | Bread | 24 mg iron + niacin + 2.5 mg thiamin per pound flour | NR | Pre-mandatory fortification | USA | US$ | 1940 | Benefit-cost ratio | NR | 35–50 cents per person annually | NR | N/A | BCR: 14.1 | NR | NR |
| Fiedler 2009 | Vitamin B12 + Folic acid + Iron | Wheat flour | Folic acid 1.5-4.0 mg/kg; Vitamin B12 (0.1% water soluble) 0.005-0.020 [mg/kg or ppm?]; Iron NaFeEDTA 20 mg/kg or Ferrous fumarate 45 mg/kg | Industrial | Pre-fortification | Afghanistan | US$ | 2009 | DALY saved | NR | 1.4 million | 48.80 | 52 | NR | 3% for DALYs; 0% for costs | NR |
| Fiedler 2009 | Vitamin B12 + Folic acid + Iron | Maize flour | Folic acid 1.5-4.0 mg/kg; Vitamin B12 (0.1% water soluble) 0.005-0.020 [mg/kg or ppm?]; Iron NaFeEDTA | Industrial | Pre-fortification | Angola | US$ | 2009 | DALY saved | NR | 43.8 million | 6064 | 8,184 | NR | 3% for DALYs; 0% for costs | NR |

| Study ID* (Author Year) | INTERVENTION-Micronutrient(s) | INTERVENTION-Food Vehicle(s) | Dose | Other Specs | Compar-ator | Country | Curr-ency | Price Year | HEALTH Outcome Type | Health Result - Difference | Costs - Difference *as REPORTED* | Incremental CE Ratio *as REPORTED* | CONVERTED ICER (using 2022 US$)† | Cost-Benefit Analysis | Dis-count Rate | Uncertainty & Sensitivity Analyses |
|---|---|---|---|---|---|---|---|---|---|---|---|---|---|---|---|---|
| | | | 20 mg/kg or Ferrous fumarate 45 mg/kg | | | | | | | | | | | | | |
| Fiedler 2009 | Vitamin B12 + Folic acid + Iron | Wheat flour | Folic acid 1.5-4.0 mg/kg; Vitamin B12 (0.1% water soluble) 0.005-0.020 [mg/kg or ppm?]; Iron NaFeEDTA 20 mg/kg or Ferrous fumarate 45 mg/kg | Industrial | Pre-fortification | Angola | US$ | 2009 | DALY saved | NR | 1.4 million | 34.12 | 46 | NR | 3% for DALYs; 0% for costs | NR |
| Fiedler 2009 | Vitamin B12 + Folic acid + Iron | Wheat flour | Folic acid 1.5-4.0 mg/kg; Vitamin B12 (0.1% water soluble) 0.005-0.020 [mg/kg or ppm?]; Iron NaFeEDTA 20 mg/kg or Ferrous fumarate 45 mg/kg | Industrial | Pre-fortification | Bangladesh | US$ | 2009 | DALY saved | NR | 9.9 million | 48.36 | 91 | NR | 3% for DALYs; 0% for costs | NR |
| Fiedler 2009 | Vitamin B12 + Folic acid + Iron | Maize flour | Folic acid 1.5-4.0 mg/kg; Vitamin B12 (0.1% water soluble) 0.005-0.020 [mg/kg or ppm?]; Iron NaFeEDTA 20 mg/kg or Ferrous | Industrial | Pre-fortification | Bolivia | US$ | 2009 | DALY saved | NR | 89.6 million | 9161 | 14,723 | NR | 3% for DALYs; 0% for costs | NR |

| Study ID* (Author Year) | INTERVENTION-Micronutrient(s) | INTERVENTION-Food Vehicle(s) | Dose | Other Specs | Compar-ator | Country | Curr-ency | Price Year | HEALTH Outcome Type | Health Result - Difference | Costs - Difference *as REPORTED* | Incremental CE Ratio *as REPORTED* | CONVERTED ICER (using 2022 US$)† | Cost-Benefit Analysis | Dis-count Rate | Uncertainty & Sensitivity Analyses |
|---|---|---|---|---|---|---|---|---|---|---|---|---|---|---|---|---|
| | | | fumarate 45 mg/kg | | | | | | | | | | | | | |
| Fiedler 2009 | Vitamin B12 + Folic acid + Iron | Wheat flour | Folic acid 1.5-4.0 mg/kg; Vitamin B12 (0.1% water soluble) 0.005-0.020 [mg/kg or ppm?]; Iron NaFeEDTA 20 mg/kg or Ferrous fumarate 45 mg/kg | Industrial | Pre-fortification | Brazil | US$ | 2009 | DALY saved | NR | 10.2 million | 40.55 | 39 | NR | 3% for DALYs; 0% for costs | NR |
| Fiedler 2009 | Vitamin B12 + Folic acid + Iron | Wheat flour | Folic acid 1.5-4.0 mg/kg; Vitamin B12 (0.1% water soluble) 0.005-0.020 [mg/kg or ppm?]; Iron NaFeEDTA 20 mg/kg or Ferrous fumarate 45 mg/kg | Industrial | Pre-fortification | Burkina Faso | US$ | 2009 | DALY saved | NR | 1.3 million | 16.41 | 16 | NR | 3% for DALYs; 0% for costs | NR |
| Fiedler 2009 | Vitamin B12 + Folic acid + Iron | Wheat flour | Folic acid 1.5-4.0 mg/kg; Vitamin B12 (0.1% water soluble) 0.005-0.020 [mg/kg or ppm?]; Iron NaFeEDTA 20 mg/kg or Ferrous fumarate 45 mg/kg | Industrial | Pre-fortification | Cambodia | US$ | 2009 | DALY saved | NR | 1.2 million | 52.61 | 72 | NR | 3% for DALYs; 0% for costs | NR |

| Study ID* (Author Year) | INTERVENTION-Micronutrient(s) | INTERVENTION-Food Vehicle(s) | Dose | Other Specs | Compar-ator | Country | Curr-ency | Price Year | HEALTH Outcome Type | Health Result - Difference | Costs - Difference *as REPORTED* | Incremental CE Ratio *as REPORTED* | CONVERTED ICER (using 2022 US$)† | Cost-Benefit Analysis | Dis-count Rate | Uncertainty & Sensitivity Analyses |
|---|---|---|---|---|---|---|---|---|---|---|---|---|---|---|---|---|
| Fiedler 2009 | Vitamin B12 + Folic acid + Iron | Wheat flour | Folic acid 1.5-4.0 mg/kg; Vitamin B12 (0.1% water soluble) 0.005-0.020 [mg/kg or ppm?]; Iron NaFeEDTA 20 mg/kg or Ferrous fumarate 45 mg/kg | Industrial | Pre-fortification | Cameroon | US$ | 2009 | DALY saved | NR | 1.1 million | 22.51 | 22 | NR | 3% for DALYs; 0% for costs | NR |
| Fiedler 2009 | Vitamin B12 + Folic acid + Iron | Wheat flour | Folic acid 1.5-4.0 mg/kg; Vitamin B12 (0.1% water soluble) 0.005-0.020 [mg/kg or ppm?]; Iron NaFeEDTA 20 mg/kg or Ferrous fumarate 45 mg/kg | Industrial | Pre-fortification | China | US$ | 2009 | DALY saved | NR | 513.7 million | 412.42 | 607 | NR | 3% for DALYs; 0% for costs | NR |
| Fiedler 2009 | Vitamin B12 + Folic acid + Iron | Wheat flour | Folic acid 1.5-4.0 mg/kg; Vitamin B12 (0.1% water soluble) 0.005-0.020 [mg/kg or ppm?]; Iron NaFeEDTA 20 mg/kg or Ferrous fumarate 45 mg/kg | Industrial | Pre-fortification | Congo, Dem. Rep. | US$ | 2009 | DALY saved | NR | 1.2 million | 2.90 | 5 | NR | 3% for DALYs; 0% for costs | NR |
| Fiedler 2009 | Vitamin B12 + Folic acid + Iron | Maize flour | Folic acid 1.5-4.0 | Industrial | Pre-fortification | Côte d'Ivoire | US$ | 2009 | DALY saved | NR | 37.1 million | 791 | 774 | NR | 3% for DALY | NR |

| Study ID* (Author Year) | INTERVENTION-Micronutrient(s) | INTERVENTION-Food Vehicle(s) | Dose | Other Specs | Compar-ator | Country | Curr-ency | Price Year | HEALTH Outcome Type | Health Result - Difference | Costs - Difference *as REPORTED* | Incremental CE Ratio *as REPORTED* | CONVERTED ICER (using 2022 US$)† | Cost-Benefit Analysis | Dis-count Rate | Uncertainty & Sensitivity Analyses |
|---|---|---|---|---|---|---|---|---|---|---|---|---|---|---|---|---|
| | | | mg/kg; Vitamin B12 (0.1% water soluble) 0.005-0.020 [mg/kg or ppm?]; Iron NaFeEDTA 20 mg/kg or Ferrous fumarate 45 mg/kg | | | | | | | | | | | | s; 0% for costs | |
| Fiedler 2009 | Vitamin B12 + Folic acid + Iron | Wheat flour | Folic acid 1.5-4.0 mg/kg; Vitamin B12 (0.1% water soluble) 0.005-0.020 [mg/kg or ppm?]; Iron NaFeEDTA 20 mg/kg or Ferrous fumarate 45 mg/kg | Industrial | Pre-fortification | Côte d'Ivoire | US$ | 2009 | DALY saved | NR | 1.3 million | 10.37 | 10 | NR | 3% for DALYs; 0% for costs | NR |
| Fiedler 2009 | Vitamin B12 + Folic acid + Iron | Wheat flour | Folic acid 1.5-4.0 mg/kg; Vitamin B12 (0.1% water soluble) 0.005-0.020 [mg/kg or ppm?]; Iron NaFeEDTA 20 mg/kg or Ferrous fumarate 45 mg/kg | Industrial | Pre-fortification | Ethiopia | US$ | 2009 | DALY saved | NR | 5.5 million | 60.45 | 84 | NR | 3% for DALYs; 0% for costs | NR |
| Fiedler 2009 | Vitamin B12 + Folic acid + Iron | Maize flour | Folic acid 1.5-4.0 mg/kg; Vitamin | Industrial | Pre-fortification | Ghana | US$ | 2009 | DALY saved | NR | 57.1 million | 1314 | 1,783 | NR | 3% for DALYs; 0% | NR |

| Study ID* (Author Year) | INTERVENTION-Micronutrient(s) | INTERVENTION-Food Vehicle(s) | Dose | Other Specs | Compar-ator | Country | Curr-ency | Price Year | HEALTH Outcome Type | Health Result - Difference | Costs - Difference *as REPORTED* | Incremental CE Ratio *as REPORTED* | CONVERTED ICER (using 2022 US$)† | Cost-Benefit Analysis | Dis-count Rate | Uncertainty & Sensitivity Analyses |
|---|---|---|---|---|---|---|---|---|---|---|---|---|---|---|---|---|
| | | | B12 (0.1% water soluble) 0.005-0.020 [mg/kg or ppm?]; Iron NaFeEDTA 20 mg/kg or Ferrous fumarate 45 mg/kg | | | | | | | | | | | | for costs | |
| Fiedler 2009 | Vitamin B12 + Folic acid + Iron | Wheat flour | Folic acid 1.5-4.0 mg/kg; Vitamin B12 (0.1% water soluble) 0.005-0.020 [mg/kg or ppm?]; Iron NaFeEDTA 20 mg/kg or Ferrous fumarate 45 mg/kg | Industrial | Pre-fortification | Ghana | US$ | 2009 | DALY saved | NR | 1.6 million | 11.35 | 15 | NR | 3% for DALYs; 0% for costs | NR |
| Fiedler 2009 | Vitamin B12 + Folic acid + Iron | Maize flour | Folic acid 1.5-4.0 mg/kg; Vitamin B12 (0.1% water soluble) 0.005-0.020 [mg/kg or ppm?]; Iron NaFeEDTA 20 mg/kg or Ferrous fumarate 45 mg/kg | Industrial | Pre-fortification | Guatemala | US$ | 2009 | DALY saved | NR | 319.4 million | 14525 | 23,763 | NR | 3% for DALYs; 0% for costs | NR |
| Fiedler 2009 | Vitamin B12 + Folic acid + Iron | Wheat flour | Folic acid 1.5-4.0 mg/kg; Vitamin B12 (0.1% water | Industrial | Pre-fortification | Guatemala | US$ | 2009 | DALY saved | NR | 1.9 million | 49.48 | 81 | NR | 3% for DALYs; 0% for costs | NR |

| Study ID* (Author Year) | INTERVENTION-Micronutrient(s) | INTERVENTION-Food Vehicle(s) | Dose | Other Specs | Compar-ator | Country | Curr-ency | Price Year | HEALTH Outcome Type | Health Result - Difference | Costs - Difference *as REPORTED* | Incremental CE Ratio *as REPORTED* | CONVERTED ICER (using 2022 US$)† | Cost-Benefit Analysis | Dis-count Rate | Uncertainty & Sensitivity Analyses |
|---|---|---|---|---|---|---|---|---|---|---|---|---|---|---|---|---|
| | | | soluble) 0.005-0.020 [mg/kg or ppm?]; Iron NaFeEDTA 20 mg/kg or Ferrous fumarate 45 mg/kg | | | | | | | | | | | | | |
| Fiedler 2009 | Vitamin B12 + Folic acid + Iron | Wheat flour | Folic acid 1.5-4.0 mg/kg; Vitamin B12 (0.1% water soluble) 0.005-0.020 [mg/kg or ppm?]; Iron NaFeEDTA 20 mg/kg or Ferrous fumarate 45 mg/kg | Industrial | Pre-fortification | Guinea | US$ | 2009 | DALY saved | NR | 1.1 million | 18.57 | 21 | NR | 3% for DALYs; 0% for costs | NR |
| Fiedler 2009 | Vitamin B12 + Folic acid + Iron | Wheat flour | Folic acid 1.5-4.0 mg/kg; Vitamin B12 (0.1% water soluble) 0.005-0.020 [mg/kg or ppm?]; Iron NaFeEDTA 20 mg/kg or Ferrous fumarate 45 mg/kg | Industrial | Pre-fortification | India | US$ | 2009 | DALY saved | NR | 85.7 million | 26.87 | 34 | NR | 3% for DALYs; 0% for costs | NR |
| Qureshy 2023 | Vitamin B12 + Folic acid + Iron | Rice | Iron 3.5 mg/100 g of rice (B12 & folate doses NR) | Ferric pyrophosphate | Unfortified food | India | US$ | NR | Benefit-cost ratio | NR | 3,300,000,000 | NR | N/A | BCR: 8.2 | 3% | Alternative scenarios changing key model parameter values revealed conclusions remained robust with positive BCRs. For each program |

| Study ID* (Author Year) | INTERVENTION-Micronutrient(s) | INTERVENTION-Food Vehicle(s) | Dose | Other Specs | Compar-ator | Country | Curr-ency | Price Year | HEALTH Outcome Type | Health Result - Difference | Costs - Difference *as REPORTED* | Incremental CE Ratio *as REPORTED* | CONVERTED ICER (using 2022 US$)[†] | Cost-Benefit Analysis | Dis-count Rate | Uncertainty & Sensitivity Analyses |
|---|---|---|---|---|---|---|---|---|---|---|---|---|---|---|---|---|
| | | | | | | | | | | | | | | | | individually, the BCRs were: 11.7 (MDM); and 3.7 (PDS). |
| Fiedler 2009 | Vitamin B12 + Folic acid + Iron | Wheat flour | Folic acid 1.5-4.0 mg/kg; Vitamin B12 (0.1% water soluble) 0.005-0.020 [mg/kg or ppm?]; Iron NaFeEDTA 20 mg/kg or Ferrous fumarate 45 mg/kg | Industrial | Pre-fortification | Indonesia | US$ | 2009 | DALY saved | NR | 1.9 million | 7.62 | 10 | NR | 3% for DALYs; 0% for costs | NR |
| Fiedler 2009 | Vitamin B12 + Folic acid + Iron | Maize flour | Folic acid 1.5-4.0 mg/kg; Vitamin B12 (0.1% water soluble) 0.005-0.020 [mg/kg or ppm?]; Iron NaFeEDTA 20 mg/kg or Ferrous fumarate 45 mg/kg | Industrial | Pre-fortification | Kenya | US$ | 2009 | DALY saved | NR | 146.4 million | 1796 | 2,618 | NR | 3% for DALYs; 0% for costs | NR |
| Fiedler 2009 | Vitamin B12 + Folic acid + Iron | Wheat flour | Folic acid 1.5-4.0 mg/kg; Vitamin B12 (0.1% water soluble) 0.005-0.020 [mg/kg or ppm?]; Iron NaFeEDTA 20 mg/kg or Ferrous | Industrial | Pre-fortification | Kenya | US$ | 2009 | DALY saved | NR | 1.5 million | 14.26 | 21 | NR | 3% for DALYs; 0% for costs | NR |

| Study ID* (Author Year) | INTERVENTION-Micronutrient(s) | INTERVENTION-Food Vehicle(s) | Dose | Other Specs | Compar-ator | Country | Curr-ency | Price Year | HEALTH Outcome Type | Health Result - Difference | Costs - Difference *as REPORTED* | Incremental CE Ratio *as REPORTED* | CONVERTED ICER (using 2022 US$)[†] | Cost-Benefit Analysis | Dis-count Rate | Uncertainty & Sensitivity Analyses |
|---|---|---|---|---|---|---|---|---|---|---|---|---|---|---|---|---|
| | | | fumarate 45 mg/kg | | | | | | | | | | | | | |
| Fiedler 2009 | Vitamin B12 + Folic acid + Iron | Wheat flour | Folic acid 1.5-4.0 mg/kg; Vitamin B12 (0.1% water soluble) 0.005-0.020 [mg/kg or ppm?]; Iron NaFeEDTA 20 mg/kg or Ferrous fumarate 45 mg/kg | Industrial | Pre-fortification | Madagascar | US$ | 2009 | DALY saved | NR | 1.2 million | 16.21 | 19 | NR | 3% for DALYs; 0% for costs | NR |
| Fiedler 2009 | Vitamin B12 + Folic acid + Iron | Maize flour | Folic acid 1.5-4.0 mg/kg; Vitamin B12 (0.1% water soluble) 0.005-0.020 [mg/kg or ppm?]; Iron NaFeEDTA 20 mg/kg or Ferrous fumarate 45 mg/kg | Industrial | Pre-fortification | Malawi | US$ | 2009 | DALY saved | NR | 29.4 million | 183 | 158 | NR | 3% for DALYs; 0% for costs | NR |
| Fiedler 2009 | Vitamin B12 + Folic acid + Iron | Wheat flour | Folic acid 1.5-4.0 mg/kg; Vitamin B12 (0.1% water soluble) 0.005-0.020 [mg/kg or ppm?]; Iron NaFeEDTA 20 mg/kg or Ferrous fumarate 45 mg/kg | Industrial | Pre-fortification | Malawi | US$ | 2009 | DALY saved | NR | 1.3 million | 24.61 | 21 | NR | 3% for DALYs; 0% for costs | NR |

| Study ID* (Author Year) | INTERVENTION-Micronutrient(s) | INTERVENTION-Food Vehicle(s) | Dose | Other Specs | Compar-ator | Country | Curr-ency | Price Year | HEALTH Outcome Type | Health Result - Difference | Costs - Difference *as REPORTED* | Incremental CE Ratio *as REPORTED* | CONVERTED ICER (using 2022 US$)† | Cost-Benefit Analysis | Dis-count Rate | Uncertainty & Sensitivity Analyses |
|---|---|---|---|---|---|---|---|---|---|---|---|---|---|---|---|---|
| Fiedler 2009 | Vitamin B12 + Folic acid + Iron | Wheat flour | Folic acid 1.5-4.0 mg/kg; Vitamin B12 (0.1% water soluble) 0.005-0.020 [mg/kg or ppm?]; Iron NaFeEDTA 20 mg/kg or Ferrous fumarate 45 mg/kg | Industrial | Pre-fortification | Mali | US$ | 2009 | DALY saved | NR | 1.3 million | 17.09 | 19 | NR | 3% for DALYs; 0% for costs | NR |
| Fiedler 2009 | Vitamin B12 + Folic acid + Iron | Maize flour | Folic acid 1.5-4.0 mg/kg; Vitamin B12 (0.1% water soluble) 0.005-0.020 [mg/kg or ppm?]; Iron NaFeEDTA 20 mg/kg or Ferrous fumarate 45 mg/kg | Industrial | Pre-fortification | Mexico | US$ | 2009 | DALY saved | NR | 3397.7 million | 23170 | 28,474 | NR | 3% for DALYs; 0% for costs | NR |
| Fiedler 2009 | Vitamin B12 + Folic acid + Iron | Wheat flour | Folic acid 1.5-4.0 mg/kg; Vitamin B12 (0.1% water soluble) 0.005-0.020 [mg/kg or ppm?]; Iron NaFeEDTA 20 mg/kg or Ferrous fumarate 45 mg/kg | Industrial | Pre-fortification | Mexico | US$ | 2009 | DALY saved | NR | 11.3 million | 76.94 | 95 | NR | 3% for DALYs; 0% for costs | NR |
| Fiedler 2009 | Vitamin B12 + Folic acid + Iron | Wheat flour | Folic acid 1.5-4.0 | Industrial | Pre-fortification | Mozambique | US$ | 2009 | DALY saved | NR | 1.7 million | 33.31 | 27 | NR | 3% for DALY | NR |

| Study ID* (Author Year) | INTERVENTION-Micronutrient(s) | INTERVENTION-Food Vehicle(s) | Dose | Other Specs | Compar-ator | Country | Curr-ency | Price Year | HEALTH Outcome Type | Health Result - Difference | Costs - Difference *as REPORTED* | Incremental CE Ratio *as REPORTED* | CONVERTED ICER (using 2022 US$)† | Cost-Benefit Analysis | Dis-count Rate | Uncertainty & Sensitivity Analyses |
|---|---|---|---|---|---|---|---|---|---|---|---|---|---|---|---|---|
| | | | mg/kg; Vitamin B12 (0.1% water soluble) 0.005-0.020 [mg/kg or ppm?]; Iron NaFeEDTA 20 mg/kg or Ferrous fumarate 45 mg/kg | | | | | | | | | | | | s; 0% for costs | |
| Fiedler 2009 | Vitamin B12 + Folic acid + Iron | Wheat flour | Folic acid 1.5-4.0 mg/kg; Vitamin B12 (0.1% water soluble) 0.005-0.020 [mg/kg or ppm?]; Iron NaFeEDTA 20 mg/kg or Ferrous fumarate 45 mg/kg | Industrial | Pre-fortification | Myanmar | US$ | 2009 | DALY saved | NR | 1.2 million | 4.34 | 1 | NR | 3% for DALYs; 0% for costs | NR |
| Fiedler 2009 | Vitamin B12 + Folic acid + Iron | Wheat flour | Folic acid 1.5-4.0 mg/kg; Vitamin B12 (0.1% water soluble) 0.005-0.020 [mg/kg or ppm?]; Iron NaFeEDTA 20 mg/kg or Ferrous fumarate 45 mg/kg | Industrial | Pre-fortification | Nepal | US$ | 2009 | DALY saved | NR | 2.1 million | 13.65 | 24 | NR | 3% for DALYs; 0% for costs | NR |
| Fiedler 2009 | Vitamin B12 + Folic acid + Iron | Wheat flour | Folic acid 1.5-4.0 mg/kg; Vitamin | Industrial | Pre-fortification | Niger | US$ | 2009 | DALY saved | NR | 1.3 million | 28.90 | 28 | NR | 3% for DALYs; 0% | NR |

| Study ID* (Author Year) | INTERVENTION-Micronutrient(s) | INTERVENTION-Food Vehicle(s) | Dose | Other Specs | Compar-ator | Country | Curr-ency | Price Year | HEALTH Outcome Type | Health Result - Difference | Costs - Difference *as REPORTED* | Incremental CE Ratio *as REPORTED* | CONVERTED ICER (using 2022 US$)† | Cost-Benefit Analysis | Dis-count Rate | Uncertainty & Sensitivity Analyses |
|---|---|---|---|---|---|---|---|---|---|---|---|---|---|---|---|---|
| | | | B12 (0.1% water soluble) 0.005-0.020 [mg/kg or ppm?]; Iron NaFeEDTA 20 mg/kg or Ferrous fumarate 45 mg/kg | | | | | | | | | | | | for costs | |
| Fiedler 2009 | Vitamin B12 + Folic acid + Iron | Wheat flour | Folic acid 1.5-4.0 mg/kg; Vitamin B12 (0.1% water soluble) 0.005-0.020 [mg/kg or ppm?]; Iron NaFeEDTA 20 mg/kg or Ferrous fumarate 45 mg/kg | Industrial | Pre-fortification | Nigeria | US$ | 2009 | DALY saved | NR | 2.0 million | 2.36 | 3 | NR | 3% for DALYs; 0% for costs | NR |
| Fiedler 2009 | Vitamin B12 + Folic acid + Iron | Wheat flour | Folic acid 1.5-4.0 mg/kg; Vitamin B12 (0.1% water soluble) 0.005-0.020 [mg/kg or ppm?]; Iron NaFeEDTA 20 mg/kg or Ferrous fumarate 45 mg/kg | Industrial | Pre-fortification | Pakistan | US$ | 2009 | DALY saved | NR | 46.8 million | 340.81 | 380 | NR | 3% for DALYs; 0% for costs | NR |
| Fiedler 2009 | Vitamin B12 + Folic acid + Iron | Wheat flour | Folic acid 1.5-4.0 mg/kg; Vitamin B12 (0.1% water | Industrial | Pre-fortification | Peru | US$ | 2009 | DALY saved | NR | 1.7 million | 34.42 | 43 | NR | 3% for DALYs; 0% for costs | NR |

| Study ID* (Author Year) | INTERVENTION-Micronutrient(s) | INTERVENTION-Food Vehicle(s) | Dose | Other Specs | Compar-ator | Country | Curr-ency | Price Year | HEALTH Outcome Type | Health Result - Difference | Costs - Difference *as REPORTED* | Incremental CE Ratio *as REPORTED* | CONVERTED ICER (using 2022 US$)† | Cost-Benefit Analysis | Dis-count Rate | Uncertainty & Sensitivity Analyses |
|---|---|---|---|---|---|---|---|---|---|---|---|---|---|---|---|---|
| | | | soluble) 0.005-0.020 [mg/kg or ppm?]; Iron NaFeEDTA 20 mg/kg or Ferrous fumarate 45 mg/kg | | | | | | | | | | | | | |
| Fiedler 2009 | Vitamin B12 + Folic acid + Iron | Wheat flour | Folic acid 1.5-4.0 mg/kg; Vitamin B12 (0.1% water soluble) 0.005-0.020 [mg/kg or ppm?]; Iron NaFeEDTA 20 mg/kg or Ferrous fumarate 45 mg/kg | Industrial | Pre-fortification | Philippines | US$ | 2009 | DALY saved | NR | 2.6 million | 23.20 | 28 | NR | 3% for DALYs; 0% for costs | NR |
| Fiedler 2009 | Vitamin B12 + Folic acid + Iron | Maize flour | Folic acid 1.5-4.0 mg/kg; Vitamin B12 (0.1% water soluble) 0.005-0.020 [mg/kg or ppm?]; Iron NaFeEDTA 20 mg/kg or Ferrous fumarate 45 mg/kg | Industrial | Pre-fortification | South Africa | US$ | 2009 | DALY saved | NR | 893.3 million | 15083 | 15,544 | NR | 3% for DALYs; 0% for costs | NR |
| Fiedler 2009 | Vitamin B12 + Folic acid + Iron | Wheat flour | Folic acid 1.5-4.0 mg/kg; Vitamin B12 (0.1% water soluble) 0.005-0.020 | Industrial | Pre-fortification | South Africa | US$ | 2009 | DALY saved | NR | 2.0 million | 49.60 | 51 | NR | 3% for DALYs; 0% for costs | NR |

| Study ID* (Author Year) | INTERVENTION-Micronutrient(s) | INTERVENTION-Food Vehicle(s) | Dose | Other Specs | Compar-ator | Country | Curr-ency | Price Year | HEALTH Outcome Type | Health Result - Difference | Costs - Difference *as REPORTED* | Incremental CE Ratio *as REPORTED* | CONVERTED ICER (using 2022 US$)† | Cost-Benefit Analysis | Dis-count Rate | Uncertainty & Sensitivity Analyses |
|---|---|---|---|---|---|---|---|---|---|---|---|---|---|---|---|---|
| | | | [mg/kg or ppm?]; Iron NaFeEDTA 20 mg/kg or Ferrous fumarate 45 mg/kg | | | | | | | | | | | | | |
| Fiedler 2009 | Vitamin B12 + Folic acid + Iron | Wheat flour | Folic acid 1.5-4.0 mg/kg; Vitamin B12 (0.1% water soluble) 0.005-0.020 [mg/kg or ppm?]; Iron NaFeEDTA 20 mg/kg or Ferrous fumarate 45 mg/kg | Industrial | Pre-fortification | Sudan | US$ | 2009 | DALY saved | NR | 2.9 million | 38.16 | 41 | NR | 3% for DALYs; 0% for costs | NR |
| Fiedler 2009 | Vitamin B12 + Folic acid + Iron | Maize flour | Folic acid 1.5-4.0 mg/kg; Vitamin B12 (0.1% water soluble) 0.005-0.020 [mg/kg or ppm?]; Iron NaFeEDTA 20 mg/kg or Ferrous fumarate 45 mg/kg | Industrial | Pre-fortification | Tanzania | US$ | 2009 | DALY saved | NR | 70.1 million | 273 | 347 | NR | 3% for DALYs; 0% for costs | NR |
| Fiedler 2009 | Vitamin B12 + Folic acid + Iron | Wheat flour | Folic acid 1.5-4.0 mg/kg; Vitamin B12 (0.1% water soluble) 0.005-0.020 [mg/kg or ppm?]; Iron | Industrial | Pre-fortification | Tanzania | US$ | 2009 | DALY saved | NR | 1.4 million | 10.85 | 14 | NR | 3% for DALYs; 0% for costs | NR |

| Study ID* (Author Year) | INTERVENTION-Micronutrient(s) | INTERVENTION-Food Vehicle(s) | Dose | Other Specs | Compar-ator | Country | Curr-ency | Price Year | HEALTH Outcome Type | Health Result - Difference | Costs - Difference *as REPORTED* | Incremental CE Ratio *as REPORTED* | CONVERTED ICER (using 2022 US$)† | Cost-Benefit Analysis | Dis-count Rate | Uncertainty & Sensitivity Analyses |
|---|---|---|---|---|---|---|---|---|---|---|---|---|---|---|---|---|
| | | | NaFeEDTA 20 mg/kg or Ferrous fumarate 45 mg/kg | | | | | | | | | | | | | |
| Fiedler 2009 | Vitamin B12 + Folic acid + Iron | Wheat flour | Folic acid 1.5-4.0 mg/kg; Vitamin B12 (0.1% water soluble) 0.005-0.020 [mg/kg or ppm?]; Iron NaFeEDTA 20 mg/kg or Ferrous fumarate 45 mg/kg | Industrial | Pre-fortification | Turkey | US$ | 2009 | DALY saved | NR | 41.4 million | 162.38 | 108 | NR | 3% for DALYs; 0% for costs | NR |
| Fiedler 2009 | Vitamin B12 + Folic acid + Iron | Maize flour | Folic acid 1.5-4.0 mg/kg; Vitamin B12 (0.1% water soluble) 0.005-0.020 [mg/kg or ppm?]; Iron NaFeEDTA 20 mg/kg or Ferrous fumarate 45 mg/kg | Industrial | Pre-fortification | Uganda | US$ | 2009 | DALY saved | NR | 36.3 million | 355 | 350 | NR | 3% for DALYs; 0% for costs | NR |
| Fiedler 2009 | Vitamin B12 + Folic acid + Iron | Wheat flour | Folic acid 1.5-4.0 mg/kg; Vitamin B12 (0.1% water soluble) 0.005-0.020 [mg/kg or ppm?]; Iron NaFeEDTA 20 mg/kg | Industrial | Pre-fortification | Uzbekistan | US$ | 2009 | DALY saved | NR | 3.0 million | 50.77 | 67 | NR | 3% for DALYs; 0% for costs | NR |

| Study ID* (Author Year) | INTERVENTION-Micronutrient(s) | INTERVENTION-Food Vehicle(s) | Dose | Other Specs | Compar-ator | Country | Curr-ency | Price Year | HEALTH Outcome Type | Health Result - Difference | Costs - Difference *as REPORTED* | Incremental CE Ratio *as REPORTED* | CONVERTED ICER (using 2022 US$)† | Cost-Benefit Analysis | Dis-count Rate | Uncertainty & Sensitivity Analyses |
|---|---|---|---|---|---|---|---|---|---|---|---|---|---|---|---|---|
| | | | or Ferrous fumarate 45 mg/kg | | | | | | | | | | | | | |
| Fiedler 2009 | Vitamin B12 + Folic acid + Iron | Wheat flour | Folic acid 1.5-4.0 mg/kg; Vitamin B12 (0.1% water soluble) 0.005-0.020 [mg/kg or ppm?]; Iron NaFeEDTA 20 mg/kg or Ferrous fumarate 45 mg/kg | Industrial | Pre-fortification | Vietnam | US$ | 2009 | DALY saved | NR | 2.0 million | 30.52 | 55 | NR | 3% for DALYs; 0% for costs | NR |
| Fiedler 2009 | Vitamin B12 + Folic acid + Iron | Wheat flour | Folic acid 1.5-4.0 mg/kg; Vitamin B12 (0.1% water soluble) 0.005-0.020 [mg/kg or ppm?]; Iron NaFeEDTA 20 mg/kg or Ferrous fumarate 45 mg/kg | Industrial | Pre-fortification | Yemen | US$ | 2009 | DALY saved | NR | 1.9 million | 32.04 | 51 | NR | 3% for DALYs; 0% for costs | NR |
| Fiedler 2009 | Vitamin B12 + Folic acid + Iron | Maize flour | Folic acid 1.5-4.0 mg/kg; Vitamin B12 (0.1% water soluble) 0.005-0.020 [mg/kg or ppm?]; Iron NaFeEDTA 20 mg/kg or Ferrous | Industrial | Pre-fortification | Zambia | US$ | 2009 | DALY saved | NR | 64.7 million | 3115 | 3,391 | NR | 3% for DALYs; 0% for costs | NR |

| Study ID* (Author Year) | INTERVENTION-Micronutrient(s) | INTERVENTION-Food Vehicle(s) | Dose | Other Specs | Compar-ator | Country | Curr-ency | Price Year | HEALTH Outcome Type | Health Result - Difference | Costs - Difference *as REPORTED* | Incremental CE Ratio *as REPORTED* | CONVERTED ICER (using 2022 US$)† | Cost-Benefit Analysis | Dis-count Rate | Uncertainty & Sensitivity Analyses |
|---|---|---|---|---|---|---|---|---|---|---|---|---|---|---|---|---|
| | | | fumarate 45 mg/kg | | | | | | | | | | | | | |
| Fiedler 2009 | Vitamin B12 + Folic acid + Iron | Wheat flour | Folic acid 1.5-4.0 mg/kg; Vitamin B12 (0.1% water soluble) 0.005-0.020 [mg/kg or ppm?]; Iron NaFeEDTA 20 mg/kg or Ferrous fumarate 45 mg/kg | Industrial | Pre-fortification | Zambia | US$ | 2009 | DALY saved | NR | 1.5 million | 52.34 | 57 | NR | 3% for DALYs; 0% for costs | NR |
| Fiedler 2009 | Vitamin B12 + Folic acid + Iron | Maize flour | Folic acid 1.5-4.0 mg/kg; Vitamin B12 (0.1% water soluble) 0.005-0.020 [mg/kg or ppm?]; Iron NaFeEDTA 20 mg/kg or Ferrous fumarate 45 mg/kg | Industrial | Pre-fortification | Zimbabwe | US$ | 2009 | DALY saved | NR | 75.8 million | 2232 | 2,612 | NR | 3% for DALYs; 0% for costs | NR |
| Fiedler 2009 | Vitamin B12 + Folic acid + Iron | Wheat flour | Folic acid 1.5-4.0 mg/kg; Vitamin B12 (0.1% water soluble) 0.005-0.020 [mg/kg or ppm?]; Iron NaFeEDTA 20 mg/kg or Ferrous fumarate 45 mg/kg | Industrial | Pre-fortification | Zimbabwe | US$ | 2009 | DALY saved | NR | 1.8 million | 42.54 | 50 | NR | 3% for DALYs; 0% for costs | NR |

| Study ID* (Author Year) | INTERVENTION-Micronutrient(s) | INTERVENTION-Food Vehicle(s) | Dose | Other Specs | Compar-ator | Country | Curr-ency | Price Year | HEALTH Outcome Type | Health Result - Difference | Costs - Difference *as REPORTED* | Incremental CE Ratio *as REPORTED* | CONVERTED ICER (using 2022 US$)† | Cost-Benefit Analysis | Dis-count Rate | Uncertainty & Sensitivity Analyses |
|---|---|---|---|---|---|---|---|---|---|---|---|---|---|---|---|---|
| Noshirvan 2021 | Vitamin B12 + Folic acid + Iron + Zinc | Wheat Flour | a) Iron (60 mg/kg, as ferrous fumarate) + b) zinc (95 mg/kg as zinc oxide) + c) vitamin B12 (0.04 mg/kg) + d) folic acid (3.75 mg/kg) | (folic acid dose is 75% of the WHO fortification target level) | Pre-fortification | Cameroon | US$ | 2013 | DALYs averted (for NTDs) | 52,750 (range, 51,880-53,530) | NR | 46.20 (range, 47.00-45.60) | 43 | NR | undiscounted | Cost/DALY of NTDs averted using 50% of target dose of folic acid instead is $49.50; while with 25% of folic acid dose it is $66.50. |
| Noshirvan 2021 | Vitamin B12 + Folic acid + Iron + Zinc | Wheat Flour | a) Iron (60 mg/kg, as ferrous fumarate) + b) zinc (95 mg/kg as zinc oxide) + c) vitamin B12 (0.04 mg/kg) + d) folic acid (5 mg/kg) | (folic acid dose is 100% of the WHO fortification target level) | Pre-fortification | Cameroon | US$ | 2013 | DALYs averted (for NTDs) | 55,050 (range, 54,140-55,860) | NR | 44.30 (range, 45.10-43.70) | 41 | NR | undiscounted | Cost/DALY of NTDs averted using 50% of target dose of folic acid instead is $49.50; while with 25% of folic acid dose it is $66.50. |
| Noshirvan 2021 | Vitamin B12 + Folic acid + Iron + Zinc | Wheat Flour | a) Iron (60 mg/kg, as ferrous fumarate) + b) zinc (95 mg/kg as zinc oxide) + c) vitamin B12 (0.04 mg/kg) + d) folic acid (2.5 mg/kg) | (This dose used of folic acid is 50% of the WHO fortification target level; as was seen in practice) | Pre-fortification | Cameroon | US$ | 2013 | DALYs averted (for NTDs and anemia) | 54,800 (range, 54,170-55,360) | 1,989,000 | 36.30 (range, 36.70-35.90) | 34 | NR | 3% for both costs and benefits | Varying discount rate to 6% gives estimate of $38.30/DALY averted; while undiscounted ICER is $34.50/DALY averted. |
| Niedermaier 2021 | Vitamin D | NR [data from several food types combined] | 400 IU/day | NR | No fortification | Germany | € | 2016 | Cancer deaths prevented | 25,281 | -996,073,000 | Dominant | Dominant | NR | NR | Sensitivity analysis of higher or lower costs of fortification (by 20%) had only |

| Study ID* (Author Year) | INTERVENTION-Micronutrient(s) | INTERVENTION-Food Vehicle(s) | Dose | Other Specs | Compar-ator | Country | Curr-ency | Price Year | HEALTH Outcome Type | Health Result - Difference | Costs - Difference *as REPORTED* | Incremental CE Ratio *as REPORTED* | CONVERTED ICER (using 2022 US$)† | Cost-Benefit Analysis | Dis-count Rate | Uncertainty & Sensitivity Analyses |
|---|---|---|---|---|---|---|---|---|---|---|---|---|---|---|---|---|
| | | | | | | | | | | | | | | | | minor impact on net savings. |
| Niedermaier 2021 | Vitamin D | NR [data from several food types combined] | 600 IU/day | NR | No fortification | Germany | € | 2016 | Cancer deaths prevented | 29,877 | -1,177,607,000 | Dominant | Dominant | NR | NR | Sensitivity analysis of higher or lower costs of fortification (by 20%) had only minor impact on net savings. |
| Niedermaier 2021 | Vitamin D | NR [data from several food types combined] | 800 IU/day | NR | No fortification | Germany | € | 2016 | Cancer deaths prevented | 29,877 | -1,359,143,000 | Dominant | Dominant | NR | NR | Sensitivity analysis of higher or lower costs of fortification (by 20%) had only minor impact on net savings. |
| Niedermaier 2021 | Vitamin D | NR [data from several food types combined] | 1000 IU/day | NR | No fortification | Germany | € | 2016 | Cancer deaths prevented | 35,164 | -1,384,395,000 | Dominant | Dominant | NR | NR | Sensitivity analysis of higher or lower costs of fortification (by 20%) had only minor impact on net savings. |
| Sandmann 2017 | Vitamin D + Calcium | Bread | Providing daily additional intake of 20 mcg (800 IU) of cholecalciferol and 200 mg of calcium | NR | No fortification | Germany | Euros (€) | 2014 | Fractures prevented | 36,705 per year | -314,800,000 per year | Dominant | Dominant | BCR 9:1 | NR [costs were adjusted for inflation] | Varying the risks of fractures led to 110% increase in net savings or 143% decrease in net savings; and modifying fracture incidence rates resulted in annual net savings changes of −34% and 34%. Analyses were largely insensitive to variations in production cost parameters and changes in fortification levels. |
| Palacios 2022 | Calcium | Flour | 156 mg per 100 g of staple food | NR | Unfortified flour | Costa Rica | US$ | NR [2019 meeting] | Achieving recommended calcium intake | NR | 212,976 | $0.043 per covered individual per year | 0.04 | NR | No discount | NR |
| Dalziel 2010 | Folic acid | Wheat flour for bread-making (High cost scenario) | 200 mcg per 100 g bread | NR | Voluntary fortification | Australia | Australian dollar (AU$) | 2006 | DALYs averted | 1970 | 11,351,261 per year | ICER AU$ 133,100/DALY averted | 143,241 | NR | 5% for both costs and | Sensitivity analysis revealed high sensitivity to selected assumptions, |

| Study ID* (Author Year) | INTERVENTION-Micronutrient(s) | INTERVENTION-Food Vehicle(s) | Dose | Other Specs | Compar-ator | Country | Curr-ency | Price Year | HEALTH Outcome Type | Health Result - Difference | Costs - Difference *as REPORTED* | Incremental CE Ratio *as REPORTED* | CONVERTED ICER (using 2022 US$)† | Cost-Benefit Analysis | Dis-count Rate | Uncertainty & Sensitivity Analyses |
|---|---|---|---|---|---|---|---|---|---|---|---|---|---|---|---|---|
| | | | | | | | | | | | | | | | benefits | particularly focusing on the cost of the intervention and the 95% CI for translating changes in mean folic acid intake into NTD. S.A. range for ICERs: AU$ 95,700 to 296,600/DALY |
| Dalziel 2010 | Folic acid | Wheat flour for bread-making (Low cost scenario) | 200 mcg per 100 g bread | NR | Voluntary fortification | Australia | Australian dollar (AU$) | 2006 | DALYs averted | 1970 | 1,552,408 per year | ICER AU$ 13,700/DALY averted | 14,744 | NR | 5% for both costs and benefits | Sensitivity analysis revealed high sensitivity to selected assumptions, particularly focusing on the cost of the intervention and the 95% CI for translating changes in mean folic acid intake into NTD. S.A. range for ICERs: AU$ 9,500 to 177,200/DALY |
| Access Economics 2006 | Folic acid | All bread making flour | 100 mcg folic acid per 100 g flour | NR | Voluntary fortification | Australia | AUD $ | 2005 | DALYs prevented | 289.9 (95% CI, 156.1-546.4) | 2,486,400 upfront; 1,152,357 ongoing per year | NR | N/A | BCR: 41.9 | 3.3% | Sensitivity analysis: conclusions are also consistent if the benefits associated with preventing terminations and still births are excluded. |
| Access Economics 2006 | Folic acid | All bread making flour | 200 mcg folic acid per 100 g flour | NR | Voluntary fortification | Australia | AUD $ | 2005 | DALYs prevented | 724.8 (95% CI, 390.3-1365.9) | 2,486,400 upfront; 1,208,357 ongoing per year | NR | N/A | BCR: 100.6 | 3.3% | Sensitivity analysis: conclusions are also consistent if the benefits associated with preventing terminations and still births are excluded. |
| Rabovskaja 2013 | Folic acid | Bread-making flour | NR | NR | Voluntary fortification | Australia | Australian dollars (A$) | 2005 | QALYs gained | 503 per year | 5,780,423 | ICER A$ 11,485 per QALY gained | 12,992 | NR | 5% for costs and benefits | Results appeared largely insensitive to single parameter variation. Scenario analyses of impact of extended survival |

| Study ID* (Author Year) | INTERVENTION-Micronutrient(s) | INTERVENTION-Food Vehicle(s) | Dose | Other Specs | Compar-ator | Country | Curr-ency | Price Year | HEALTH Outcome Type | Health Result - Difference | Costs - Difference *as REPORTED* | Incremental CE Ratio *as REPORTED* | CONVERTED ICER (using 2022 US$)† | Cost-Benefit Analysis | Dis-count Rate | Uncertainty & Sensitivity Analyses |
|---|---|---|---|---|---|---|---|---|---|---|---|---|---|---|---|---|
| | | | | | | | | | | | | | | | | of patients, upfront costs, discounting by 0% and 7%, and inclusion of loss of consumer choice on an ongoing basis did not vary considerably except for inclusion of loss of the consumer choice; with addition of loss of consumer choice, ICER increased to ~AU$ 800,000/QALY. |
| Saing 2019/ Australian HMAC 2017 | Folic acid | Bread-making flour | 134–200 mcg/L | NR | Voluntary fortification | Australia | Australian $ | 2014 | QALYs gained | 293 per year | -1,450,326 | Dominant | Dominant | NR | 5% for both costs and benefits | Sensitivity analyses varying model parameters: the model was most sensitive to estimation of post-fortification rate of NTDs by age group. |
| Rodrigues 2023 | Folic acid | Flour | NR | NR | Pre-fortification | Brazil | R$ | 2019 | Neural tube defects prevented | 7,367 among all births (3,499 among live births) | -21,106,715 (cost-of-illness) | Dominant | Dominant | NR | NR | NR |
| Vosti 2023 | Folic acid | Bouillon cubes | 100 mg/kg | NR | No fortification | Cameroon | US$ | 2019 | Child lives saved | 9605 | Total program costs US $ 3,314,000 | 345 | 354 | NR | NR | The program selection of “high” or “low” effectiveness results did not influence the key policy messages. |
| Vosti 2023 | Folic acid | Wheat flour | 1.65 mg/kg | NR | No fortification | Cameroon | US$ | 2019 | Child lives saved | 4373 | Total program costs US $ 1,551,000 | 354 | 363 | NR | NR | The program selection of “high” or “low” effectiveness results did not influence the key policy messages. |
| Vosti 2023 | Folic acid | Wheat flour | 5 mg/kg | NR | No fortification | Cameroon | US$ | 2019 | Child lives saved | 6878 | Total program costs | 681 | 698 | NR | NR | The program selection of “high” or “low” |

| Study ID* (Author Year) | INTERVENTION-Micronutrient(s) | INTERVENTION-Food Vehicle(s) | Dose | Other Specs | Compar-ator | Country | Curr-ency | Price Year | HEALTH Outcome Type | Health Result - Difference | Costs - Difference *as REPORTED* | Incremental CE Ratio *as REPORTED* | CONVERTED ICER (using 2022 US$)† | Cost-Benefit Analysis | Dis-count Rate | Uncertainty & Sensitivity Analyses |
|---|---|---|---|---|---|---|---|---|---|---|---|---|---|---|---|---|
| | | | | | | | | | | | US $4,684,000 | | | | | effectiveness results did not influence the key policy messages. |
| Adams 2022/ Vosti 2023 | Folic acid | Wheat flour + Bouillon cube | wheat flour (5 mg/kg) + bouillon cube (100 mg/kg) | NR | No fortification /interventio n | Cameroon | US$ | 2019 | Effective coverage: those at risk of deficiency (inadequate intake) who then received sufficient additional intake from intervention | 32,293 | 17,718 | $ 0.55 per WRA-year effectively covered | 0.56 | NR | NR | NR |
| Llanos 2007 | Folic acid | Wheat flour | 2.2 mg per kg flour | NR | Pre-fortification | Chile | Interna tional Dollars (I$) | 2001 | DALYs averted per year | 2,300 (range, 2,000-2,500) | -2,300,000 | ICER I$ 89/DALY averted (range, 101–79) | 13 | BCR: 11.8 | 3% for health; 0% for costs | Sensitivity analysis without discounting benefits resulted in a lower implementation cost of I$42/DALY averted. Taking into account averted costs of care, fortification was dominant (net savings). |
| Jentink 2008 | Folic acid | Flour | 140 mcg per 100 g flour | NR | No fortification | Netherlands | Euro | 2005 | QALYs gained | 7 NTDs prevented | NR | Dominant | Dominant | NR | 4% for costs; 1.5% for health gains | Was cost-saving if 6 or more NTDs are averted annually. Remains cost-effective as long as the enrichment costs do not exceed Euros 5.5 million (threshold at Euros 20,000/QALY). |
| Dalziel 2010 | Folic acid | Wheat flour for bread-making | 135 mcg per 100 g bread | NR | Voluntary fortification | New Zealand | New Zealan d dolllars (NZ$) | 2006 | DALYs averted | 574 | 2,966,437 per year | ICER NZ$ 138,500/DALY averted | 156,357 | NR | 5% for both costs and benefit s | Sensitivity analysis revealed high sensitivity to selected assumptions, particularly focusing on the cost of the intervention and the |

| Study ID* (Author Year) | INTERVENTION-Micronutrient(s) | INTERVENTION-Food Vehicle(s) | Dose | Other Specs | Compar-ator | Country | Curr-ency | Price Year | HEALTH Outcome Type | Health Result - Difference | Costs - Difference *as REPORTED* | Incremental CE Ratio *as REPORTED* | CONVERTED ICER (using 2022 US$)† | Cost-Benefit Analysis | Dis-count Rate | Uncertainty & Sensitivity Analyses |
|---|---|---|---|---|---|---|---|---|---|---|---|---|---|---|---|---|
| | | | | | | | | | | | | | | | | 95% CI for translating changes in mean folic acid intake into NTD. S.A. range for ICERs: AU$ 79,700 to 251,300/DALY |
| Access Economics 2006 | Folic acid | All bread making flour | 100 mcg folic acid per 100 g flour | NR | Voluntary fortification | New Zealand | NZD $ | 2005 | DALYs prevented | 108.1 (95% CI, 54.1-198.3) | 1,690,000 upfront; 2,336,910 ongoing per year | NR | N/A | BCR: 9.5 | 3.8% | Sensitivity analysis: if the benefits associated with preventing terminations and still births are excluded, based on the lower estimate of NTDs prevented, the costs outweigh the benefits. |
| Access Economics 2006 | Folic acid | All bread making flour | 200 mcg folic acid per 100 g flour | NR | Voluntary fortification | New Zealand | NZD $ | 2005 | DALYs prevented | 218.0 (95% CI, 109.0-399.6) | 1,690,000 upfront; 2,348,658 ongoing per year | NR | N/A | BCR: 19.4 | 3.8% | Sensitivity analysis: conclusions are also consistent if the benefits associated with preventing terminations and still births are excluded. |
| Sayed 2008 | Folic acid | Maize meal and wheat flour | Wheat flour 1.5 mg/kg and Maize meal 2.21 mg/kg | NR | Pre-fortification | South Africa | South African Rand (ZAR) | 2006 | Decline in perinatal mortality rates for NTDs | 65.9% | NR | NR | N/A | BCR 30:1 | NR | If the true national prevalence rate for NTDs is greater than that observed in the hospital based surveillance system, the BCR will be greater. |
| Bentley 2009 | Folic acid | Grain products | 140 mcg/100 g | NR | Pre-fortification | USA | US$ | 2005 | QALYs gained annually | 56,291 | -780,500,000 annually | Dominant | Dominant | NR | 3% for both costs and QALYs | Sensitivity analyses of a range of estimates of the model's assumptions for QALY, costs, disease risks, and discount rates showed that, even with extreme estimates, the conclusions remained consistent. |

| Study ID* (Author Year) | INTERVENTION- Micronutrient(s) | INTERVENTION- Food Vehicle(s) | Dose | Other Specs | Compar- ator | Country | Curr- ency | Price Year | HEALTH Outcome Type | Health Result - Difference | Costs - Difference *as REPORTED* | Incremental CE Ratio *as REPORTED* | CONVERTED ICER (using 2022 US$)[†] | Cost- Benefit Analysis | Dis- count Rate | Uncertainty & Sensitivity Analyses |
|---|---|---|---|---|---|---|---|---|---|---|---|---|---|---|---|---|
| Bentley 2009 | Folic acid | Grain products | 350 mcg/100 g | NR | Pre- fortification | USA | US$ | 2005 | QALYs gained annually | 185,165 | - 2,528,800,000 annually | Dominant | Dominant | NR | 3% for both costs and QALYs | Sensitivity analyses of a range of estimates of the model's assumptions for QALY, costs, disease risks, and discount rates showed that, even with extreme estimates, the conclusions remained consistent. |
| Bentley 2009 | Folic acid | Grain products | 700 mcg/100 g | NR | Pre- fortification | USA | US$ | 2005 | QALYs gained annually | 322,940 | - 4,364,800,000 annually | Dominant | Dominant | NR | 3% for both costs and QALYs | Sensitivity analyses of a range of estimates of the model's assumptions for QALY, costs, disease risks, and discount rates showed that, even with extreme estimates, the conclusions remained consistent. |
| Grosse 2016 | Folic acid | Cereal grain products | 140 mcg per 100 g | Mills-based fortification | Pre- mandatory fortification | USA | US$ | 2014 | Spina bifida cases (live births) averted | 614 annually | -299,000 (excluding caregiver savings; "worst case") | Dominant | Dominant | NR | NR | Anaylses with increased fortification cost (or also spina bifida averted or including caregiver savings) resulted in consistent conclusions (i.e., dominant ICER) with greater CE. |
| Hoddinott 2018 | Folic acid | Maize flour | 2.1 mg folic acid per 2.21 kg maize flour | NR | Pre- fortification | Zambia | US$ | 2017 | DALYs averted | 172,860 | $2,576,000 | $ 14.90/DALY averted | 15 | NR | No discounting | Sensitivity analyses found these ICERs, as cost per DALY averted: High effectiveness $9.93; Low effectiveness $29.80; High cost $23.43; Low cost $11.22; High cost + |

| Study ID* (Author Year) | INTERVENTION-Micronutrient(s) | INTERVENTION-Food Vehicle(s) | Dose | Other Specs | Compar-ator | Country | Curr-ency | Price Year | HEALTH Outcome Type | Health Result - Difference | Costs - Difference *as REPORTED* | Incremental CE Ratio *as REPORTED* | CONVERTED ICER (using 2022 US$)† | Cost-Benefit Analysis | Dis-count Rate | Uncertainty & Sensitivity Analyses |
|---|---|---|---|---|---|---|---|---|---|---|---|---|---|---|---|---|
| | | | | | | | | | | | | | | | | Low effectiveness $46.86 |
| GAIN 2017 | Folic acid + Iron | Wheat flour | NR | NR | No fortification | Afghanistan | US$ | 2017 | Benefit-cost ratio | NR | 53,340,000 | NR | N/A | BCR: 12.46 | 1.5% | NR |
| Kancherla 2021 | Folic acid + Iron | Wheat flour | Iron 40 ppm NaFeEDTA, 60 ppm ferrous fumarate/sulfate; Folic acid 2.6 ppm | NR | No fortification | Angola | US$ | NR | DALYs averted | DALYs Averted = 57,522 | NR | NR | N/A | BCR: 38.27 | 3% | Sensitivity analysis varying discount rate to 5% found conclusions remained consistent. |
| Kancherla 2021 | Folic acid + Iron | Wheat flour and Rice | Wheat Flour: Iron 40 ppm NaFeEDTA, 60 ppm ferrous fumarate/sulfate; Folic acid 5 ppm. Rice: Iron 70 ppm as micronized ferric pyrophosphate; Folic acid 1 ppm | NR | No fortification | Bangladesh | US$ | NR | DALYs averted | DALYs Averted = 369,392 | NR | NR | N/A | BCR: 3.7 | 3% | Sensitivity analysis varying discount rate to 5% found conclusions remained consistent. |
| Kancherla 2021 | Folic acid + Iron | Rice | Iron 120 ppm as micronized ferric pyrophosphate; folic acid 2.6 ppm | NR | No fortification | Benin | US$ | NR | DALYs averted | DALYs Averted = 38,998 | NR | NR | N/A | BCR: 5.58 | 3% | Sensitivity analysis varying discount rate to 5% found conclusions remained consistent. |
| Asian Develop. Bank 2004 | Folic acid + Iron | Wheat flour | 60 parts per million (ppm) iron and 2 ppm for folic acid | Roller flour mills. NaFeEDTA iron compound | Unfortified food | China | US$ | NR | Deaths averted | 32,000 | 543,346,000 | NR | N/A | BCR: 3.0 | NR [Discounted costs and benefits] | NR |

| Study ID* (Author Year) | INTERVENTION-Micronutrient(s) | INTERVENTION-Food Vehicle(s) | Dose | Other Specs | Compar-ator | Country | Curr-ency | Price Year | HEALTH Outcome Type | Health Result - Difference | Costs - Difference *as REPORTED* | Incremental CE Ratio *as REPORTED* | CONVERTED ICER (using 2022 US$)† | Cost-Benefit Analysis | Dis-count Rate | Uncertainty & Sensitivity Analyses |
|---|---|---|---|---|---|---|---|---|---|---|---|---|---|---|---|---|
| Kancherla 2021 | Folic acid + Iron | Wheat flour and Rice | Wheat Flour: Iron 20 ppm NaFeEDTA, 30 ppm ferrous fumarate/sulfate, 60 ppm as electrolytic iron; Folic acid 1.3 ppm. Rice: iron 70 ppm as micronized ferric pyrophosphate; Folic acid 1.3 ppm | NR | No fortification | China | US$ | NR | DALYs averted | DALYs Averted = 1,701,396 | NR | NR | N/A | BCR: 14.23 | 3% | Sensitivity analysis varying discount rate to 5% found conclusions remained consistent. |
| Kancherla 2021 | Folic acid + Iron | Rice | Iron 70 ppm as micronized ferric pyrophosphate; folic acid 1.3 ppm | NR | No fortification | Côte d'Ivoire | US$ | NR | DALYs averted | DALYs Averted = 26,294 | NR | NR | N/A | BCR: 6.87 | 3% | Sensitivity analysis varying discount rate to 5% found conclusions remained consistent. |
| Kancherla 2021 | Folic acid + Iron | Wheat flour | Iron 15 ppm NaFeEDTA, 20 ppm ferrous fumarate/sulfate, 40 ppm as electrolytic iron; Folic acid 1 ppm | NR | No fortification | Egypt | US$ | NR | DALYs averted | DALYs Averted = 161,352 | NR | NR | N/A | BCR: 7.11 | 3% | Sensitivity analysis varying discount rate to 5% found conclusions remained consistent. |
| Kancherla 2021 | Folic acid + Iron | Wheat flour | Iron 40 ppm NaFeEDTA, 60 ppm ferrous fumarate/sulfate; Folic | NR | Voluntary fortification | Ethiopia | US$ | NR | DALYs averted | DALYs Averted = 140,088 | NR | NR | N/A | BCR: 1.5 | 3% | Sensitivity analysis varying discount rate to 5% found conclusions remained consistent. |

| Study ID* (Author Year) | INTERVENTION- Micronutrient(s) | INTERVENTION- Food Vehicle(s) | Dose | Other Specs | Compar- ator | Country | Curr- ency | Price Year | HEALTH Outcome Type | Health Result - Difference | Costs - Difference *as REPORTED* | Incremental CE Ratio *as REPORTED* | CONVERTED ICER (using 2022 US$)† | Cost- Benefit Analysis | Dis- count Rate | Uncertainty & Sensitivity Analyses |
|---|---|---|---|---|---|---|---|---|---|---|---|---|---|---|---|---|
| | | | acid 2.6 ppm | | | | | | | | | | | | | |
| Kancherla 2021 | Folic acid + Iron | Rice | Iron 120 ppm as micronized ferric pyrophosphate; folic acid 2.6 ppm | NR | No fortification | Ghana | US$ | NR | DALYs averted | DALYs Averted = 54,008 | NR | NR | N/A | BCR: 6.59 | 3% | Sensitivity analysis varying discount rate to 5% found conclusions remained consistent. |
| Fiedler 2012 | Folic acid + Iron | Wheat flour (in Gujarat's Public Distribution System) | Iron (30 ppm) 2.4-3.0 mg/day; Folic acid dose NR | NR | Unfortified food | India | US$ | Unclear | DALYs saved | 39,821 | 355,860 annually | Cost per DALY averted US$ 8.9 | 10 | NR | NR | Estimate for the 3 programs combined is $ 27.6 /DALY saved but this considers each of the SSNPs independently, not considering that some individuals may benefit from more than one of the programs. |
| Kancherla 2021 | Folic acid + Iron | Wheat flour and Rice | Wheat Flour: Iron 20 ppm NaFeEDTA; Folic acid 1.3 ppm. Rice: Iron 70 ppm as micronized ferric pyrophosphate; Folic acid 1.3 ppm | NR | Voluntary fortification | India (17 states) | US$ | NR | DALYs Averted; Economic Value of DALYs Averted (in USD $ m) - (Cases of anemia in WRA, cases of neural tube defects, child deaths) | DALYs Averted = 2,198,103 | NR | NR | N/A | BCR: 9.01 | 3% | Sensitivity analysis varying discount rate to 5% found conclusions remained consistent. |
| Kancherla 2021 | Folic acid + Iron | Rice | Iron 70 ppm as micronized ferric pyrophosphate; folic acid 1.3 ppm | NR | No fortification | Liberia | US$ | NR | DALYs averted | DALYs Averted = 8,555 | NR | NR | N/A | BCR: 1.82 | 3% | Sensitivity analysis varying discount rate to 5% found conclusions remained consistent. |

| Study ID* (Author Year) | INTERVENTION-Micronutrient(s) | INTERVENTION-Food Vehicle(s) | Dose | Other Specs | Compar-ator | Country | Curr-ency | Price Year | HEALTH Outcome Type | Health Result - Difference | Costs - Difference *as REPORTED* | Incremental CE Ratio *as REPORTED* | CONVERTED ICER (using 2022 US$)† | Cost-Benefit Analysis | Dis-count Rate | Uncertainty & Sensitivity Analyses |
|---|---|---|---|---|---|---|---|---|---|---|---|---|---|---|---|---|
| Kancherla 2021 | Folic acid + Iron | Rice | Iron 120 ppm as micronized ferric pyrophosphate; folic acid 2.6 ppm | NR | No fortification | Nigeria | US$ | NR | DALYs averted | DALYs Averted = 362,564 | NR | NR | N/A | BCR: 19.47 | 3% | Sensitivity analysis varying discount rate to 5% found conclusions remained consistent. |
| MQSUN 2014 | Folic acid + Iron | Wheat flour | NR | Compound: Iron (NaFeEDTA), folic acid (NR); fortification methods (millers/ small-scale Chakki millers) | No fortification /Current coverage=0 | Pakistan | US Dollars (presumably) | NR | Maternal lives saved | 491 | 36,000,000 | $73,423 per Maternal life saved | 62,128 | BCR 7.2:1 (for maternal and children combined) | 3% | NR |
| MQSUN 2014 | Folic acid + Iron | Wheat flour | NR | Compound: Iron (NaFeEDTA), folic acid (NR); fortification methods (millers/ small-scale Chakki millers) | No fortification /Current coverage=0 | Pakistan | US Dollars (presumably) | NR | Child lives saved | 3,285 | 36,000,000 | $10,973 per Child life saved | 9,285 | (see above) | 3% | NR |

| Study ID* (Author Year) | INTERVENTION- Micronutrient(s) | INTERVENTION- Food Vehicle(s) | Dose | Other Specs | Compar- ator | Country | Curr- ency | Price Year | HEALTH Outcome Type | Health Result - Difference | Costs - Difference *as REPORTED* | Incremental CE Ratio *as REPORTED* | CONVERTED ICER (using 2022 US$)† | Cost- Benefit Analysis | Dis- count Rate | Uncertainty & Sensitivity Analyses |
|---|---|---|---|---|---|---|---|---|---|---|---|---|---|---|---|---|
| Asian Develop. Bank 2004 | Folic acid + Iron | Wheat flour | 60 parts per million (ppm) iron and 1.5 ppm folic acid | Roller flour mills | Unfortified food | Pakistan | US$ | NR | Deaths averted | 38,000 | 337,862,000 | NR | N/A | BCR: 2.0 | NR [Discounted costs and benefits] | NR |
| Ghauri 2015 | Folic acid + Iron | Wheat flour (Atta) | NR | NR | No fortification | Pakistan | US$ | 2015 | Benefit-cost ratio | NR | 442,000,000 | NR | N/A | BCR: 5.34 | 4% | If premix is exempted from taxes and duties, overall cost will reduce to $ 0.270 billion leading to 8.75 times more benefit than its cost. |
| Kancherla 2021 | Folic acid + Iron | Rice | Iron 70 ppm as micronized ferric pyrophosphate; folic acid 1.3 ppm | NR | No fortification | Senegal | US$ | NR | DALYs averted | DALYs Averted = 32,183 | NR | NR | N/A | BCR: 4.06 | 3% | Sensitivity analysis varying discount rate to 5% found conclusions remained consistent. |
| Ghauri 2016 | Folic acid + Iron | Wheat flour | NR | NR | No fortification | Tajikistan | US$ | 2016 | Benefit-cost ratio | NR | 32,058,000 | NR | N/A | BCR: 9.44 | 2.5% | NR |
| Kancherla 2021 | Folic acid + Iron | Wheat flour | Iron 15 ppm NaFeEDTA, 20 ppm ferrous fumarate/sulfate, 40 ppm as electrolytic iron; Folic acid 1 ppm | NR | Voluntary fortification | Tajikistan | US$ | NR | DALYs averted | DALYs Averted = 11,371 | NR | NR | N/A | BCR: 3.3 | 3% | Sensitivity analysis varying discount rate to 5% found conclusions remained consistent. |
| Asian Develop. Bank 2004 | Folic acid + Iron | Wheat flour | 60 parts per million (ppm) iron and 2 ppm for folic acid | Roller flour mills | Unfortified food | Thailand | US$ | NR | Deaths averted | 5,000 | 13,400,000 | NR | N/A | BCR: 4.9 | NR [Discounted costs and benefits] | NR |
| Asian Develop. Bank 2004 | Folic acid + Iron | Wheat flour | 60 parts per million (ppm) iron and 2 ppm | Roller flour mills. NaFeEDTA | Unfortified food | Viet Nam | US$ | NR | Deaths averted | 8,000 | 37,125,000 | NR | N/A | BCR: 9.7 | NR [Discounted costs and | NR |

| Study ID* (Author Year) | INTERVENTION-Micronutrient(s) | INTERVENTION-Food Vehicle(s) | Dose | Other Specs | Compar-ator | Country | Curr-ency | Price Year | HEALTH Outcome Type | Health Result - Difference | Costs - Difference *as REPORTED* | Incremental CE Ratio *as REPORTED* | CONVERTED ICER (using 2022 US$)† | Cost-Benefit Analysis | Dis-count Rate | Uncertainty & Sensitivity Analyses |
|---|---|---|---|---|---|---|---|---|---|---|---|---|---|---|---|---|
| | | | for folic acid | iron compo und | | | | | | | | | | | benefit s] | |
| Australian HMAC 2017 | Iodine | Salt in bread | 25-65 mg per kg of salt | NR | Voluntary fortification | Australia | AUD $ | 2014 | Achieving threshold UIC level (100 µg/L non-pregnant or 150 µg/L pregnant) | 0.30 | 10,576,624 | $AUD 347,604 per percentage point increase in proportion of population with UIC above threshold | 293,466 | NR | 5% | NR |
| Huang 2020 | Iodine | Salt | NR | NR | Pre-fortification | China | RMB | 1995 | Children attending primary school every year (in high-goiter counties) | 39,040 to 58,560 | 153,800,000 | NR | N/A | Cost of saving (i.e., now attending school) an out-of-primary-school child = 2,626 to 3,940 RMB [which equals 1.66-2.50 times of rural citizens' per capita annual income in 1995]. (And additional schooling would translate into a lifetime wage gain of 55,370 RMB). (BCR NR). | NR | NR |
| Rajkumar 2012 | Iodine | Salt | * | NR | Unfortified | Ethiopia | US$ | 2007 | Deaths averted | NR | Cost per beneficiary $0.05 | $ 170.64 per death averted | 291 | BCR: 81.00 | 5% | NR |

| Study ID* (Author Year) | INTERVENTION- Micronutrient(s) | INTERVENTION- Food Vehicle(s) | Dose | Other Specs | Compar- ator | Country | Curr- ency | Price Year | HEALTH Outcome Type | Health Result - Difference | Costs - Difference *as REPORTED* | Incremental CE Ratio *as REPORTED* | CONVERTED ICER (using 2022 US$)† | Cost- Benefit Analysis | Dis- count Rate | Uncertainty & Sensitivity Analyses |
|---|---|---|---|---|---|---|---|---|---|---|---|---|---|---|---|---|
| Rochau 2019 | Iodine | Salt | NR | NR | No fortification program/prevention | Germany | Euro (€) | 2018 | QALYs gained | 0.04 per person (or more) | -522 per person (or more savings) | Dominant | Dominant | NR | 3% for costs and QALYs | For all 3 scenarios, and in most sensitivity analyses, the prevention program remained dominant. |
| Pandav 2012 | Iodine | Salt | NR | Potassium iodate | No fortification /preventive program | India (Sikkim state) | Indian rupee (₹) | 2010 | Visible goiter person-years | -772,532 | 59,225,964 (total) | NR | N/A | BCR: 1.61 | 10% | NR |
| Australian HMAC 2017 | Iodine | Salt in bread | 25-65 mg per kg of salt | NR | Voluntary fortification | New Zealand | NZD $ | 2014 | Achieving threshold UIC level (100 µg/L non-pregnant or 150 µg/L pregnant) | 0.32 | 1,622,354 | $NZD 50,293 per percentage point increase in proportion of population with UIC above threshold | 40,535 | NR | 5% | Only small changes in productivity or health-related quality of life are required before the benefits exceed the costs of mandatory iodine fortification. |
| Horton 2008 | Iodine | Salt | NR | NR | Unfortified salt | South Asia, sub-Saharan Africa, CEE/CIS | US$ | 2000 | Benefit-cost ratio | NR | $19,000,000 per year | NR | N/A | BCR 30:1 | 3% | Sensitivity analysis increasing discount rate to 6% gives BCR of 12:1. Using DALY value of $5000 gave consistent results to $1000/DALY. |
| Gorstein 2020 | Iodine | Salt | NR | Universal Salt Iodization Programs | Pre-fortification | Worldwide (>100 countries) | US$ | 2020 | Goiter prevalence | -9.9% | NR (benefits of nearly $33 billion saved from IDD losses) | Dominant | Dominant | NR | 3% | NR |
| Baltussen 2004 | Iron | Cereal flours | 80% geographical coverage; dose NR | electrolytic iron (<45 microm, 325 mesh) | no iron fortification or supplementation program | African subregion AfrD (Algeria, Angola, Benin, Burkina Faso, Cameroon, Cape Verde, Chad, Comoros, Equatorial Guinea, Gabon, Gambia, | international dollars | 2000 | DALY averted | 913,414 | I$ 19,192,967 | ICER I$ 21/DALY averted | 91 | NR | 3% for both costs and health effects | Uncertainty analysis focusing on the impact of varying assumptions about epidemiology, costs, and intervention effectiveness revealed that the results were quite robust to variation in the model parameters within +/-25%. Varying geographical coverage from 80% |

| Study ID* (Author Year) | INTERVENTION-Micronutrient(s) | INTERVENTION-Food Vehicle(s) | Dose | Other Specs | Compar-ator | Country | Curr-ency | Price Year | HEALTH Outcome Type | Health Result - Difference | Costs - Difference *as REPORTED* | Incremental CE Ratio *as REPORTED* | CONVERTED ICER (using 2022 US$)† | Cost-Benefit Analysis | Dis-count Rate | Uncertainty & Sensitivity Analyses |
|---|---|---|---|---|---|---|---|---|---|---|---|---|---|---|---|---|
| | | | | | | Ghana, Guinea, Guinea-Bissau, Liberia, Madagascar, Mali, Mauritania, Mauritius, Niger, Nigeria, Sao Tome And Principe, Senegal, Seychelles, Sierra Leone, Togo) | | | | | | | | | | to: 50% or 95% gave ICERs of I$ 27 or 20 per DALY averted (resp'ly). |
| Wei 2023 | Iron | Soy sauce | Dose NR. Media promotion. | NR | Pre-Iron-fortified soy sauce promotion intervention | China | RMB (yuan) | 2013 | Anemia prevalence | -9.2% | major cost of the project 156,400 RMB; and total benefits from projects 1,484,485 RMB | Dominant | Dominant | BCR: 9.49 | 3% | When per capita annual disposable income is adjusted (range 10,000 – 35,000 yuan) and discount rate is adjusted (range0.5 – 4.5%), iron-fortified soy sauce always dominates pre-intervention measures (ratio range 1 : 4.32 to 1 : 21.16). |
| Ma 2008 | Iron | Wheat flour and Soy sauce | NR | NR | No fortification | China | international dollars ($Int) | 2000 | DALYs saved | NR | 0.06 per capita per year | ICER I$ 66/DALY saved | 166 | NR | NR | NR |
| Asian Develop. Bank 2004 | Iron | Soy sauce | 60 parts per million (ppm) iron | NaFeEDTA | Unfortified food | China | US$ | NR | Deaths averted | 500 | 2,250,238,000 | NR | N/A | BCR: 9.4 | NR [Discounted costs and benefits] | NR |
| Baltussen 2004 | Iron | Cereal flours | 80% geographical coverage; dose NR | electrolytic iron (<45 micro | no iron fortification or supplement | European subregion EURA (Andorra, Austria, | international dollars | 2000 | DALY averted | 7,452 | I$ 41,649,470 | ICER I$ 5,589/DALY averted | 25,232 | NR | 3% for both costs and | Uncertainty analysis focusing on the impact of varying assumptions about epidemiology, costs, |

| Study ID* (Author Year) | INTERVENTION-Micronutrient(s) | INTERVENTION-Food Vehicle(s) | Dose | Other Specs | Compar-ator | Country | Curr-ency | Price Year | HEALTH Outcome Type | Health Result - Difference | Costs - Difference *as REPORTED* | Incremental CE Ratio *as REPORTED* | CONVERTED ICER (using 2022 US$)† | Cost-Benefit Analysis | Dis-count Rate | Uncertainty & Sensitivity Analyses |
|---|---|---|---|---|---|---|---|---|---|---|---|---|---|---|---|---|
| | | | | m, 325 mesh) | ation program | Belgium, Croatia, Czech Republic, Denmark, Finland, France, Germany, Greece, Iceland, Ireland, Israel, Italy, Luxembourg, Malta, Monaco, Netherlands, Norway, Portugal, San Marino, Slovenia, Spain, Sweden, Switzerland, United Kingdom) | | | | | | | | | health effects | and intervention effectiveness revealed that the results were quite robust to variation in the model parameters within +/-25%. Varying geographical coverage from 80% to 50% or 95% gave ICERs of I$ 7,574 or 5,573 per DALY averted, resp'ly. |
| Detzel 2016/ Patron 2016 | Iron | Infant food | NR | NR | No fortification | India | US$ | NR | Anemia (IDA) | 182,000 DALYs averted annually | -$461 million' | NR | N/A | NR | NR | Using estimates of efficacy from systematic reviews of clinical trials (instead of from a national health survey) found even greater reduction of DALYs and more cost savings. |
| Prieto-Patron 2022 | Iron | Infant cereals | 1 serving per day | NR | Unfortified infant food | Indonesia | US$ | 2020 | Anemia (IDA) | 43,000 DALYs averted | -$171,000,000 | NR | N/A | NR | 3% | PSA: much wider variability in the economic losses than in the intangible cost (DALYs), as in addition to the uncertainty of the health consequences of IDA, the economic |

| Study ID* (Author Year) | INTERVENTION- Micronutrient(s) | INTERVENTION- Food Vehicle(s) | Dose | Other Specs | Compar- ator | Country | Curr- ency | Price Year | HEALTH Outcome Type | Health Result - Difference | Costs - Difference *as REPORTED* | Incremental CE Ratio *as REPORTED* | CONVERTED ICER (using 2022 US$)† | Cost- Benefit Analysis | Dis- count Rate | Uncertainty & Sensitivity Analyses |
|---|---|---|---|---|---|---|---|---|---|---|---|---|---|---|---|---|
| | | | | | | | | | | | | | | | | parameters play an important role. |
| Baltussen 2004 | Iron | Cereal flours | 80% geographical coverage; dose NR | electrolytic iron (<45 microm, 325 mesh) | no iron fortification or supplementation program | South American subregion AmrB (Antigua And Barbuda, Argentina, Bahamas, Barbados, Belize, Brazil, Chile, Colombia, Costa Rica, Dominica, Dominican Republic, El Salvador, Grenada, Guyana, Honduras, Jamaica, Mexico, Panama, Paraguay, Saint Kitts And Nevis, Saint Lucia, Saint Vincent And The Grenadines, Suriname, Trinidad And Tobago, Uruguay, Venezuela) | international dollars | 2000 | DALY averted | 116,599 | I$ 16,510,432 | ICER I$ 142/DALY averted | 1,910 | NR | 3% for both costs and health effects | Uncertainty analysis focusing on the impact of varying assumptions about epidemiology, costs, and intervention effectiveness revealed that the results were quite robust to variation in the model parameters within +/-25%. Varying geographical coverage from 80% or 95% to: 50% or 95% gave ICERs of I$ 214 or 134 per DALY averted (resp'ly). |
| Baltussen 2004 | Iron | Cereal flours | 80% geographical coverage; dose NR | electrolytic iron (<45 microm, 325 mesh) | no iron fortification or supplementation program | Southeast Asian subregion SearD (Bangladesh, Bhutan, Democratic People's Republic Of Korea, India, | international dollars | 2000 | DALY averted | 939,284 | I$ 30,155,000 | ICER I$ 32/DALY averted | 47 | NR | 3% for both costs and health effects | Uncertainty analysis focusing on the impact of varying assumptions about epidemiology, costs, and intervention effectiveness revealed that the results were quite robust to variation in |

| Study ID* (Author Year) | INTERVENTION-Micronutrient(s) | INTERVENTION-Food Vehicle(s) | Dose | Other Specs | Compar-ator | Country | Curr-ency | Price Year | HEALTH Outcome Type | Health Result - Difference | Costs - Difference *as REPORTED* | Incremental CE Ratio *as REPORTED* | CONVERTED ICER (using 2022 US$)† | Cost-Benefit Analysis | Dis-count Rate | Uncertainty & Sensitivity Analyses |
|---|---|---|---|---|---|---|---|---|---|---|---|---|---|---|---|---|
| | | | | | | Maldives, Myanmar, Nepal ) | | | | | | | | | | the model parameters within +/-25%. Varying geographical coverage from 80% to: 50% or 95% gave ICERs of I$ 43 or 35 per DALY averted (resp'ly). |
| Asian Develop. Bank 2004 | Iron | Fish sauce | 60 parts per million (ppm) iron | Ferrous sulfate | Unfortified food | Thailand | US$ | NR | Deaths averted | 14 | 160,725,000 | NR | N/A | BCR: 6.6 | NR [Discounted costs and benefits] | NR |
| Asian Develop. Bank 2004 | Iron | Fish sauce | 60 parts per million (ppm) iron | NaFeE DTA | Unfortified food | Viet Nam | US$ | NR | Deaths averted | 800 | 257,825,000 | NR | N/A | BCR: 11.9 | NR [Discounted costs and benefits] | NR |
| Dainelli 2017 | Potassium | Milk powder product | 700mg/day | A milk powder product fortified with potassium | Unfortified product (same except for fortification ) | China | International dollar (intl$) | NR | QALYs gained | 0.26 | 1234 | ICER 4711.56 Int$ per QALY gained (best case) | 6,821 | NR | 3% for both costs and utilities | In sensitivity analyses varying parameters, results were overall robust. Fortification was the preferred strategy up to a (lower) effectiveness of potassium on systolic BP of 50%. Hospitalisation after stroke had the largest impact on final costs. |
| Edejer 2005 | Zinc | Wheat | 80% coverage; dose NR | Zinc oxide | No fortification | Afr-E (sub-Saharan Africa) region | international dollars ($Int) | 2000 | DALYs averted yearly (millions) | 0.13 | 8,000,000 yearly | ICER $ 60/DALY saved | 306 | NR | 3% for both costs and benefits | Removal of age weighting and discounting for DALYs makes the interventions more cost effective. Varying geographical coverage from 80% to: 50% or 95% |

| Study ID* (Author Year) | INTERVENTION-Micronutrient(s) | INTERVENTION-Food Vehicle(s) | Dose | Other Specs | Compar-ator | Country | Curr-ency | Price Year | HEALTH Outcome Type | Health Result - Difference | Costs - Difference *as REPORTED* | Incremental CE Ratio *as REPORTED* | CONVERTED ICER (using 2022 US$)† | Cost-Benefit Analysis | Dis-count Rate | Uncertainty & Sensitivity Analyses |
|---|---|---|---|---|---|---|---|---|---|---|---|---|---|---|---|---|
| | | | | | | | | | | | | | | | | gave ICERs of I$ 82 or 55 per DALY saved (resp'ly). |
| Vosti 2023 | Zinc | Wheat flour | 31.4 mg/kg | NR | No fortification | Cameroon | US$ | 2019 | Child lives saved | 903 | Total program costs US $ 2,003,000 | 2,218 | 2,274 | NR | NR | The program selection of "high" or "low" effectiveness results did not influence the key policy messages. |
| Vosti 2023 | Zinc | Wheat flour | 95 mg/kg | NR | No fortification | Cameroon | US$ | 2019 | Child lives saved | 10,604 | Total program costs US $ 6,070,000 | 572 | 586 | NR | NR | The program selection of "high" or "low" effectiveness results did not influence the key policy messages. |
| Vosti 2023 | Zinc | Bouillon cubes | 600 mg/kg | NR | No fortification | Cameroon | US$ | 2019 | Achieves adequate intake due to the intervention (effective coverage) | 9.2% | Total program costs US $ 5,583,000 | US$ 1.65 per child effectively covered | 2 | NR | NR | The program selection of "high" or "low" effectiveness results did not influence the key policy messages. |
| Ma 2008 | Zinc | NR [food fortification] | NR | NR | No fortification | China | international dollars ($Int) | 2000 | DALYs saved | NR | 0.01 per capita per year | ICER I$ 153/DALY saved | 385 | NR | NR | NR |
| Moges 2020 | Zinc | Wheat flour | Unclear (80mg in either 100g/1kg) | NR | Pre-fortification | Ethiopia | US$ | 2011 | Achieving adequate intake (effectively covered as shift from inadequacy) in young children plus women of reproductive age | 25,376,408 person-years | 59,251,806 | US$ 2.33 per effectively covered Child+WRA | 3.80 | NR | NR | NR |
| Edejer 2005 | Zinc | Wheat | 80% coverage; dose NR | Zinc oxide | No fortification | Sear-D (South East Asia) region | international dollars ($Int) | 2000 | DALYs averted yearly (millions) | 0.75 | 11,000,000 yearly | ICER $ 15/DALY saved | 22 | NR | 3% for both costs and benefits | Removal of age weighting and discounting for DALYs makes the interventions more cost effective. |

| Study ID* (Author Year) | INTERVENTION- Micronutrient(s) | INTERVENTION- Food Vehicle(s) | Dose | Other Specs | Compar- ator | Country | Curr- ency | Price Year | HEALTH Outcome Type | Health Result - Difference | Costs - Difference *as REPORTED* | Incremental CE Ratio *as REPORTED* | CONVERTED ICER (using 2022 US$)† | Cost- Benefit Analysis | Dis- count Rate | Uncertainty & Sensitivity Analyses |
|---|---|---|---|---|---|---|---|---|---|---|---|---|---|---|---|---|
| | | | | | | | | | | | | | | | | Varying geographical coverage from 80% to: 50% or 95% gave ICERs of I$ 19 or 14 per DALY saved (resp'ly). |

*See the 'List of Included Studies' IDs' above for the full bibliography.

†Costs in *THIS* column only have been converted to 2022 US$; where not specified, the ICER corresponds to the Health Outcome Type column (e.g., per DALY averted, etc.). "Dominant" ICER means the fortification was more effective AND less costly than the comparator.

Abbreviations: BCR=benefit-cost ratio; CBA=cost-benefit analysis; CE= cost-effectiveness; CEA= cost-effectiveness analysis; CUA= cost-utility analysis; DALY=disability-adjusted life year; ICER=incremental cost-effectiveness ratio; IU=international units; N/A=not applicable; NaFeEDTA=sodium iron ethylenediaminetetraacetate; NR=not reported; NTDs=neural tube defects; ppm=parts per million; QALY=quality-adjusted life year; Specs=specifications of fortification method; UIC=urinary iodine concentration; WRA=women of reproductive age.

***Table S3*: Hypothetical cost-effectiveness threshold summary results using ‘example’ percentages of GDP per capita (calculated per study country)**

| Country Economy Classification | <50% of GDP per capita (no. evaluations) | <35% of GDP per capita (no. evaluations) | <20% of GDP per capita (no. evaluations) | TOTAL no. of evaluations* |
|---|---|---|---|---|
| **LMICs (combined)** | **198** | **190** | **163** | **227** |
| *Low income* | *47* | *43* | *37* | *52* |
| *Lower middle income* | *124* | *121* | *103* | *141* |
| *Upper middle income* | *27* | *26* | *23* | *34* |
| **High income** | **3** | **3** | **1** | **5** |
| **ALL*** | **201** | **193** | **164** | **232** |

*Based on ICERs of cost per *DALY averted*.
Many studies reported relevant data on multiple evaluations (analyses) since they may have evaluated different interventions, countries, etc.
Abbreviations: GDP=gross domestic product; LMICs=low- and middle-income economy countries; no.=number.

### *Table S4*: Quality appraisal of models for each study [Philips' modeling framework]

### Page 1:

Structure: 'Structural' aspects relate to the scope and mathematical structure of the model including the strategies under evaluation.

| | Statement of decision problem / objective | | | Statement of scope and perspective | | | | Rationale for structure | | | | Structural assumptions | | Strategies / Comparators | | | Model type | Time horizon | | Disease states / pathways | Cycle length |
|---|---|---|---|---|---|---|---|---|---|---|---|---|---|---|---|---|---|---|---|---|---|
| **Study ID* (Author Year)** | S1.1: Is there a clear statement of decision problem? | S1.2: Is the objective of the evaluation and model specified and consistent with the decision problem? | S1.3: Is the primary decision maker specified? | S2.1: Is the perspective of the model stated clearly? | S2.2: Are the model inputs consistent with the stated perspective? | S2.3: Has the scope of the model been stated and justified? | S2.4: Are the outcomes of the model consistent with the perspective, scope and overall objective of the model? | S3.1: Has the evidence regarding the model structure been described? | S3.2: Is the structure of the model consistent with a coherence theory of the health condition under evaluation? | S3.3: Are the sources of data used to develop the structure of the model specified? | S3.4: Are the causal relationships described by the model structure justified appropriately? | S4.1: Are the structural assumptions transparent and justified? | S4.2: Are the structural assumptions reasonable given the overall objective, perspective and scope of the model? | S5.1: Is there a clear definition of the option under evaluation? | S5.2: Have all feasible and practical options been evaluated? | S5.3: Is there any justification for exclusion of feasible options? | S6.1: Is the chosen model type appropriate given the decision problem and specified causal relationships within the model? | S7.1: Is the time horizon of the model sufficient to reflect all the important differences between options? | S7.2: Is the time horizon of the model, and the duration of treatment and treatment effect described and justified? | S8.1: Do the disease states (state transition model) or the pathways (decision tree model) reflect the underlying biological process of the disease in question and the impact of interventions? | S9.1: Is the cycle length defined and justified in terms of the natural history of disease? |
| Access Economics 2006 | Yes | Yes | Yes | Yes | Yes | Yes | Yes | Yes | Yes | Yes | Yes | Yes | Yes | Yes | NA | Yes | Yes | Yes | Yes | Yes | ? |
| Asian Develop. Bank 2004 | Yes | Yes | Yes/No | Yes | Yes | Yes | Yes | Yes | Yes | Yes | Yes | Yes | Yes | Yes | NA | Yes | Yes | Yes | Yes | Yes | ? |
| Australian HMAC 2017 | Yes | Yes | Yes | Yes | Yes | Yes | Yes | Yes | Yes | Yes | Yes | Yes | Yes | Yes | NA | Yes | Yes | Yes | Yes | Yes | ? |
| Baltussen 2004 | Yes | Yes | Yes | Yes | Yes | Yes | Yes | Yes | Yes | Yes | No | Yes/No | Yes | Yes | Yes | ? | Yes | Yes | Yes/No | Yes | ? |
| Bentley 2009 | Yes | Yes | No | No | Yes | Yes | Yes | Yes | Yes | Yes | Yes | Yes | Yes | Yes | NA | Yes | Yes | Yes | Yes | Yes | ? |
| Chow 2010 | No | NA | Yes/No | No | NA | No | NA | Yes | Yes | Yes/No | Yes/No | Yes | Yes | Yes | NA | NA | Yes | Yes | No | Yes | ? |
| Connelly 1996 | Yes | Yes | Yes | No | NA | Yes | Yes | Yes | Yes | Yes | Yes | Yes | Yes | Yes | NA | NA | Yes | Yes | Yes | Yes | ? |
| Dainelli 2017 | Yes | Yes | No | Yes | Yes | Yes | Yes | Yes | Yes | Yes | Yes | Yes | Yes | Yes | ? | No | Yes | Yes | Yes | Yes | ? |
| Dalziel 2010/Segal 2007 | Yes | Yes | Yes | Yes | Yes | Yes | Yes | No | Yes | Yes | Yes | Yes | Yes | Yes | ? | No | Yes | Yes | Yes | Yes | ? |
| Detzel 2016 | Yes | Yes | No | Yes | Yes | Yes/No | Yes | No | Yes/No | Yes | No | ? | ? | Yes/No | Yes | Yes | Yes | ? | ? | ? | ? |
| Edejer 2005 | Yes | Yes | Yes/No | Yes | Yes | Yes | Yes | No | Yes | Yes | Yes | Yes | Yes | Yes | ? | Yes | Yes | Yes | Yes | Yes | ? |
| Fiedler 2000 | Yes | Yes | Yes | No | NA | Yes/No | Yes | Yes/No | Yes | Yes/No | Yes/No | Yes | Yes | Yes | NA | NA | Yes | No | No | Yes | ? |
| Fiedler 2009 | Yes | Yes | Yes/No | Yes | Yes | Yes | Yes | Yes | Yes | Yes | Yes | Yes | Yes | Yes | Yes | ? | Yes | Yes | No | Yes | ? |
| Fiedler 2010 | Yes | Yes | Yes | Yes | Yes | Yes | Yes | No | Yes | Yes | Yes | Yes | Yes | Yes | ? | No | Yes | ? | No | Yes | ? |
| Fiedler 2012 | Yes | Yes | Yes/No | No | NA | Yes/No | Yes | Yes | Yes | Yes/No | Yes/No | Yes/No | Yes | Yes | NA | NA | Yes | ? | ? | Yes | ? |
| Fiedler 2013 | Yes | Yes | Yes | No | ? | Yes/No | ? | Yes/No | Yes | Yes/No | Yes/No | Yes | ? | Yes | Yes | No | Yes | ? | No | Yes | ? |
| Fiedler 2014 | Yes | Yes | Yes/No | Yes/No | Yes/No | Yes | Yes/No | Yes/No | Yes | Yes | Yes/No | Yes | Yes/No | Yes | Yes | No | Yes | Yes | Yes | Yes | ? |
| Fiedler 2015 | Yes | Yes | Yes | No | ? | Yes | NA | Yes | Yes | Yes | Yes | Yes | NA | Yes | Yes | NA | Yes | ? | No | Yes | ? |
| GAIN 2017 | Yes | Yes | No | No | Yes | Yes | Yes | Yes/No | Yes | Yes/No | Yes/No | Yes/No | ? | Yes/No | ? | No | Yes | Yes | No | Yes | ? |
| Ghauri 2015 | Yes | Yes | No | No | Yes | Yes | Yes | Yes/No | Yes | Yes | Yes/No | Yes/No | Yes | Yes/No | NA | Yes | Yes | Yes/No | Yes/No | Yes | ? |
| Ghauri 2016 | Yes | Yes | No | No | Yes | Yes | Yes | Yes/No | Yes | Yes | Yes/No | Yes/No | Yes | Yes/No | NA | Yes | Yes | Yes/No | Yes/No | Yes | ? |
| Gorstein 2020 | Yes | Yes | Yes/No | No | ? | Yes/No | ? | Yes | Yes | Yes | Yes/No | Yes/No | ? | Yes | No | No | ? | Yes | Yes | Yes | ? |
| Grosse 2016 | Yes | Yes | Yes | Yes | Yes | Yes | Yes | No | Yes | Yes | Yes | Yes | Yes | Yes | ? | No | Yes | Yes | Yes | Yes | ? |
| Hoddinott 2018 | Yes | Yes | No | No | ? | Yes/No | ? | No | Yes | Yes | Yes | Yes/No | ? | Yes | ? | No | ? | Yes | Yes | Yes | ? |
| Horton 2003 | Yes | Yes | Yes/No | No | ? | Yes | ? | Yes/No | Yes | Yes | Yes | Yes/No | ? | Yes | Yes | No | Yes | Yes | Yes | Yes | ? |
| Horton 2008 | Yes | Yes | Yes/No | Yes | Yes | Yes | Yes | No | Yes | Yes | Yes/No | Yes | Yes | Yes | ? | Yes | Yes | Yes | Yes | Yes | ? |
| Huang 2020 | Yes | Yes | No | No | Yes | Yes | Yes | Yes/No | Yes/No | Yes/No | Yes/No | Yes/No | Yes/No | Yes | NA | Yes | Yes/No | ? | No | Yes | ? |
| Jentink 2008 | Yes | Yes | Yes/No | Yes | Yes | Yes | Yes | Yes | Yes | Yes/No | Yes | Yes | Yes | Yes | NA | Yes | Yes | Yes | Yes | Yes | ? |
| Johnson 2021 | Yes | Yes | Yes | Yes | Yes | Yes | Yes | Yes | Yes | Yes | Yes | Yes | Yes | Yes | NA | Yes | Yes | Yes | Yes | Yes | ? |
| Kakietek 2018 | Yes | Yes | Yes | Yes | Yes | Yes | Yes | Yes | Yes | Yes | Yes | Yes | Yes | Yes/No | NA | Yes | Yes | Yes/No | Yes | Yes | ? |
| Kancherla 2021 | Yes | Yes | Yes/No | No | Yes | Yes | Yes | Yes | Yes | Yes | Yes/No | Yes | Yes | Yes | NA | Yes | Yes | Yes | Yes | Yes | ? |
| Llanos 2007 | Yes | Yes | Yes | No | Yes | Yes | Yes | Yes | Yes | Yes | Yes | Yes | Yes | Yes | NA | Yes | Yes | ? | ? | Yes | ? |
| Ma 2008 | Yes | Yes | Yes | No | Yes | Yes | Yes/No | Yes/No | Yes | Yes | Yes/No | Yes/No | Yes | Yes/No | Yes | Yes | ? | ? | Yes/No | Yes | ? |
| Mardones-Santander 1991 | Yes | Yes | Yes | Yes | Yes | Yes | Yes | Yes/No | Yes | Yes | Yes | Yes | Yes | Yes | Yes | Yes | Yes | Yes | Yes | Yes | ? |
| Moges 2020/Vosti 2020b | Yes | Yes | Yes | No | ? | Yes | Yes | Yes/No | Yes | Yes | Yes | Yes | Yes | Yes | ? | No | Yes | Yes | Yes | Yes | ? |
| MQSUN 2014 | Yes | Yes | No | No | ? | No | ? | Yes/No | Yes | Yes | Yes | No | ? | Yes | ? | Yes | Yes | Yes/No | ? | Yes | ? |
| Niedermaier 2021 | Yes | Yes | Yes/No | No | Yes | Yes | Yes | No | Yes | Yes | Yes | Yes | Yes | Yes | NA | Yes | Yes | Yes | ? | Yes | ? |
| Niemesh 2015 | Yes | Yes | No | No | Yes | Yes | Yes | Yes | Yes | Yes | Yes/No | Yes | Yes | Yes | NA | Yes | Yes | Yes | Yes/No | Yes | ? |
| Noshirvan 2021 | Yes | Yes | Yes | No | ? | Yes | ? | Yes | Yes | Yes | Yes | Yes | ? | Yes | ? | No | Yes | Yes | Yes | Yes | ? |
| Palacios 2022 | Yes | Yes | Yes | No | Yes | Yes | Yes | Yes | Yes | Yes | NA | Yes | Yes | Yes | NA | Yes | Yes | Yes | NA | Yes | ? |
| Pandav 2012 | Yes | Yes | Yes | Yes | Yes | Yes | Yes | Yes/No | Yes | Yes | Yes | Yes | Yes | Yes | NA | Yes | Yes | Yes | Yes | Yes | ? |
| Phillips 1996 | Yes | Yes | No | No | NA | Yes | NA | No | No | Yes/No | Yes | Yes/No | NA | Yes | ? | No | ? | ? | No | Yes | ? |
| Popkin 1980 | Yes | Yes | Yes/No | Yes | Yes | Yes | Yes | Yes/No | Yes | Yes | Yes | Yes | Yes | Yes | NA | Yes | Yes | Yes | Yes | Yes | ? |
| Prieto-Patron 2022 | Yes | Yes | Yes | Yes/No | Yes/No | Yes | Yes | Yes | Yes/No | Yes | No | Yes | Yes | Yes | ? | No | Yes | Yes | Yes | Yes | ? |
| Qureshy 2023 | Yes | Yes | Yes | No | Yes | Yes | Yes | Yes | Yes | Yes | Yes | Yes | Yes | Yes | NA | Yes | Yes | Yes | Yes | Yes | ? |
| Rabovskaja 2013 | Yes | Yes | Yes | Yes | Yes | Yes | Yes | Yes | Yes | Yes | Yes | Yes | Yes | Yes | NA | Yes | Yes | Yes | Yes | Yes | ? |
| Rajkumar 2012 | Yes | Yes | Yes/No | Yes/No | Yes | Yes | Yes | Yes/No | Yes | Yes | Yes | Yes | Yes | Yes/No | Yes | NA | Yes/No | ? | ? | Yes | ? |
| Rochau 2019 | Yes | Yes | No | No | Yes | No | Yes | No | ? | Yes | No | No | ? | Yes/No | ? | No | Yes | Yes | Yes | Yes | ? |
| Sablah 2012 | Yes | Yes | Yes/No | No | Yes/No | Yes/No | Yes | Yes/No | Yes/No | No | Yes/No | No | Yes/No | Yes/No | NA | Yes | ? | ? | ? | Yes | ? |
| Sandmann 2017 | Yes | Yes | Yes | Yes | Yes | Yes | Yes | Yes | Yes | Yes | Yes | Yes | Yes | Yes | NA | Yes | Yes | Yes | Yes | Yes | ? |
| Vosti 2023/Adams 2022 | Yes | Yes | Yes | No | Yes | Yes | Yes | Yes/No | Yes | Yes | Yes | Yes | Yes | Yes | Yes | ? | Yes | Yes | Yes | Yes | ? |
| Walters 2019 | Yes | Yes | Yes | Yes | Yes | Yes | Yes | Yes | Yes | Yes | Yes | Yes/No | Yes | Yes | Yes | Yes | Yes | Yes/No | Yes | Yes | ? |
| Wei 2023 | Yes | Yes | Yes | No | Yes | Yes | Yes | Yes/No | Yes | Yes/No | NA | Yes | Yes | Yes/No | NA | Yes | Yes | Yes | Yes/No | Yes | ? |

### Page 2:

| | Data: 'Data' issues include data identification methods and how uncertainty should be addressed. Data identification | | | | | | Pre-model data analysis | Pre-model: Baseline data | | | Pre-model: Treatment effects | | | | | Pre-model: Quality of life weights (utility) | | |
|---|---|---|---|---|---|---|---|---|---|---|---|---|---|---|---|---|---|---|
| **Study ID* (Author Year)** | D1.1: Are the data identification methods transparent and appropriate given the objectives of the model? | D1.2: Where choices have been made between data sources, are these justified appropriately? | D1.3: Has particular attention been paid to identifying data for the important parameters in the model? | D1.4: Has the process of selecting key parameters been justified and systematic methods used to identify the most appropriate data? | D1.5: Has the quality of the data been assessed appropriately? | D1.6: Where expert opinion has been used, are the methods described and justified? | D2.1: Are the pre-model data analysis method based on justifiable statistical and epidemiological techniques? | D2.a1: Is the choice of baseline data described and justified? | D2.a2: Are transition probabilities calculated appropriately? | D2.a3: Has a half cycle correction been applied to both cost and outcome; or if not, has this omission been justified? | D2.b1: If relative treatment effects have been derived from trial data, have they been synthesised using appropriate techniques? | D2.b2: Have the methods and assumptions used to extrapolate short-term results to final outcomes been documented and justified? | D2.b3: Have alternative extrapolation assumptions been explored through sensitivity analysis? | D2.b4: Have assumptions regarding the continuing effect of treatment once treatment is complete been documented and justified? | D2.b5: Have alternative assumptions regarding the continuing effect of treatment been explored through sensitivity analysis? | D2.c1: Are the utilities incorporated into the model appropriate? | D2.c2: Is the source for the utility weights referenced? | D2.c3: Are the methods of derivation for the utility weights justified? |
| Access Economics 2006 | Yes | Yes | Yes | Yes | Yes | NA | Yes | Yes | Yes | ? | NA | Yes | Yes | Yes | No | NA | NA | NA |
| Asian Develop. Bank 2004 | Yes | Yes | Yes | Yes | ? | ? | Yes | Yes | Yes | ? | NA | Yes | No | NA | No | NA | NA | NA |
| Australian HMAC 2017 | Yes | Yes | Yes | Yes | Yes | NA | Yes | Yes | Yes | ? | NA | Yes | Yes | Yes | Yes | Yes | Yes | Yes |
| Baltussen 2004 | Yes | ? | Yes | Yes | Yes | Yes/No | Yes | Yes | Yes | ? | Yes | NA | NA | No | No | NA | NA | NA |
| Bentley 2009 | Yes | Yes | Yes | Yes | ? | NA | Yes | Yes | Yes | ? | NA | Yes | Yes | Yes | Yes | Yes | Yes | Yes |
| Chow 2010 | No | NA | ? | No | ? | NA | ? | ? | NA | ? | NA | NA | Yes | No | Yes | NA | NA | NA |
| Connelly 1996 | Yes | Yes | Yes | Yes | ? | NA | ? | Yes | Yes | ? | NA | Yes | Yes | No | Yes | NA | NA | NA |
| Dainelli 2017 | No | NA | Yes | No | ? | NA | Yes | Yes | ? | ? | ? | ? | No | No | No | Yes | Yes | Yes |
| Dalziel 2010/Segal 2007 | Yes | Yes | Yes | Yes | Yes | No | Yes | Yes | Yes | ? | ? | ? | No | No | No | NA | NA | NA |
| Detzel 2016 | Yes | ? | ? | No | ? | NA | ? | No | ? | ? | NA | ? | No | No | No | NA | NA | NA |
| Edejer 2005 | Yes | Yes/No | Yes | Yes/No | ? | Yes | Yes | Yes | ? | ? | ? | No | No | Yes | No | NA | NA | NA |
| Fiedler 2000 | Yes/No | Yes/No | Yes | ? | Yes/No | No | ? | Yes | NA | ? | NA | NA | Yes | No | No | NA | NA | NA |
| Fiedler 2009 | Yes | Yes | Yes | Yes/No | ? | ? | Yes | Yes | ? | ? | Yes | Yes | No | Yes | No | NA | NA | NA |
| Fiedler 2010 | No | Yes | Yes/No | No | ? | NA | Yes | Yes | ? | ? | ? | ? | No | No | No | NA | NA | NA |
| Fiedler 2012 | Yes/No | Yes/No | ? | Yes | ? | No | Yes | Yes | ? | ? | NA | ? | No | No | No | NA | NA | NA |
| Fiedler 2013 | Yes | Yes | Yes | Yes | ? | No | Yes | Yes | ? | ? | Yes | Yes | No | No | No | NA | NA | NA |
| Fiedler 2014 | Yes | NA | Yes | Yes/No | Yes | No | Yes | Yes | ? | ? | NA | ? | No | No | No | NA | NA | NA |
| Fiedler 2015 | Yes | NA | Yes | Yes | Yes | NA | Yes | Yes | Yes | ? | NA | Yes | No | No | No | NA | NA | NA |
| GAIN 2017 | Yes | ? | ? | ? | ? | ? | ? | Yes | ? | ? | NA | Yes/No | No | Yes/No | No | NA | NA | NA |
| Ghauri 2015 | Yes/No | Yes/No | Yes | Yes/No | Yes/No | Yes/No | Yes | Yes | Yes/No | ? | NA | Yes/No | No | Yes/No | No | NA | NA | NA |
| Ghauri 2016 | Yes/No | Yes/No | Yes | Yes/No | Yes/No | Yes/No | Yes | Yes | Yes/No | ? | NA | Yes/No | No | NA | No | NA | NA | NA |
| Gorstein 2020 | Yes/No | NA | Yes | Yes | ? | NA | Yes | Yes | ? | ? | NA | ? | No | No | No | NA | NA | NA |
| Grosse 2016 | No | NA | Yes | No | ? | ? | Yes | Yes | ? | ? | ? | ? | No | No | No | NA | NA | NA |
| Hoddinott 2018 | Yes/No | NA | Yes/No | Yes/No | ? | NA | Yes | Yes | ? | ? | ? | ? | Yes | No | No | NA | NA | NA |
| Horton 2003 | Yes | Yes | Yes | No | ? | NA | Yes | Yes | ? | ? | NA | ? | No | No | No | NA | NA | NA |
| Horton 2008 | No | Yes | Yes | Yes/No | ? | NA | Yes | Yes | ? | ? | ? | ? | No | Yes | No | NA | NA | NA |
| Huang 2020 | ? | ? | ? | ? | ? | NA | ? | No | ? | ? | NA | ? | Yes | ? | No | NA | NA | NA |
| Jentink 2008 | Yes | Yes | Yes | Yes | Yes | Yes | Yes | Yes | Yes | ? | NA | Yes | Yes | Yes | Yes | Yes | Yes | ? |
| Johnson 2021 | Yes | Yes | Yes | Yes | Yes | NA | Yes | Yes | Yes | ? | NA | Yes/No | No | NA | No | NA | NA | NA |
| Kakietek 2018 | Yes | Yes | Yes | Yes | Yes/No | NA | Yes | Yes | Yes | ? | NA | Yes | No | NA | No | NA | NA | NA |
| Kancherla 2021 | Yes | Yes | Yes | Yes | ? | No | ? | Yes | ? | ? | NA | Yes/No | Yes | No | No | NA | NA | NA |
| Llanos 2007 | Yes | Yes | Yes | Yes | ? | NA | ? | Yes | ? | ? | NA | ? | Yes | ? | No | NA | NA | NA |
| Ma 2008 | Yes | NA | Yes | Yes | ? | NA | ? | Yes | ? | ? | NA | ? | No | ? | No | NA | NA | NA |
| Mardones-Santander 1991 | Yes | NA | Yes | Yes | Yes | NA | Yes | Yes | Yes | ? | Yes | NA | No | NA | NA | NA | NA | NA |
| Moges 2020/Vosti 2020b | Yes | Yes | Yes | Yes/No | ? | NA | Yes | Yes | ? | ? | ? | ? | Yes | No | No | NA | NA | NA |
| MQSUN 2014 | No | Yes/No | Yes | Yes/No | ? | NA | Yes | Yes | ? | ? | ? | ? | No | Yes | No | NA | NA | NA |
| Niedermaier 2021 | Yes | Yes | Yes | Yes | Yes | NA | ? | Yes | Yes | ? | NA | Yes | Yes | Yes | No | NA | NA | NA |
| Niemesh 2015 | Yes | Yes | Yes | Yes | ? | NA | ? | Yes | ? | ? | NA | ? | No | ? | No | NA | NA | NA |
| Noshirvan 2021 | Yes | Yes | Yes | Yes/No | ? | NA | Yes | Yes | ? | ? | NA | Yes | Yes | Yes | Yes | NA | NA | NA |
| Palacios 2022 | Yes | NA | Yes | Yes | ? | NA | Yes | Yes | NA | ? | NA | NA | No | NA | No | NA | NA | NA |
| Pandav 2012 | Yes | Yes | Yes | Yes | ? | ? | Yes | Yes | Yes | ? | NA | ? | Yes | ? | Yes | NA | NA | NA |
| Phillips 1996 | No | Yes/No | No | No | Yes | NA | No | Yes | ? | ? | NA | ? | Yes | No | No | NA | NA | NA |
| Popkin 1980 | Yes | NA | Yes | Yes | Yes | NA | Yes | Yes | Yes | ? | Yes | Yes | Yes | Yes | No | NA | NA | NA |
| Prieto-Patron 2022 | No | No | Yes | No | ? | NA | Yes | Yes | NA | ? | NA | Yes | No | Yes | No | NA | NA | NA |
| Qureshy 2023 | Yes | Yes | Yes | Yes | Yes | NA | Yes | Yes | Yes | ? | NA | Yes | Yes | Yes | Yes | NA | NA | NA |
| Rabovskaja 2013 | Yes | NA | Yes | Yes | Yes | NA | Yes | Yes | Yes | ? | NA | Yes | Yes | Yes | Yes | Yes | Yes | Yes |
| Rajkumar 2012 | Yes | NA | Yes | Yes | ? | NA | Yes | Yes | Yes | ? | NA | Yes/No | No | No | No | NA | NA | NA |
| Rochau 2019 | Yes/No | No | ? | No | ? | No | ? | No | ? | ? | ? | No | Yes | No | Yes | ? | No | ? |
| Sablah 2012 | ? | ? | ? | ? | ? | ? | ? | ? | ? | ? | NA | ? | No | No | No | NA | NA | NA |
| Sandmann 2017 | Yes | ? | Yes | Yes | Yes | NA | Yes | Yes | Yes | ? | NA | Yes | Yes | Yes | Yes | NA | NA | NA |
| Vosti 2023/Adams 2022 | Yes | Yes | Yes | Yes/No | Yes | NA | Yes | Yes | ? | ? | NA | ? | Yes | No | No | NA | NA | NA |
| Walters 2019 | Yes | Yes | Yes | NA | ? | NA | Yes | Yes | ? | ? | NA | ? | No | No | No | NA | NA | NA |
| Wei 2023 | Yes | NA | NA | NA | ? | NA | Yes | Yes | NA | ? | NA | NA | Yes | NA | NA | NA | NA | NA |

### Page 3:

Data: 'Data' issues include data identification methods and how uncertainty should be addressed.

| | Data incorporation | | | | | Assessment of uncertainty | Uncertainty: Methodological | Uncertainty: Structural | Jncertainty: Heterogeneit | Uncertainty: Parameter | |
|---|---|---|---|---|---|---|---|---|---|---|---|
| **Study ID* (Author Year)** | D3.1: Have all data incorporated into the model been described and referenced in sufficient detail? | D3.2: Has the use of mutually inconsistent data been justified (i.e. are assumptions and choices appropriate)? | D3.3: Is the process of data incorporation transparent? | D3.4: If data have been incorporated as distributions, has the choice of distribution for each parameter been described and justified? | D3.5: If data have been incorporated as distribution, is it clear that second order uncertainty is reflected? | D4.1: Have the four principal types of uncertainty been addressed; or if not, has the omission of particular forms of uncertainty been justified? | D4.a1: Have methodological uncertainties been addressed by running alternative versions of the model with different methodological assumptions? | D4.b1: Is there evidence that structural uncertainties have been addressed via sensitivity analysis? | D4.c1: Has heterogeneity been dealt with by running the model separately for different sub-groups? | D4.d1: Are the methods of assessment of parameter uncertainty appropriate? | D4.d2: If data are incorporated as point estimates, are the range used for sensitivity analysis stated clearly and justified? |
| Access Economics 2006 | Yes | NA | Yes | Yes | NA | Yes | Yes | Yes | Yes | ? | Yes |
| Asian Develop. Bank 2004 | Yes | NA | Yes | NA | NA | Yes/No | No | No | Yes | ? | NA |
| Australian HMAC 2017 | Yes | NA | Yes | Yes | NA | Yes | Yes | Yes | Yes | ? | Yes |
| Baltussen 2004 | Yes | Yes | Yes | ? | ? | Yes/No | No | No | Yes | Yes | Yes |
| Bentley 2009 | Yes | NA | Yes | NA | NA | Yes | Yes | Yes | Yes | ? | Yes |
| Chow 2010 | No | NA | Yes | Yes | NA | Yes/No | No | No | Yes | Yes | NA |
| Connelly 1996 | Yes | NA | Yes | NA | NA | Yes/No | No | No | Yes | NA | Yes |
| Dainelli 2017 | Yes | Yes | Yes | No | No | Yes/No | Yes | Yes | No | Yes | NA |
| Dalziel 2010/Segal 2007 | Yes | Yes | Yes | NA | NA | Yes/No | Yes | Yes | No | Yes | Yes |
| Detzel 2016 | No | NA | ? | NA | NA | No | No | No | No | NA | NA |
| Edejer 2005 | Yes | NA | Yes | NA | NA | Yes/No | Yes | Yes | Yes | No | NA |
| Fiedler 2000 | Yes/No | NA | Yes | NA | NA | Yes/No | Yes | No | Yes | Yes | Yes/No |
| Fiedler 2009 | Yes | ? | Yes/No | NA | NA | No | No | No | No | No | No |
| Fiedler 2010 | Yes | NA | Yes/No | NA | NA | No | No | No | Yes | NA | No |
| Fiedler 2012 | Yes/No | NA | Yes | NA | NA | Yes/No | No | No | No | NA | NA |
| Fiedler 2013 | Yes/No | NA | Yes | NA | NA | Yes/No | Yes | Yes | No | No | No |
| Fiedler 2014 | Yes | NA | Yes | NA | NA | Yes/No | Yes | Yes | No | Yes | No |
| Fiedler 2015 | Yes | Yes | Yes | NA | NA | No | No | No | No | NA | NA |
| GAIN 2017 | Yes/No | NA | Yes/No | NA | NA | No | No | No | No | ? | NA |
| Ghauri 2015 | Yes | NA | Yes/No | NA | NA | No | No | No | No | ? | NA |
| Ghauri 2016 | Yes | NA | Yes/No | NA | NA | No | No | No | No | ? | NA |
| Gorstein 2020 | Yes | NA | Yes | NA | NA | No | No | No | No | No | No |
| Grosse 2016 | Yes | NA | Yes | NA | NA | Yes/No | No | Yes | No | No | Yes |
| Hoddinott 2018 | Yes | NA | Yes/No | NA | NA | Yes/No | No | Yes | No | Yes | Yes |
| Horton 2003 | Yes | NA | Yes | NA | NA | No | No | No | No | No | No |
| Horton 2008 | Yes | NA | Yes/No | NA | NA | Yes/No | Yes | Yes | No | No | No |
| Huang 2020 | Yes | NA | ? | ? | NA | Yes/No | ? | No | No | ? | ? |
| Jentink 2008 | Yes | NA | Yes | Yes | ? | Yes/No | Yes | Yes | No | ? | Yes |
| Johnson 2021 | Yes | NA | Yes/No | NA | NA | No | No | No | No | ? | NA |
| Kakietek 2018 | Yes | NA | Yes | NA | NA | Yes/No | No | No | Yes | ? | Yes |
| Kancherla 2021 | Yes/No | NA | Yes | NA | NA | Yes/No | Yes | Yes | No | ? | Yes |
| Llanos 2007 | Yes/No | NA | Yes | NA | NA | Yes/No | Yes | Yes | No | ? | Yes |
| Ma 2008 | Yes/No | NA | Yes | NA | NA | No | No | No | No | ? | NA |
| Mardones-Santander 1991 | Yes | NA | Yes | NA | NA | No | No | No | No | ? | NA |
| Moges 2020/Vosti 2020b | Yes | NA | Yes | NA | NA | Yes/No | Yes | Yes | Yes | Yes | No |
| MQSUN 2014 | Yes | NA | Yes | NA | NA | No | No | No | No | No | No |
| Niedermaier 2021 | Yes | NA | Yes | NA | NA | Yes/No | Yes | Yes | No | ? | Yes |
| Niemesh 2015 | Yes | NA | Yes | NA | NA | No | No | No | No | ? | NA |
| Noshirvan 2021 | Yes | NA | Yes/No | NA | NA | Yes/No | No | Yes | Yes | Yes | Yes |
| Palacios 2022 | Yes | NA | Yes | NA | NA | No | No | No | No | ? | NA |
| Pandav 2012 | Yes | NA | Yes | NA | NA | Yes/No | Yes | Yes | No | ? | Yes |
| Phillips 1996 | Yes/No | NA | Yes/No | NA | NA | Yes/No | No | No | No | ? | Yes |
| Popkin 1980 | Yes | NA | Yes | NA | NA | Yes/No | Yes | Yes | Yes | ? | ? |
| Prieto-Patron 2022 | Yes | Yes | Yes | Yes | Yes/No | Yes/No | No | No | No | Yes | Yes/No |
| Qureshy 2023 | Yes | NA | Yes | Yes | NA | Yes/No | Yes | Yes | No | ? | Yes |
| Rabovskaja 2013 | Yes | NA | Yes | Yes | NA | Yes/No | Yes | Yes | No | ? | Yes |
| Rajkumar 2012 | Yes | NA | Yes | NA | NA | No | No | No | No | ? | NA |
| Rochau 2019 | No | ? | No | NA | NA | Yes/No | Yes/No | Yes | Yes | ? | ? |
| Sablah 2012 | No | NA | No | ? | NA | No | No | No | No | ? | ? |
| Sandmann 2017 | Yes | NA | Yes | Yes | NA | Yes/No | Yes | Yes | No | Yes | Yes |
| Vosti 2023/Adams 2022 | Yes | NA | Yes | NA | NA | Yes/No | Yes | Yes | Yes | Yes | No |
| Walters 2019 | Yes | NA | Yes | NA | NA | Yes/No | No | Yes | No | Yes | No |
| Wei 2023 | Yes/No | NA | Yes | NA | NA | Yes/No | Yes | Yes | No | ? | Yes/No |

**Page 4:**

| | Consistency: 'Consistency' relates to the overall quality of the model. | | | | |
|---|---|---|---|---|---|
| | Internal consistency | External consistency | | | |
| **Study ID* (Author Year)** | C1.1: Is there evidence that the mathematical logic of the model has been tested thoroughly before use? | C2.1: Are the conclusions valid given the data presented? | C2.2: Are any counterintuitive results from the model explained and justified? | C2.3: If the model has been calibrated against independent data, have any differences been explained and justified? | C2.4: Have the results of the model been compared with those of previous models and any differences in results explained? |
| Access Economics 2006 | Yes | Yes | NA | ? | Yes |
| Asian Develop. Bank 2004 | Yes | Yes | NA | ? | Yes |
| Australian HMAC 2017 | Yes | Yes | NA | ? | Yes |
| Baltussen 2004 | No | Yes | Yes | ? | No |
| Bentley 2009 | No | Yes | NA | ? | Yes |
| Chow 2010 | No | Yes | Yes | ? | No |
| Connelly 1996 | ? | Yes | NA | ? | No |
| Dainelli 2017 | No | Yes | NA | ? | Yes |
| Dalziel 2010/Segal 2007 | No | Yes | NA | ? | Yes |
| Detzel 2016 | No | Yes | NA | ? | No |
| Edejer 2005 | No | Yes | Yes | ? | Yes |
| Fiedler 2000 | No | Yes | NA | ? | No |
| Fiedler 2009 | ? | Yes | Yes/No | ? | Yes |
| Fiedler 2010 | ? | Yes | NA | ? | No |
| Fiedler 2012 | Yes | Yes | NA | ? | No |
| Fiedler 2013 | ? | Yes | Yes | ? | ? |
| Fiedler 2014 | No | Yes | Yes | ? | Yes |
| Fiedler 2015 | Yes | Yes | Yes | ? | No |
| GAIN 2017 | No | Yes | NA | ? | No |
| Ghauri 2015 | No | Yes | NA | ? | No |
| Ghauri 2016 | No | Yes | NA | ? | No |
| Gorstein 2020 | No | Yes | Yes | ? | Yes |
| Grosse 2016 | No | Yes | NA | ? | Yes |
| Hoddinott 2018 | No | Yes | NA | ? | No |
| Horton 2003 | No | Yes | Yes | ? | Yes |
| Horton 2008 | No | Yes | No | ? | Yes |
| Huang 2020 | No | Yes | NA | ? | No |
| Jentink 2008 | No | Yes | NA | ? | Yes |
| Johnson 2021 | No | Yes | NA | ? | No |
| Kakietek 2018 | Yes | Yes | NA | ? | Yes |
| Kancherla 2021 | No | Yes | NA | ? | Yes/No |
| Llanos 2007 | Yes | Yes | NA | ? | Yes |
| Ma 2008 | No | Yes | NA | ? | No |
| Mardones-Santander 1991 | Yes | Yes | NA | ? | No |
| Moges 2020/Vosti 2020b | Yes | Yes | Yes | ? | Yes |
| MQSUN 2014 | No | Yes | Yes/No | ? | Yes |
| Niedermaier 2021 | No | Yes | Yes | ? | Yes |
| Niemesh 2015 | No | Yes | NA | ? | Yes |
| Noshirvan 2021 | No | Yes | Yes | ? | Yes |
| Palacios 2022 | ? | Yes | NA | ? | Yes |
| Pandav 2012 | No | Yes | NA | ? | Yes |
| Phillips 1996 | No | Yes | NA | ? | No |
| Popkin 1980 | No | Yes | NA | ? | No |
| Prieto-Patron 2022 | No | Yes | Yes | ? | Yes |
| Qureshy 2023 | Yes/No | Yes | NA | ? | Yes |
| Rabovskaja 2013 | No | Yes | Yes | ? | Yes |
| Rajkumar 2012 | No | Yes | NA | ? | Yes |
| Rochau 2019 | No | No | NA | ? | No |
| Sablah 2012 | No | ? | NA | ? | No |
| Sandmann 2017 | Yes | Yes | NA | ? | Yes |
| Vosti 2023/Adams 2022 | Yes | Yes | Yes | ? | Yes |
| Walters 2019 | No | Yes | Yes | ? | Yes |
| Wei 2023 | No | Yes | NA | ? | Yes |

*See the 'List of Included Studies' IDs' above for the full bibliography.

***Table S5*: Quality appraisal of primary studies [using Evers CHEC-list]**

| Study ID* (Author Year) | 1. Is the study population clearly described? | 2. Are competing alternatives clearly described? | 3. Is a well-defined research question posed in answerable form? | 4. Is the economic study design appropriate to the stated objective? | 5. Is the chosen time horizon appropriate in order to include relevant costs and consequences? | 6. Is the actual perspective chosen appropriate? | 7. Are all important and relevant costs for each alternative identified? | 8. Are all costs measured appropriately in physical units? | 9. Are costs valued appropriately? | 10. Are all important and relevant outcomes for each alternative identified? | 11. Are all outcomes measured appropriately? | 12. Are outcomes valued appropriately? | 13. Is an incremental analysis of costs and outcomes of alternatives performed? | 14. Are all future costs and outcomes discounted appropriately? | 15. Are all important variables, whose values are uncertain, appropriately subjected to sensitivity analysis? | 16. Do the conclusions follow from the data reported? | 17. Does the study discuss the generalizability of the results to other settings and patient/client groups? | 18. Does the article indicate that there is no potential conflict of interest of study researcher(s) and funder(s)? | 19. Are ethical and distributional issues discussed appropriately? |
|---|---|---|---|---|---|---|---|---|---|---|---|---|---|---|---|---|---|---|---|
| Sayed 2008 | No | No | No | Yes | No | No | No | No | No | No | Yes | Yes | No | Yes | No | No | No | Yes | No |
| van Stuijvenberg 2001 | Yes | Yes | Yes | Yes | No | No | No | No | No | No | Yes | Yes | No | Yes | No | Yes | No | Yes | Yes |
| Rodrigues 2023 | Yes | Yes | Yes | Yes | Yes | Yes | No | Yes | No | Yes | No | No | No | No | No | Yes | Yes | Yes | Yes |

*See the 'List of Included Studies' IDs' above for the full bibliography.
